# Convergent and Divergent Neural Signatures of Cue-Induced Response in Substance and Behavioral Addictions: A Coordinate-Based Neuroimaging Meta-Analysis

**DOI:** 10.64898/2026.01.23.26344591

**Authors:** Qihong Zheng, Taoyu Wu, Xiaoqin Yang, Zifeng Wang, Jing Peng, Yuchen Huang, Yixuan Song, Xiao Lin, Anise M. S. Wu, Tianye Jia, Jie Shi, Yan Sun

## Abstract

Addiction-related cues trigger craving and precipitate relapse, yet whether substance use disorders (SUDs) and behavioral addictions (BAs) share a universal neurobiological cue-reactivity core remains unresolved. We conducted a coordinate-based meta-analysis using Seed-based d Mapping with Permutation of Subject Images (SDM-PSI) to synthesize whole-brain fMRI studies comparing visually evoked cue-reactivity in strictly diagnosed SUDs and BAs versus healthy controls. An updated systematic search through September 12, 2026, yielded 50 eligible articles comprising 53 independent contrasts and 2,578 participants. Pooled analyses across all addictions revealed convergent cue-elicited activations in the mid-cingulate cortex, precentral gyrus, and a midline midbrain region. However, a formal SUD ∩ BA conjunction analysis yielded no family-wise error (FWE)-corrected shared clusters. Direct comparisons demonstrated greater activation in the left hippocampus, pregenual anterior cingulate cortex, and precuneus for SUDs relative to BAs. Importantly, the left hippocampal and precuneus differences survived age adjustment; furthermore, the hippocampal finding remained robust when restricting analyses to highly controlled cue-versus-neutral paradigms, while the precuneus finding was retained within picture-cue modalities. Furthermore, exploratory analyses of clinical characteristics indicated that cue-elicited neural responses were associated with specific clinical states and treatment variables (e.g., methadone maintenance in heroin use disorder), highlighting the modulatory effect of clinical symptom profiles on these regional functional signatures. Overall, the results motivate further investigation of memory- and self-related processes, as well as control- or action-related processes. These findings offer a critical foundation for developing targeted clinical interventions, distinguishing between generalized transdiagnostic neuromodulation targets and disorder-specific therapies tailored to distinct clinical and demographic profile.

## 1. Introduction

Addiction is a chronic, relapsing brain disorder characterized by compulsive substance use or behaviors despite escalating negative consequences (1). The global burden of substance use disorders (SUDs) is substantial. The World Health Organization estimated that 400 million people worldwide were living with alcohol and drug use disorders in 2019, including 209 million with alcohol dependence (2). More recently, the United Nations Office on Drugs and Crime estimated that 64 million people had a drug use disorder in 2023, yet only approximately one in 12 received drug-related treatment (3). Parallel to SUDs, behavioral addictions (BAs) have emerged as a critical public health challenge (3–5). Disorders such as internet gaming disorder (IGD) and gambling disorder (GD) are now formally recognized in the International Classification of Diseases, 11th revision (ICD-11) (6). Recent data indicate a global IGD prevalence of 9.9% among youth, and the WHO separately estimates that approximately 1.2% of adults worldwide have GD in 2024 (7, 8).

Given these surging prevalence rates, the commonalities and differences of the neurobiological mechanisms underlying SUDs and BAs remain a subject of intense debate. Several reviews have suggested similar neurobiological mechanisms underlying SUDs and BAs, including dysregulation of regions related to executive function, reward systems, and emotional function (9). Furthermore, distinctions persist. SUDs are characterized by direct neurotoxic insults and profound neurotransmitter adaptations resulting from exogenous substance intake (10), whereas BAs appear more closely tied to maladaptive plastic changes in learning and memory systems, reflecting the complex skill acquisition inherent in gaming or gambling (11). Clinically, SUDs typically present with more severe executive dysfunction than BAs (12), and withdrawal in SUDs includes both affective and somatic distress (13, 14), while BAs withdrawal is primarily characterized by affective symptoms (15). However, few studies have directly compared the mechanisms underlying BAs with SUDs, with the vast majority of studies restricted to single-disorder cohorts, leaving the distinct and shared neural mechanisms between SUDs and BAs incompletely understood.

Cue-reactivity paradigms (CRP) serve as a vital method for investigating these mechanisms, using stimuli to elicit related neural responses and craving, and are considered one of the strongest predictors of relapse (16, 17). The basis of the CRP is rooted in associative learning accounts, positing that conditional cues previously paired with drug or addictive behaviors can evoke stimulus-associated responses, such as an intense urge to use drug or engage in the behavior (18). According to Incentive-Sensitization Theory (IST), re-exposure to addiction-related cues triggers an immediate, hyper-sensitized activation of the mesolimbic dopamine system, generating a powerful, bottom-up motivational drive manifested as an irrepressible urge of “wanting”, a pattern documented across both SUDs and BAs (19). Complementing this view, the Impaired Response Inhibition and Salience Attribution (iRISA) model suggests that cue-reactivity reflects a global imbalance between the brain’s cognitive control and value-assessment networks: addiction cues acquire “hyper-salience” that overrides natural reinforcers (e.g., food), a phenomenon primarily driven by (and reciprocally exacerbating) subjective craving that leads to diminished top-down inhibitory control, thereby creating a vicious cycle consistently observed across both SUDs and BAs (20).

Building on these theoretical frameworks, CRP engage a complex ensemble of neurocognitive systems, including attention, reward processing, and emotion regulation (21). Neuroimaging studies have consistently implicated the mesocorticolimbic system, including the ventromedial prefrontal cortex (vmPFC), anterior cingulate cortex (ACC), striatum, and amygdala, in substance craving and relapse risk across SUDs (22). Parallel dysregulation within the ventral striatal and prefrontal circuits has been documented in BAs (23, 24), alongside a functional shift toward the dorsal striatum as behavior transitions from impulsive to compulsive habits in both disorder classes (24–27). Recent comprehensive meta-analyses have significantly advanced our understanding of addiction neurobiology; for instance, Roberge et al. (28) examined alterations across task domains in SUDs, whereas Fascher et al. (29) investigated factors that modulate drug cue reactivity within SUDs. Other syntheses have compared SUDs with behavioral addictions using resting-state connectivity or aggregated data across multiple task domains, including cue reactivity, reward processing, and cognitive control (30, 31). However, these broader syntheses leave a more focused question unresolved: whether SUDs and clinically recognized behavioral addiction phenotypes show convergent or divergent neural responses when examined within a comparable cue-reactivity framework. Addressing this requires isolating the visual cue-reactivity domain to reduce confounding variance from different cognitive tasks and sensory modalities. Furthermore, substantial methodological heterogeneity across original studies, such as variations in clinical sub-populations, image acquisition parameters, and statistical modeling pipelines, has contributed to inconsistent findings in the literature. For instance, studies have reported conflicting hyperactivation and hypoactivation within the middle frontal gyrus activation in alcohol use disorder (AUD) (32, 33) and anterior cingulate cortex in IGD (23, 34). These discrepancies underscore the critical need for a large-scale, methodologically rigorous quantitative synthesis to clarify cross-diagnostic cue-reactivity patterns.

To address these limitations, we conducted a coordinate-based functional magnetic resonance imaging (fMRI) meta-analysis restricted to visual cue-reactivity paradigms. Notably, this synthesis focuses exclusively on functional neuroimaging studies utilizing visual cue-reactivity paradigms due to focusing on one sensory modality improves the comparability of SUD and BA studies. Furthermore, while SUDs involve multimodal cues, BAs such as IGD and GD are primarily driven by visual stimuli. Consequently, this focus on visual cues provides the most comparable and standardized modality for directly evaluating the shared and distinct neural signatures between SUDs and BAs, consistent with the dominant paradigms in contemporary addiction research (35).

We aim to: (1) clarify the shared and distinct neural substrates of SUDs and BAs; (2) compare the neuroimaging profiles of AUD, heroin use disorder (HUD), and IGD; and (3) explore brain-behavior correlations with clinical metrics. By comparing SUDs with BAs under the same task framework, we sought to assess whether cue-related abnormalities extend across addiction categories and to identify patterns that may differ between them. Such evidence may help distinguish candidate processes associated with addictive behavior across diagnostic groups from responses that vary with substance exposure or compulsive behavior-specific features. Clinically, these findings may guide hypotheses about shared and disorder-specific mechanisms for future longitudinal and intervention studies.

## 2. Methods

### 2.1 Search strategy and selection criteria

This study adhered to the PRISMA 2020 (Preferred Reporting Items for Systematic Reviews and Meta-Analyses) statement (36) and was prospectively registered on PROSPERO (CRD42024603551).

A systematic literature search was performed across PubMed, Cochrane, Embase, PsycINFO, Scopus, and Web of Science for studies published up to September 12, 2026. The search utilized Boolean logic to combine terms related to “Addiction”, “Cue-reactivity”, “Neuroimaging”, “Control”, and “Participant”. Because IGD and GD are classified as disorders due to addictive behaviors in ICD-11, our search was restricted to these conditions. To maintain strict diagnostic boundaries and clarity of comparison, studies focused on everyday food craving, obesity, binge-eating disorder, or proposed ‘food addiction’ were not eligible. These conditions lack a formalized placement within recognized addictive disorder categories, and their hybrid nature—involving both substance ingestion and behavioral engagement—precludes their unambiguous classification as either substance-based or purely behavioral addictive disorders. Detailed search strategies are provided in Table S1.

Studies were included if they met the following criteria: 1) Eligibility required the clinical group to have an explicitly reported formal diagnosis of a SUD or BA, or documented fulfillment of established diagnostic criteria for the relevant disorder. Participants who were abstinent at the time of scanning remained eligible for inclusion, provided that a qualifying formal diagnosis had been established prior to their abstinence. 2) Utilized fMRI with a visual cue-reactivity task. 3) Reported whole-brain group addiction vs. control comparisons in standardized stereotactic space (Montreal Neurological Institute (MNI) (37) or Talairach (38)). Cue-reactivity was defined to encompass three group-comparative contrast types in this study: addicts vs. controls under ⍰ addiction cues vs. neutral cues, ⍰ addiction cues alone, or ⍰ addiction cues vs. baseline. 4) Provided essential statistical data, including activation foci coordinates, *t*/*z*/*p*-values, and sample sizes.

Studies were excluded based on the following: 1) studies based solely on food craving, obesity, binge-eating disorder, or proposed food addiction; 2) non-empirical publications (e.g., reviews, conference abstracts); 3) reporting only region-of-interest (ROI) results; 4) unretrievable full texts.

### 2.2 Screening procedure and data extraction

Following PRISMA, 50 articles (2,578 participants) were included (Fig. 1a). Given that some studies provided separate contrasts for distinct participants groups, these 50 articles yielded a total of 53 independent contrasts, comprising 37 SUDs contrasts (13 HUD, 13 AUD, 2 each for nicotine use disorder (NUD), methamphetamine use disorder (MUD), cannabis use disorder (CanUD), cocaine use disorder (CoUD), and 1 each for betel-quid dependence (BQD), ketamine use disorder (KUD), and combined stimulant use disorder (included cocaine and amphetamine)) and 16 BAs contrasts (12 IGD and 4 GD) (Fig. 1b). Specifically, Wei et al., 2021 (39) and Tabatabaei et al., 2014 (40) each contributed separate contrasts for protracted abstinence (PA) and methadone maintenance (MMT) groups; Kober et al., 2016 (41) contributed separate contrasts for CoUD and GD. For studies comprising multiple patient cohorts sharing a common control group, the control data were counted only once in overall analyses to prevent duplicate counting.

**Figure 1.**
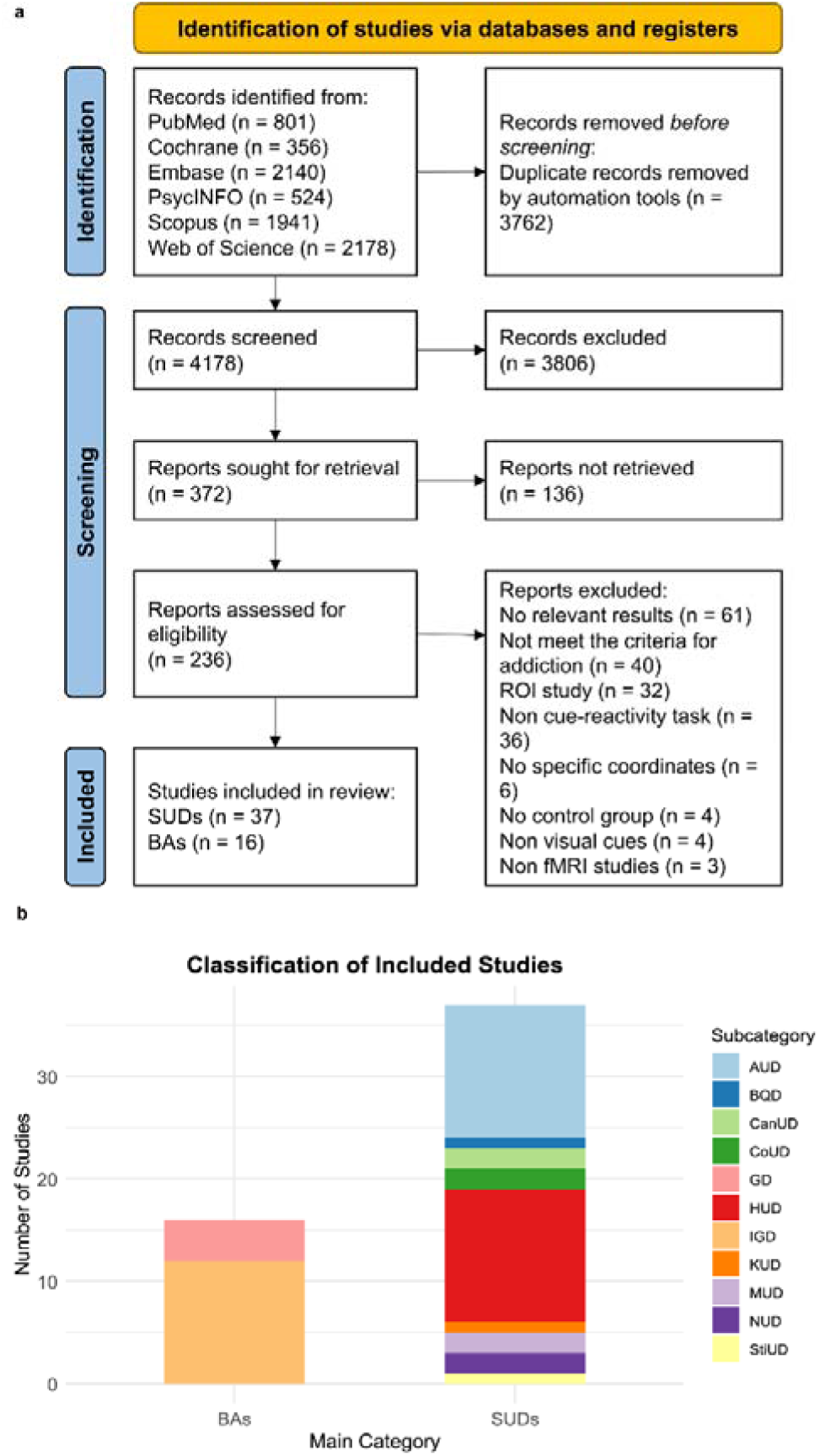
PRISMA flowchart and the composition of included studies. (a) PRISMA 2020 flow diagram depicting the identification, screening, eligibility assessment, and inclusion of studies retrieved from six electronic databases, updated through September 12, 2026. After the removal of duplicates, 4,178 records were screened, 236 full-text articles were assessed for eligibility, and 50 unique studies met the strict diagnostic inclusion criteria (which rigorously excluded sub-clinical heavy use and conceptually ambiguous conditions such as food addiction). (b) Classification of the included literature into SUDs and BAs, yielding 53 independent contrasts from the 50 unique studies. The SUDs category comprises 37 contrasts across 9 subtypes, and the BAs category includes 16 contrasts across 2 subtypes. Abbreviations: SUDs, substance use disorders; BAs, behavioral addictions; AUD, alcohol use disorder; HUD, heroin use disorder; NUD, nicotine use disorder; MUD, methamphetamine use disorder; CanUD, cannabis use disorder; BQD, betel-quid dependence; CoUD, cocaine use disorder; KUD, ketamine use disorder; StiUD, stimulant use disorder; IGD, internet gaming disorder; GD, gambling disorder.

Brain imaging, diagnostic, and clinical data were extracted following standardized guidelines (42, 43). For each clinical group, we recorded the specific diagnostic classification and the standardized criteria used to establish it. Abstinence status and duration, treatment history, and specific interventions (e.g., methadone maintenance status or dosage) at the time of scanning were extracted when available. Detailed extraction information is provided in Tables S2 and S3. Clinical effect sizes were quantified using Hedges’ *g* to correct for small-sample bias (44), with all calculations performed in R (v4.3.0) via the *metafor* package (45). To ensure comparability across studies in subsequent meta-regression analyses, all time-related variables reported in different units (e.g., weeks, months) were converted to a standard unit of days.

Two sets of researchers independently performed the study screening and data extraction, respectively. For study screening, two researchers evaluated titles, abstracts, and full texts against the eligibility criteria. For data extraction, a separate pair of researchers collected data using a standardized form. In both phases, discrepancies were resolved through consensus, with unresolved disagreements adjudicated by the wider research team.

### 2.3 Quality assessment

The quality of neuroimaging studies was evaluated using a 12-point checklist adapted for task-based fMRI (46, 47). Assessment results are provided in Table S5.

### 2.4 Meta-analysis procedure

Coordinate-based meta-analysis was performed using Seed-based d Mapping with Permutation of Subject Images (SDM-PSI, v6.23) (42). This tool reconstructs study-level maps to approximate original participant activation patterns (48). Unlike conventional coordinate-based meta-analytic methods that treat all reported peaks equivalently, SDM-PSI employs a multiple imputation and bounded effect-size approach that imposes a penalized upper bound on reconstructed effect-size maps for studies employing relatively liberal thresholds, ensuring that such studies are appropriately down-weighted and do not disproportionately inflate global or subgroup effect-size estimates. Preprocessing was conducted within a gray-matter mask using an isotropic Gaussian kernel with a full width at half maximum (FWHM) of 20 mm, on a standard template with 2 mm isotropic voxels (49). For mean analyses, permutation tests were configured with 50 imputations and 1000 permutations. Family-wise error (FWE) correction was subsequently applied using 1000 permutations.

A hierarchical thresholding strategy was implemented to balance inferential rigor with exploratory transparency. Primary results in the main text are based exclusively on FWE-corrected *p* < 0.05 using threshold-free cluster enhancement (TFCE) (43), combined with a cluster-extent threshold of k > 20 voxels. Additionally, exploratory results based on an uncorrected threshold of *p* < 0.005 with k > 20 voxels are provided in the Supplementary Materials as a heuristic reference for future fMRI studies on addiction phenotypes (50), consistent with practice in rigorous quantitative neuroimaging syntheses (51), and should be interpreted with appropriate caution.

#### 2.4.1 General and subgroup meta-analyses

A general meta-analysis was performed for the combined addiction group (SUDs ∪ BAs) and controls. Subsequently, subgroup meta-analyses were conducted separately by mean analysis for SUDs, BAs, AUD, HUD, and IGD. Each subgroup analysis included only studies belonging to the respective category.

#### 2.4.2 Conjunction and contrast meta-analyses

To delineate shared neural signatures, conjunction analyses (e.g., SUDs ∩ BAs) were performed using the SDM multimodal utility. Differential activations were identified through contrast meta-analyses using linear models with addiction category as a binary moderator (coded 0/1). Exploratory post-hoc meta-regressions examined associations between whole-brain activation patterns and continuous study-level clinical variables. For clarity, these variables are presented in three conceptual domains: (1) addiction involvement, exposure, and clinical-course indicators, including duration of substance use, total substance consumption, daily dosage, weekly gaming hours, withdrawal duration, and treatment duration or methadone dosage; (2) addiction symptom-severity scores; and (3) emotional-symptom measures, including depression and anxiety. To ensure statistical robustness, meta-regression results are reported in the main text only where the contributing study count n (number of studies) > 5 and between-study heterogeneity (*I²*) ≤ 50%. Analyses failing either criterion are provided in the Supplementary Materials and should be interpreted with appropriate caution.

### 2.5 Demographic covariate analyses

We conducted two additional sensitivity analyses to examine the influence of demographic composition on diagnostic comparisons. First, we re-estimated the four direct diagnostic contrasts (SUDs vs. BAs, HUD vs. AUD, HUD vs. IGD, and AUD vs. IGD) with combined-sample mean age entered as a study-level covariate. Second, given the predominance of male participants in the included studies, we repeated applicable analyses using only contrasts in which both the patient and control groups comprised male participants.

Study-level combined demographic metrics were calculated for each included study. Specifically, the combined total mean age (*Total_Age*) and its corresponding pooled standard deviation (*Total_Age_sd*) were mathematically reconstructed from the separately reported descriptive statistics of the patient and control groups using the following formulae:

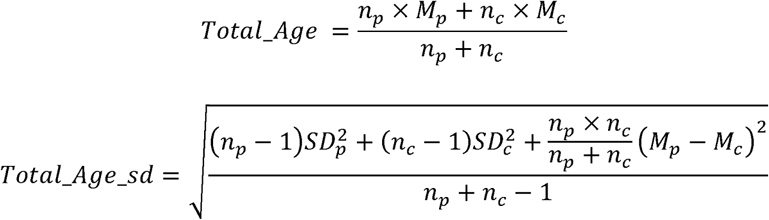

where *n_p_* and *n*_C_ denote the sample sizes of the patient and control groups, respectively; *M_p_* and *M_c_* represent the respective mean ages; and *SD_p_* and *SD_c_* signify the respective standard deviations of age. Following identical logic, the combined male proportion was calculated as:

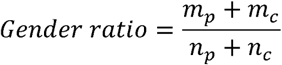

where *m_p_* and *m_C_* represent the number of male individuals within the patient and control groups respectively.

For each paired comparison block and each demographic metric, a separate sample-size-weighted linear regression model was constructed in R (v4.3.0), with diagnostic subgroup membership as the predictor and the study-level combined-sample mean age or male proportion as the outcome. Each study was weighted by its total sample size (*N_total_ = n_p_ + n_C_*) to ensure that larger and statistically more stable studies exerted proportionally greater influence on the pooled estimates. Subgroup-specific weighted means and 95% confidence intervals (CIs) were estimated, and the inter-group difference (*β*), *t*-statistic, and two-tailed *p*-value were derived to test for statistical significance.

These demographic comparisons were descriptive and were not used to determine covariate inclusion on the basis of statistical significance. In the separate age-adjusted sensitivity analyses, diagnostic subgroup was entered as the variable of interest and combined-sample mean age as a study-level covariate in each direct diagnostic comparison. Separately, to examine sensitivity to sex composition, applicable analyses were repeated using only contrasts in which both the patient and control groups comprised male participants.

### 2.6 Sensitivity analyses

To examine the robustness of the primary findings to variation in fMRI contrast and cue modality, we conducted separate sensitivity analyses. First, analyses were restricted to the contrast of “addiction cues versus control cues”. Second, studies using picture and video cues were analyzed separately. Because relatively few contrasts used video cues, the video-cue analysis was limited to the pooled addiction group. Finally, to explore the potential influence of methadone maintenance treatment (MMT) within HUD, we analyzed HUD contrasts with and without MMT separately and compared their resulting neural patterns. All sensitivity analyses applied the same hierarchical thresholding criteria as the primary analyses.

### 2.7 Heterogeneity, publication bias, visualization and functional decoding

Heterogeneity and publication bias were assessed based on the peak coordinates (52). The level of study heterogeneity quantified by the *I^2^*statistic was interpreted as low (*I^2^* ≤ 25%), moderate (25% < *I^2^*≤ 50%), substantial (50% < *I^2^* ≤ 75%), or considerable (*I^2^* > 75%) (53). Publication bias was assessed for each significant contrast via Egger’s test (*p* < 0.1) (54).

Coordinates were localized using the xjView toolbox (https://www.alivelearn.net/xjview) and the Automated Anatomical Labeling (AAL) atlas (55). Three-dimensional brain models were rendered using BrainNet Viewer (56). Finally, to interpret the functional significance of the identified meta-analytic peaks, we performed location-based functional decoding using NiMARE with the Neurosynth v7 database. For each reported coordinate, studies reporting at least one peak within a 6-mm radius were identified, and terms from Neurosynth abstract-based TF-IDF annotations were ranked using reverse-inference z-scores (zReverse). The five highest-ranked terms remaining after anatomical and structural terms were excluded were retained. These terms represent descriptive associations and were not treated as statistically significant results.

## 3. Results

### 3.1 Demographic characteristics

Sample-size-weighted demographic characteristics, calculated from the combined patient and control samples within each contrast, are summarized in Table 1; contrast-level data are provided in Table S3. Age data were available for 52 contrasts and male-proportion data for 50 contrasts. The weighted mean age was 35.86 years for SUDs and 24.49 years for BAs. The weighted male proportion ranged from 71.05% in AUD to 93.11% in HUD.

**Table 1.** Sample-size-weighted demographic characteristics of included studies.

| Variable | SUDs (Mean<br>[95% CIs]) | BAs (Mean<br>[95% CIs]) | HUD (Mean<br>[95% CIs]) | AUD (Mean<br>[95% CIs]) | IGD (Mean<br>[95% CIs]) |
| --- | --- | --- | --- | --- | --- |
| Total Age<br>(years) | 35.86 [33.40,<br>38.33] | 24.49 [21.58,<br>27.40] | 35.95 [33.66,<br>38.24] | 40.67 [35.19,<br>46.15] | 21.81 [20.34,<br>23.29] |
| Gender<br>ratio (%<br>male) | 82.86%<br>[76.78%,<br>88.95%] | 80.96%<br>[68.13%,<br>93.79%] | 93.11%<br>[84.91%,<br>100.00%] | 71.05%<br>[60.03%,<br>82.07%] | 80.59%<br>[64.15%,<br>97.04%] |

Values are sample-size-weighted contrast means [95% confidence intervals (CIs)] from separate intercept-only weighted linear models. Each contrast combines patients and controls and is weighted by $n_p + n_c$ . The HUD male proportion CI upper limit was capped at 100%. Age data were available for 52 contrasts and sex data for 50. SUDs: substance use disorders; BAs: behavioral addictions; HUD: heroin use disorder; AUD: alcohol use disorder; IGD: internet gaming disorder.

Sample-size-weighted linear regression showed that the SUDs had a higher mean age than the BAs (β = 11.37, *t* = 5.18, *p* < 0.001; Table S4). The age difference between HUD and AUD was not statistically significant (β = -4.72, *t* = -1.73, *p* = 0.097). Both HUD and AUD had higher mean ages than IGD (HUD vs. IGD: β = 14.14, *t* = 11.08, *p* < 0.001; AUD vs. IGD: β = 18.86, *t* = 6.90, *p* < 0.001).

The male proportion was higher in HUD than in AUD contrasts (β = 22.05, *t* = 3.51, *p* = 0.002). The other comparisons did not reach statistical significance (SUDs vs. BAs: β = 1.90, *t* = 0.30, *p* = 0.769; HUD vs. IGD: β = 12.51, *t* = 1.64, *p* = 0.116; AUD vs. IGD: β = −9.54, *t* = −1.11, *p* = 0.282).

### 3.2 Common and distinct neural signatures between SUDs and BAs

The pooled analysis across all addiction types (SUDs ∪ BAs) (n =53) identified significant hyperactivation in the right mid-cingulate cortex (MCC, peak: 4, -18, 30), left precentral gyrus (PreCG, peak: -46, 8, 32), and midbrain (PAG vicinity) (peak: 0, -22, -6) during cue-reactive responding (Fig. 2a). Neurosynth decoding associated the MCC peak with response-inhibition terms, the PreCG peak with phonological and reading-related terms, and midbrain (PAG vicinity) peak with reward- and fear-conditioning terms (Table S6). These associations suggest that the pooled cue-reactivity pattern may involve control, stimulus-related, and motivational-affective processes.

**Figure 2.**
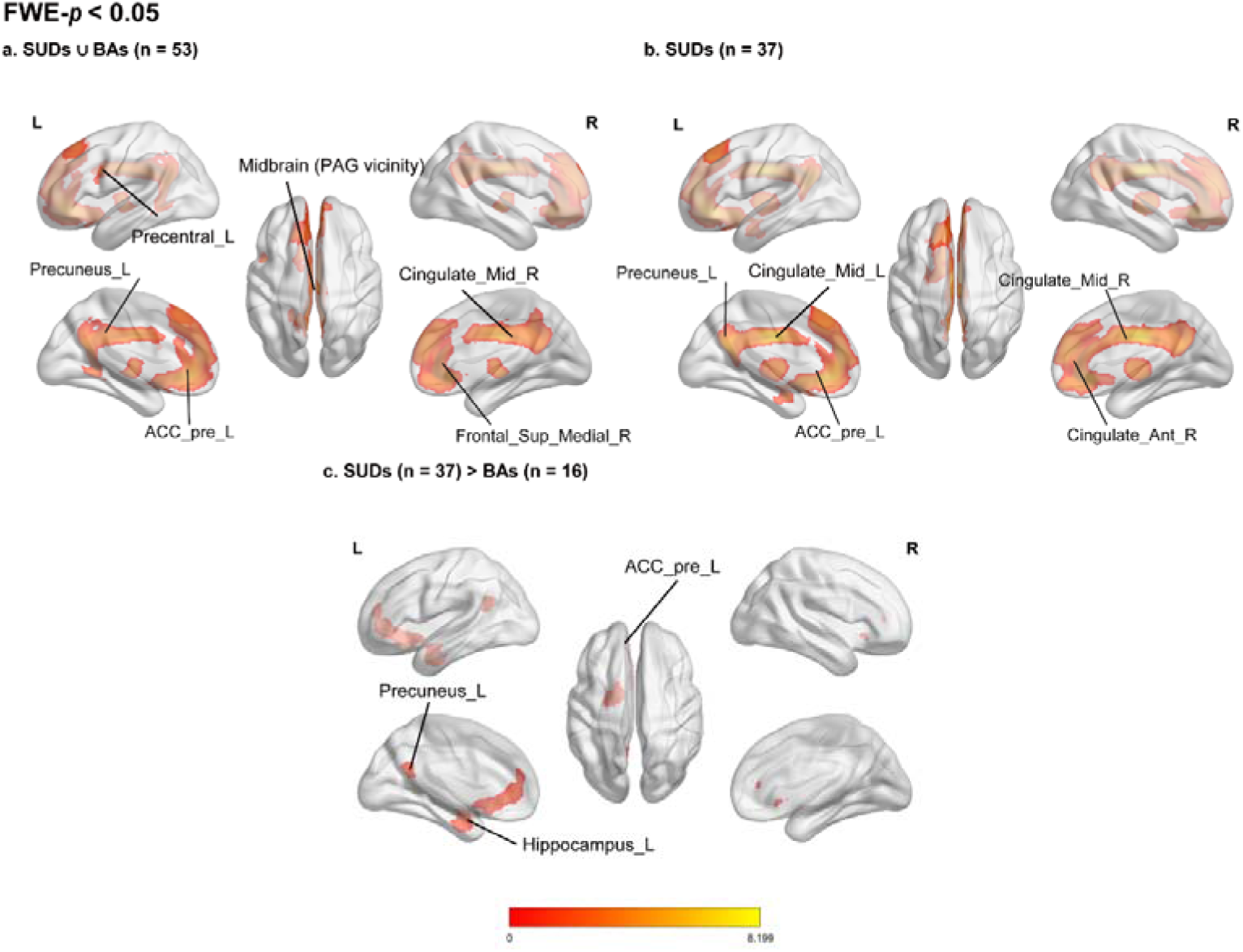
Common and distinct neural activation patterns during cue reactivity in addictions. Maps show FWE-corrected results (p < 0.05). (a) The pooled SUDs ∪ BAs analysis (n = 53 contrasts) shows annotated clusters in the left precentral gyrus (Precentral_L), left precuneus (Precuneus_L), left anterior cingulate cortex (ACC_pre_L), right middle cingulate gyrus (Cingulate_Mid_R), right medial superior frontal gyrus (Frontal_Sup_Medial_R), and a midbrain cluster (PAG vicinity). (b) The SUDs analysis (n = 37) shows annotated clusters in the left precuneus (Precuneus_L), left anterior cingulate cortex (ACC_pre_L), left and right middle cingulate gyri (Cingulate_Mid_L and Cingulate_Mid_R), and right anterior cingulate gyrus (Cingulate_Ant_R). (c) The direct contrast SUDs (n = 37) > BAs (n = 16) shows greater responses in SUDs in the left anterior cingulate cortex (ACC_pre_L), left precuneus (Precuneus_L), and left hippocampus (Hippocampus_L). Maps are displayed on a standard MNI template in neurological orientation (L = left hemisphere). The warm color bar indicates positive SDM-Z values.

The SUDs-specific (n = 37) hyperactivation was observed in the left MCC (peak: -2, -12, 32) (Fig. 2b). Neurosynth terms associated with this peak were broad, including personality-trait-related terms, and did not permit a specific cognitive attribution (Table S6). Its proximity to the MCC peak in the pooled analysis suggests that SUD studies may contribute substantially to that pooled finding, but does not establish an absence of MCC involvement in BAs.

The direct comparison revealed significantly greater cue-related responses in SUDs than in BAs in the left hippocampus (HIP, peak: -20, -10, -22), left pregenual anterior cingulate cortex (ACCpre, peak: -8, 42, 2), and left precuneus (PCUN, peak: -4, -56, 22) (Fig. 2c). Neurosynth decoding associated the HIP peak with emotional and face-related terms, the ACCpre peak with monetary reward, autobiographical, and regulation-related terms, and the PCUN peak with autobiographical memory and default mode network terms (Table S6). Together, these associations are consistent with greater recruitment of affective and self-referential memory-related regions during cue reactivity in SUDs than in BAs. The inverse contrast (BAs > SUDs) did not yield any significant clusters.

Neither the BA-only analysis (n = 16) nor the SUDs ∩ BAs conjunction yielded FWE-corrected clusters. At the exploratory uncorrected threshold (p < 0.005), the BA-only analysis showed right triangular inferior frontal and left middle frontal clusters, together with left calcarine and cerebellar clusters (Fig. S1a; Table S6). To maintain analytical transparency, exploratory uncorrected findings (*p* < 0.005), comprehensive coordinates, statistical indices, and expanded functional decoding annotations for all sub-threshold clusters are comprehensively documented in the Supplementary Material (Table S6).

### 3.3 Subgroup-specific comparisons (AUD, HUD, and IGD)

Across the three diagnostic subgroups, suprathreshold hyperactivation surviving FWE correction was uniquely observed within the HUD group (n = 13). This profile encompassed the right thalamus (THA, peak: 6, -8, 4), left ACCpre (peak: 0, 44, 10), right orbital inferior frontal gyrus (ORBinf, peak: 44, 34, -8), left cerebellum (CC, Crus I; peak: -16, -76, -24), left superior parietal gyrus (SPG, peak: -22, -66, 50), and left inferior parietal gyrus (IPL, peak: -46, -50, 48) (Fig. 3a). Neurosynth decoding linked the THA peak to pain and reward terms, the ACCpre peak to default mode and affective terms, and the ORBinf peak to emotional reappraisal (Table S7). The SPG and IPL peaks were associated with visuospatial attention and working-memory terms, whereas decoding of the cerebellar peak did not indicate a specific cognitive process (Table S7). This distributed pattern may reflect the involvement of motivational-affective and attention-related processes in heroin cue reactivity. In contrast, no activation clusters surviving FWE correction were observed within the AUD (n = 13) or IGD (n = 12) subgroups.

**Figure 3.**
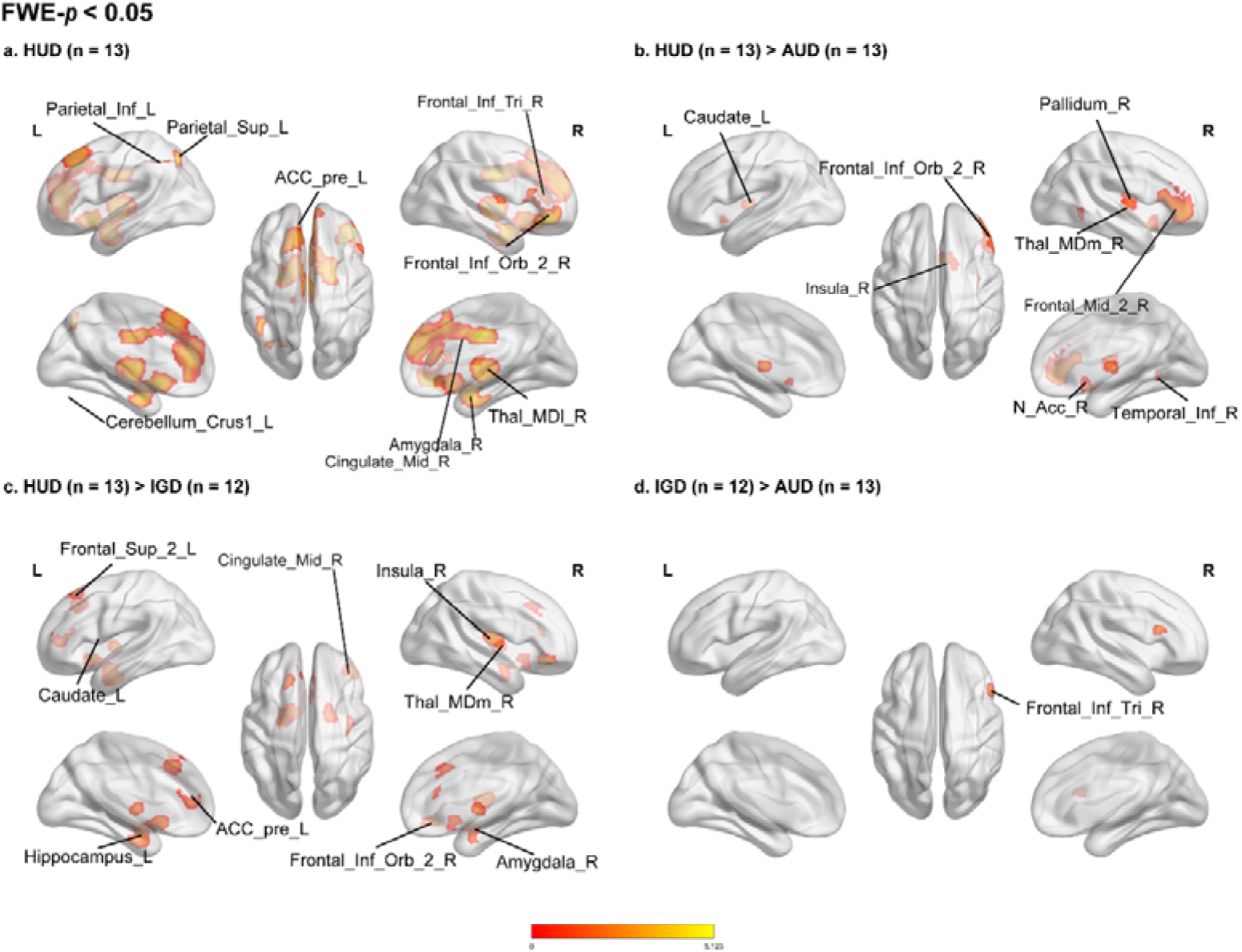
Subtype-specific and comparative neural activation patterns during cue reactivity. Maps show FWE-corrected results (p < 0.05). (a) The HUD subgroup (n = 13) shows annotated clusters in the left inferior and superior parietal regions (Parietal_Inf_L and Parietal_Sup_L), left anterior cingulate cortex (ACC_pre_L), right triangular and orbital inferior frontal gyri (Frontal_Inf_Tri_R and Frontal_Inf_Orb_2_R), right amygdala (Amygdala_R), right middle cingulate gyrus (Cingulate_Mid_R), right mediodorsal thalamus (Thal_MDl_R), and left cerebellar Crus I (Cerebellum_Crus1_L). (b) HUD (n = 13) > AUD (n = 13) shows greater responses in HUD in the left caudate (Caudate_L), right insula (Insula_R), right orbital inferior frontal gyrus (Frontal_Inf_Orb_2_R), right mediodorsal thalamus (Thal_MDm_R), right pallidum (Pallidum_R), right nucleus accumbens (N_Acc_R), right inferior temporal gyrus (Temporal_Inf_R), and right middle frontal gyrus (Frontal_Mid_2_R). (c) HUD (n = 13) > IGD (n = 12) shows greater responses in HUD in the left superior frontal gyrus (Frontal_Sup_2_L), left caudate (Caudate_L), left anterior cingulate cortex (ACC_pre_L), left hippocampus (Hippocampus_L), right insula (Insula_R), right mediodorsal thalamus (Thal_MDm_R), right orbital inferior frontal gyrus (Frontal_Inf_Orb_2_R), and right amygdala (Amygdala_R). (d) IGD (n = 12) > AUD (n = 13) shows a cluster in the right triangular inferior frontal gyrus (Frontal_Inf_Tri_R). Maps are displayed on a standard MNI template in neurological orientation. The warm color bar indicates positive SDM-Z values.

Direct pairwise sub-group comparisons further delineated these clinical phenotypes. The HUD > AUD contrast revealed significantly heightened cue-induced responses (CIR) in HUD within the right ORBinf (peak: 48, 36, -8), right pallidum (PAL, peak: 22, 4, 2), right nucleus accumbens (NAcc, peak: 8, 16, -6), right THA (peak: 4, -12, 4), right inferior temporal gyrus (ITG, peak: 62, -58, -10), and left caudate (CAU, peak: -10, 8, 10) (Fig. 3b). Decoding associated the ORBinf with emotional reappraisal, the NAcc with incentive and reward anticipation, and the PAL and CAU with basal-ganglia, motor, and reward-related terms (Table S7). The THA peak was linked to reward and pain terms, whereas the ITG terms were not sufficiently specific for a clear functional attribution (Table S7). The pattern is compatible with greater recruitment of regions associated with incentive processing and action-related functions in HUD than in AUD, without establishing a treatment- or substance-specific mechanism.

Compared with IGD, HUD showed greater responses in the left HIP (peak: -18, -8, -20), left ACCpre (peak: -6, 44, 8), right insula (INS, peak: 36, -16, 14), right ORBinf (peak: 44, 34, -8), right amygdala (AMYG, peak: 20, -4, -16), right THA (peak: 6, -8, 2), left superior frontal gyrus (SFG, peak: -18, 24, 54), and left CAU (peak: -8, 4, 8) (Fig. 3c). Neurosynth associated the HIP and AMYG peaks with emotional and face-related terms; the ACCpre with default mode and appraisal terms; the INS with pain and nociception; the ORBinf with emotional reappraisal; and the THA and CAU with reward-related terms (Table S7). The SFG decoding was less specific. These findings may point to stronger affective-memory and reward-related responses in HUD.

For the AUD versus IGD comparison, IGD showed a greater cue-related response in the right triangular inferior frontal gyrus (IFGtriang, peak: 50, 20, 20) (Fig. 3d). Its Neurosynth terms were broad, including emotional and social terms, and do not support assigning this isolated finding to a specific cognitive operation (Table S7).

Finally, no conjunction analyses (HUD ∩ AUD, HUD ∩ IGD, or AUD ∩ IGD) yielded clusters that withstood FWE correction. To ensure comprehensive documentation, exploratory uncorrected findings (*p* < 0.005) along with exhaustive coordinates, statistical values, and expanded functional decoding annotations for all sub-threshold clusters are comprehensively tabulated in the Supplementary Material (Table S7).

### 3.4 Clinical correlations

All meta-regression analyses reported below are designated as exploratory post-hoc analyses. The number of contributing studies (n) and between-study heterogeneity (*I²*) for each analysis are reported in Table S8.

Among addiction involvement and clinical-course indicators, withdrawal duration provided a notable cross-diagnostic comparison. In HUD, longer withdrawal duration (n = 7) was negatively associated with cue-reactivity effects in the left medial superior frontal gyrus (SFGmed; peak: -2, 44, 24), whereas in AUD it was positively associated with effects in the same broad anatomical region (n = 7; peak: -6, 54, 30) and in the left mid-cingulate cortex (peak: -4, -46, 34) (Fig. 4a,b). Thus, the study-level associations had opposite directions in nearby left SFGmed locations. The peaks were not identical, however this pattern should therefore be interpreted descriptively rather than as a statistically established difference between diagnostic groups. Neurosynth decoding returned network-level and mentalizing-related terms for the HUD SFGmed peak, mentalizing-related terms for the AUD SFGmed peak, and default-mode-network terms for the AUD MCC peak (Table S8).

**Figure 4.**
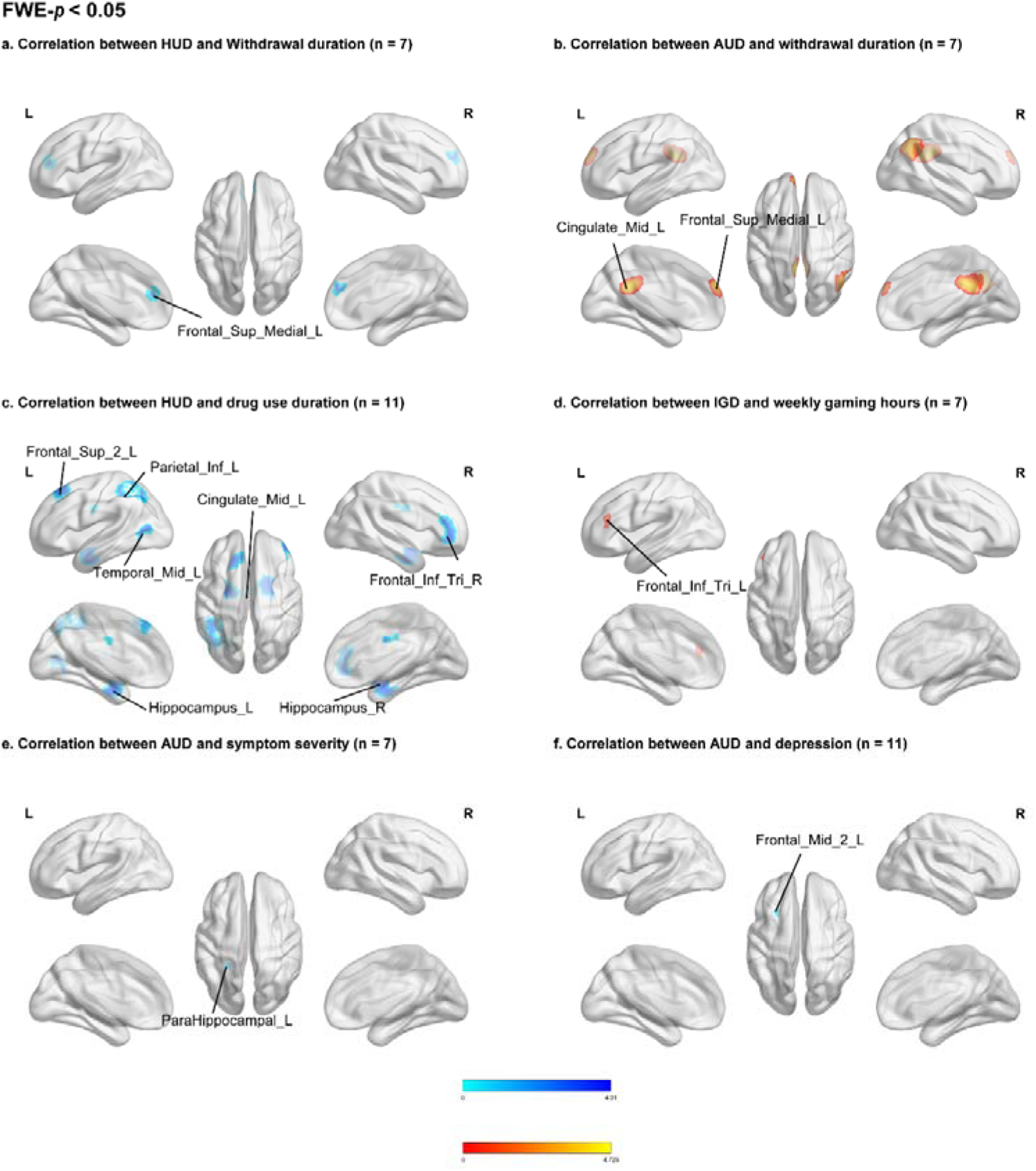
Study-level meta-regression of clinical measures and cue-reactivity effects. Maps show FWE-corrected results (p < 0.05). (a) In HUD, withdrawal duration (n = 7) was negatively associated with effects in the left medial superior frontal gyrus (Frontal_Sup_Medial_L). (b) In AUD, withdrawal duration (n = 7) was positively associated with effects in the left middle cingulate gyrus (Cingulate_Mid_L) and left medial superior frontal gyrus (Frontal_Sup_Medial_L). (c) In HUD, drug-use duration (n = 11) was negatively associated with effects in the left superior frontal gyrus (Frontal_Sup_2_L), left inferior parietal lobule (Parietal_Inf_L), left middle cingulate gyrus (Cingulate_Mid_L), left middle temporal gyrus (Temporal_Mid_L), right triangular inferior frontal gyrus (Frontal_Inf_Tri_R), and bilateral hippocampi (Hippocampus_L and Hippocampus_R). (d) In IGD, weekly gaming hours (n = 7) were positively associated with effects in the left triangular inferior frontal gyrus (Frontal_Inf_Tri_L). (e) In AUD, symptom severity (n = 7) was negatively associated with effects in the left parahippocampal gyrus (ParaHippocampal_L). (f) In AUD, depression (n = 11) was negatively associated with effects in the left middle frontal gyrus (Frontal_Mid_2_L). Maps are displayed on a standard MNI template in neurological orientation. Cool and warm color bars indicate negative and positive associations, respectively.

Other indicators showed distinct associations. Longer chronological duration of drug use in HUD (n = 11) was negatively associated with cue-reactivity effects in the left hippocampus (-18, -10, -22), right hippocampus (22, -6, -20), left inferior parietal gyrus (-44, -50, 48), right triangular inferior frontal gyrus (42, 42, -2), left superior frontal gyrus (-18, 26, 52), left middle temporal gyrus (-48, -72, 10), and left mid-cingulate cortex (0, -8, 34) (Fig. 4c). Neurosynth decoding associated the bilateral hippocampal peaks with emotional-processing terms, the inferior parietal peak with working-memory terms, the superior frontal peak with default-mode and control-network terms, the middle temporal peak with visual-motion terms, and the mid-cingulate peak with pain- and impulsivity-related terms; terms for the IFGtriang peak were less specific (Table S8). In IGD, greater weekly gaming hours (n = 7) were positively associated with effects in the left triangular inferior frontal gyrus (-44, 36, 16; Fig. 4d), for which decoding yielded phonological and semantic-retrieval terms (Table S8).

For addiction symptom severity, greater AUD symptom severity (n = 7) was negatively associated with cue-reactivity effects in the left parahippocampal gyrus (peak: -24, -42, -6; Fig. 4e, Table S8). Neurosynth decoding returned scene-, navigation-, and autobiographical-memory-related terms for this peak. No FWE-corrected clusters were identified for IGD symptom severity (n = 10). For emotional symptoms, greater depressive symptom severity in AUD (n = 11) was negatively associated with effects in the left middle frontal gyrus (peak: -26, 18, 42; Fig. 4f, Table S8). Neurosynth decoding returned autobiographical-memory, recall, and default-network terms for this peak. No FWE-corrected clusters were identified for AUD anxiety severity (n = 5).

No FWE-corrected clusters were identified for HUD total drug consumption (n = 6) or treatment duration (n = 6). No clusters even at the uncorrected threshold (p < 0.005) were identified for HUD daily dosage (n = 8) or average daily methadone dosage (n = 5). All exploratory uncorrected findings, coordinates, statistical values, and functional-decoding annotations are provided in Table S8.

### 3.5 Demographic sensitivity analyses

#### 3.5.1 Age-adjusted diagnostic comparisons

The age-adjusted AUD (n = 12)–IGD (n = 12) comparison showed the most pronounced change in spatial pattern among the diagnostic contrasts, which is also the pair with the largest study-level age difference. In the primary analysis, IGD > AUD was represented by a single right IFGtriang cluster. After age adjustment, this peak shifted to the middle frontal gyrus, and several clusters emerged for AUD > IGD. Thus, adjustment was accompanied by a marked reorganization of the AUD–IGD results, rather than simple attenuation of the primary finding. However, this pattern does not establish that age alone caused the change.

The other comparisons also showed varying sensitivity. In the SUDs (n=36) > BAs (n=16) comparison retained FWE-corrected clusters in the left hippocampus and precuneus, while the anterior cingulate peak shifted from the pregenual to the subgenual region, and an additional left superior frontal cluster emerged. In the HUD (n=13) > AUD (n=12) comparison, four clusters remained, including the right orbital inferior frontal gyrus and pallidum. By contrast, no clusters survived FWE correction in the age-adjusted HUD (n = 13) > IGD (n = 12) comparison, compared with eight in the primary analysis. Overall, these changes indicate that some subgroup findings are sensitive to age adjustment, particularly given the strong association between age and diagnostic category. The results are reported in Figure S6A and Table S9.

#### 3.5.2 Male-only sensitivity analysis of sex composition

The male-only analyses included 20 SUD and 10 BA contrasts, and 10 HUD and 7 IGD contrasts. In the category comparison, SUDs showed greater effects in the right insula, whereas BAs showed greater effects in the left precentral gyrus. In the HUD-IGD comparison, HUD showed greater effects in the right insula, while IGD showed greater effects in the left precentral and right middle occipital gyri. The primary hippocampal, anterior cingulate, and precuneus findings were not reproduced in the male-only category comparison. Because restricting the analysis to male-only samples also altered the composition of contributing studies, these results indicate sensitivity to overall sample characteristics and should not be interpreted as isolating a pure biological sex effect (Figure S6B and Table S9).

### 3.6 Sensitivity analyses

#### 3.6.1 Analyses restricted to addiction cues versus control cues

To address contrast-definition heterogeneity, we restricted the dataset exclusively to studies comparing addiction cues directly against neutral control cues. This restricted analysis included 45 contrasts (32 SUDs and 13 BAs). The SUD subset was primarily driven by HUD (n = 10) and AUD (n = 13), alongside other substance categories (n = 9), while the BA subset comprised IGD (n = 10) and GD (n = 3).

In the pooled analysis, FWE-corrected convergence was observed in the right MCC, a midline region near the midbrain, the olfactory cortex, and the opercular inferior frontal gyrus. The SUD-only analysis retained supracallosal anterior cingulate and MCC clusters, whereas the BA-only analysis and the SUDs and BAs conjunction yielded no FWE-corrected clusters. Crucially, in the direct SUDs > BAs comparison, a left hippocampal cluster remained robust under this stringent restriction, whereas the primary ACCpre and PCUN differences did not survive. Regarding subtype analyses, multiple HUD-related clusters were retained, but no clusters survived in the IGD > AUD comparison or subtype conjunctions. Full results are detailed in Table S10.

#### 3.6.2 Analyses stratified by visual cue type

To evaluate the potential influence of stimulus modality, we stratified the analyses by visual cue type. The picture-cue analysis included 44 contrasts (33 SUDs and 11 BAs), while the video-cue analysis included 9 contrasts. Due to the limited sample size, the video-cue analysis was restricted to the pooled addiction group, identifying FWE-corrected clusters in the right medial orbitofrontal cortex and medial superior frontal gyrus.

For the picture-cue modality, the pooled analysis identified significant clusters in the MCC, supracallosal anterior cingulate, and midbrain (PAG vicinity). Notably, the direct SUDs > BAs comparison restricted to picture cues successfully retained a left precuneus cluster. Subtype analyses within this modality yielded multiple HUD-related clusters, as well as significant HUD > AUD (n = 12 vs. 12) and HUD > IGD (n = 12 vs. 9) differences. In contrast, no FWE-corrected clusters were found in the AUD-only, IGD-only, or IGD > AUD picture-cue analyses. Because the video subset was underpowered for cross-diagnostic evaluation, no formal statistical comparison between picture and video modalities was conducted; therefore, the differing spatial maps between these modalities should be interpreted descriptively (Table S11).

#### 3.6.3 Exploratory analyses of methadone maintenance status in HUD

To explore the potential influence of concurrent pharmacological interventions, the HUD dataset was stratified into non-MMT (7 contrasts) and MMT (6 contrasts) subgroups. Independent pooling of these subsets revealed divergent FWE-corrected activation profiles: the non-MMT group recruited the left ACCpre and MCC, whereas the MMT group engaged the left cerebellar Crus I and right ORBinf. A direct comparison between these subgroups demonstrated greater engagement of the right SFGmed in non-MMT individuals, contrasting with heightened left Crus I activity in the MMT group. However, owing to the small, non-randomized composition of these subsets, these differences preclude causal interpretations regarding treatment effects. Rather, these findings highlight treatment status as a critical potential confounder and a notable source of variation in HUD cue reactivity, underscoring the strictly exploratory nature of this sub-analysis (Table S12).

### 3.7 Heterogeneity and publication bias

Among the 28 FWE-corrected peaks in the primary category and subtype analyses (Tables S6-S7), heterogeneity was low for 20, moderate for 6, and substantial for 2. Of the 14 FWE-corrected clinical meta-regression peaks (Table S8), 13 had low heterogeneity. Across 80 FWE-corrected peaks in the demographic and other sensitivity analyses (Tables S9-S12), heterogeneity was low for 54, moderate for 12, substantial for 13, and considerable for one. No available Egger’s test for these peaks met the prespecified *p* < 0.1 threshold; however, the small number of contrasts in several analyses limits the power of this test.

## 4. Discussion

The present meta-analysis compared SUDs and BAs within a visual cue-reactivity framework. This approach complements and extends recent authoritative syntheses, such as the extensive SUD-focused meta-analysis by Roberge et al. (28), by introducing a direct cross-diagnostic comparison to determine whether behavioral addictions recruit the same neural circuitry as substance use disorders (28–31). The pooled analysis identified cue-related effects in the MCC, PreCG, and midbrain (PAG vicinity). Direct comparison identified greater effects in SUDs than in BAs in the left hippocampus, ACCpre, and PCUN. However, the SUDs ∩ BAs conjunction did not yield an FWE-corrected cluster. These findings refine hypotheses about cross-diagnostic cue processing and category-related differences.

Consistent with previous meta-analytic syntheses of addiction cue-reactivity (57–59), our pooled analysis identified significant convergence in the MCC and PreCG. This cross-diagnostic convergence suggests that these regions support generalized processes during cue exposure, such as motivational evaluation and action-preparation or sensorimotor processing, irrespective of the specific addiction category. However, because the pooled dataset contained substantially more SUD than BA contrasts and the BA-specific analysis yielded no FWE-corrected clusters, these effects do not establish equal contributions from both categories or a shared cross-diagnostic signature. Furthermore, although previous human meta-analyses have frequently identified midbrain and ventral tegmental area (VTA) responses to drug cues, particularly for illicit substances (59), our coordinate-based approach did not detect stable midbrain convergence. Nevertheless, animal models provide a robust circuit-level context for our cortical findings: cue-induced opioid seeking in rodents relies on a distributed network encompassing the cingulate cortex, amygdala, and striato-midbrain structures (60), with specific anterior cingulate-to-VTA projections actively driving drug-context seeking (61). Collectively, these translational findings support the plausibility of cingulate and affective-motivational circuitry in cue responding as core components of a broader, well-conserved cue-reactivity network.

Among the direct category differences, both the left hippocampal and precuneus effects persisted after study-level age adjustment. The hippocampal effect also survived restriction to addiction cues versus control cues, whereas the precuneus effect survived restriction to picture cues; neither was reproduced in the male-only analysis. The ACCpre finding showed less stable localization. Previous studies have reported medial-temporal, precuneus, and anterior-cingulate cue responses in both substance and behavioral addictions (58, 62–64). Neurosynth associations with autobiographical-memory terms for the ACCpre and PCUN offer one hypothesis for interpreting these findings, whereas the hippocampal decoding was broader and included emotional- and face-related terms (Table S6). Our findings therefore concern differences in the relative magnitude and spatial distribution of case-control effects in this dataset, not regions exclusive to SUDs or evidence of a pharmacologically caused mechanism. Taken together, the direct comparison motivates further investigation of memory- and self-related cue processing.

Subtype comparisons showed similarly uneven stability. The right orbital inferior frontal gyrus and pallidum effects in HUD > AUD persisted across the primary, age-adjusted, contrast-restricted, and picture-restricted analyses, making them candidates for further direct testing. Prior opioid cue-reactivity findings provide contextual support but do not independently replicate an HUD-AUD difference (64). By contrast, no HUD > IGD cluster survived age adjustment, and the IGD-AUD pattern changed substantially in the adjusted model. Nor does the absence of FWE-corrected AUD-only or IGD-only clusters imply an absence of cue reactivity in those disorders; an earlier AUD synthesis, for example, identified medial prefrontal and cingulate effects (65). However, given the strong association between age and diagnosis, these less stable contrasts should not be presented as established subtype signatures.

Our contrast-restricted sensitivity analysis directly addresses a critical source of methodological variance. Specifically, contrasts comparing addiction cues to neutral cues, addiction cues alone, and addiction cues to a low-level baseline do not necessarily capture identical neural processes. By restricting our dataset exclusively to strictly controlled ‘addiction cue versus control cue’ designs, the pooled MCC and midline findings, as well as the left hippocampal difference in the SUDs > BAs comparison, remained robust. Conversely, the primary between-category differences in the ACCpre and PCUN did not survive this rigorous restriction. We therefore caution that the ACCpre and PCUN findings may be analysis-sensitive. This attenuation could reflect the successful elimination of contrast-definition heterogeneity, or it may simply result from reduced statistical power following the exclusion of contrasts; the present dataset cannot definitively disentangle these competing explanations. Similarly, stratifying by stimulus modality revealed differing spatial maps between picture- and video-cue paradigms. However, because the video-cue subset was heavily underpowered (comprising only nine contrasts), no formal cross-modality comparison was feasible, and these modality-specific differences remain descriptive(29, 35, 66).

Clinical moderator findings were exploratory. Withdrawal duration showed opposite study-level association slopes in nearby left medial superior frontal regions in HUD and AUD, with each analysis based on seven contrasts. Although the peaks were not identical, this pattern raises the possibility that the relationship between abstinence and cue-related neural responses may vary across SUDs. This interpretation is clinically relevant in light of prior heroin fMRI evidence linking longer abstinence to lower cue-related responses in medial prefrontal and inferior parietal regions (67), and AUD evidence that abstinence duration is associated with prefrontal-striatal responses to alcohol cues and subsequent drinking outcomes (68). The MMT-stratified HUD analysis likewise raises treatment status as a possible source of variation. Prior studies have reported differences in cue responses across treatment contexts (39), and acute reductions in some responses after methadone dosing in the same patients (69). Together, these studies and the present analysis highlight treatment status and dose timing as clinically relevant factors to document in cue-reactivity research.

The main contribution of this study is a direct comparison of SUDs and BAs within a relatively consistent visual cue-reactivity framework. The left hippocampal difference was retained in the age-adjusted and cue-versus-control analyses, whereas the precuneus finding was retained after age adjustment and in the picture-only analysis; neither pattern persisted across all sensitivity analyses. Taken together, the findings motivate two candidate dimensions for future investigation: memory- and self-related cue processing, suggested by the direct SUD–BA differences in the hippocampus, ACCpre, and PCUN, and control- or action-related processes, suggested by the pooled MCC and PreCG findings. These dimensions are an interpretive framework rather than established or separable mechanisms, and their evidence comes from different levels of analysis. No FWE-corrected conjunction cluster was identified; thus, a shared neural core was not established. The sensitivity of some findings to age adjustment and analytic restrictions also argues against treating them as diagnostic signatures. Prospective evidence linking cue and craving measures to subsequent substance use and relapse (70), together with meta-analytic evidence relating cue-elicited brain activity to SUD treatment outcomes (71), makes these candidate patterns relevant for future clinical research. Such work may clarify which cue-related processes generalize across diagnostic categories and which require disorder-specific approaches. For instance, the greater precuneus involvement in SUDs suggests a stronger emotional and episodic memory component during cue exposure, highlighting these regions as potential targets for substance-specific interventions (e.g., memory retrieval-extinction procedures), whereas shared prefrontal targets might benefit transdiagnostic neuromodulation strategies.

Several limitations warrant consideration. First, the behavioral addiction (BA) dataset was relatively small (16 contrasts: 12 IGD, 4 GD) compared to the SUD group. This sample imbalance restricts the statistical power of BA-only and conjunction analyses. Consequently, the lack of FWE-corrected shared clusters indicates an absence of robust evidence under current statistical thresholds, rather than definitive proof that shared neural mechanisms are entirely absent. Second, demographic profiles were highly collinear with diagnostic categories; for instance, SUD cohorts were substantially older on average than BA cohorts. Because study-level meta-regression cannot fully disentangle such collinear effects, disorder-specific differences, especially those sensitive to age adjustment, must be interpreted cautiously. Furthermore, the source literature is heavily male-skewed. The scarcity of female-inclusive studies means sex-related confounding cannot be definitively excluded, limiting the generalizability of these neural signatures to women. Third, although the meta-analysis was strictly restricted to visual cues to reduce sensory variance, paradigm designs, including specific stimulus content, control conditions, and fMRI contrast definitions, remained heterogeneous across primary studies, which may introduce methodological noise. Fourth, unmeasured clinical variables remain a potential confounder. While exploratory analyses revealed divergent activation profiles between MMT and non-MMT subgroups, pervasive inconsistent reporting of concurrent medications, psychotherapy, abstinence duration, and withdrawal states precluded comprehensive moderator analyses. The pooled neuroimaging signals must therefore be interpreted with this clinical heterogeneity in mind. Finally, the current findings should not be generalized to non-visual sensory domains, food-related conditions, or other proposed behavioral addictions. Such conditions currently lack a formalized diagnostic consensus or present a hybrid clinical profile that obscures the distinct boundary between substance ingestion and behavioral engagement.

In conclusion, visual cue-reactivity studies motivated further investigation of memory- and self-related cue processing, suggested by the direct category differences, and control- or action-related processes, suggested by pooled findings. The regional patterns varied across sensitivity analyses, and no FWE-corrected conjunction cluster was identified; therefore, a shared neural core or diagnostic signature was not established. Larger, more demographically balanced longitudinal studies with standardized cue contrasts and detailed treatment information are needed to test whether these candidate patterns generalize across disorders and have predictive or treatment-related value.

## Data Availability

Data will be made available on request.

## Acknowledgments

This work was supported by the STI2030-Major Projects (2022ZD021120) and the National Natural Science Foundation of China (82471515). Data will be made available on request.

Y.S. and Q.H.Z. designed the study. Q.H.Z., Z.F.W., J.P., and Y.C.H. performed literature screening and data extraction. Q.H.Z. analyzed the data and drafted the manuscript, with methodological support from X.Q.Y., T.Y.W., X.L., and T.Y.J. Y.X.S. contributed to result interpretation. J.S., A.M.S.W., T.Y.J. and Y.S. supervised the project. All authors critically reviewed and approved the final manuscript.

