## Supplementary Materials for "Convergent and Divergent Neural Signatures of Cue-Induced Response in Substance and Behavioral Addictions: A Coordinate-Based Neuroimaging Meta-Analysis"

**Supplementary material**

**Contents**

Table S1: Detailed search terms for PubMed	2

Table S2: Study characteristics and clinical indexes extracted from studies included	4

Table S3: Detailed demographic characteristics of the patient and control groups in included studies	12

Table S4: Weighted linear regression analyses of age and gender differences across addiction subgroups	14

Table S5: Quality assessment scores of included studies based on a 12‑item neuroimaging checklist	15

Table S6: Meta-analytic results of cue-reactivity related neural activation for addiction categories	17

Table S7: Subgroup‑specific and comparative neural activation patterns for addiction categories	19

Table S8: Brain–behavior correlations between clinical measures and cue‑reactivity activation	22

Table S9: Demographic sensitivity analyses for subgroup comparisons	24

Table S10: Sensitivity analyses in the contrast “addiction cues vs. control cues”	26

Table S11: Sensitivity analyses in different cue types	30

Table S12: Sensitivity analyses for methadone maintenance treatment in HUD	34

Figure S1: Activation patterns in BAs and shared neural core across addictions	36

Figure S2: Subtype-specific and shared neural activation patterns across specific addiction subtypes.	37

Figure S3: Exploratory correlation between brain activation and other clinical measures	38

Figure S4: Sensitivity analysis restricted to “addiction cue vs. control cue” contrasts	39

Figure S5: Sensitivity analyses stratified by visual cue type	41

Figure S6: Age-adjusted diagnostic comparisons and male-only sensitivity analyses	44

Figure S7: Exploratory HUD analyses stratified by methadone maintenance treatment	45

Figure S8: Funnel plots for FWE-corrected peaks in Table S6	46

Figure S9: Funnel plots for FWE-corrected peaks in Table S7	47

Figure S10: Funnel plots for FWE-corrected peaks in Table S8	48

References	49

**Table S1. Detailed search terms for PubMed**

| **Category** | **Number** | **Search Term** |
| --- | --- | --- |
| Addiction | #1 | (Substance-Related Disorders[MeSH Terms]) OR (Alcohol-Related Disorders[MeSH Terms]) OR (Alcoholic Intoxication[MeSH Terms]) OR (Alcoholism[MeSH Terms]) OR (Binge Drinking[MeSH Terms]) OR (Amphetamine-Related Disorders[MeSH Terms]) OR (Cocaine-Related Disorders[MeSH Terms]) OR (Drug Overdose[MeSH Terms]) OR (Opiate Overdose[MeSH Terms]) OR (Inhalant Abuse[MeSH Terms]) OR (Marijuana Abuse[MeSH Terms]) OR (Narcotic-Related Disorders[MeSH Terms]) OR (Neonatal Abstinence Syndrome[MeSH Terms]) OR (Phencyclidine Abuse[MeSH Terms]) OR (Psychoses, Substance-Induced[MeSH Terms]) OR (Substance Abuse, Intravenous[MeSH Terms]) OR (Substance Abuse, Oral[MeSH Terms]) OR (Substance Withdrawal Syndrome[MeSH Terms]) OR (Alcohol Withdrawal Delirium[MeSH Terms]) OR (Alcohol Withdrawal Seizures[MeSH Terms]) OR (Tobacco Use Disorder[MeSH Terms]) |
|  | #2 | (Substance-Related Disorders*[Title/Abstract]) OR (Alcohol-Related Disorders*[Title/Abstract]) OR (Alcoholic Intoxication*[Title/Abstract]) OR (Alcoholism*[Title/Abstract]) OR (Binge Drinking*[Title/Abstract]) OR (Amphetamine-Related Disorders*[Title/Abstract]) OR (Cocaine-Related Disorders*[Title/Abstract]) OR (Drug Overdose*[Title/Abstract]) OR (Opiate Overdose*[Title/Abstract]) OR (Inhalant Abuse*[Title/Abstract]) OR (Marijuana Abuse*[Title/Abstract]) OR (Narcotic-Related Disorders*[Title/Abstract]) OR (Opioid-Induced Constipation*[Title/Abstract]) OR (Neonatal Abstinence Syndrome*[Title/Abstract]) OR (Phencyclidine Abuse*[Title/Abstract]) OR (Psychoses, Substance-Induced*[Title/Abstract]) OR (Substance Abuse, Intravenous*[Title/Abstract]) OR (Substance Abuse, Oral*[Title/Abstract]) OR (Substance Withdrawal Syndrome*[Title/Abstract]) OR (Alcohol Withdrawal Delirium*[Title/Abstract]) OR (Alcohol Withdrawal Seizures*[Title/Abstract]) OR (Tobacco Use Disorder*[Title/Abstract]) OR (substance dependence[Title/Abstract]) OR (substance abuse[Title/Abstract]) OR (substance use disorder[Title/Abstract]) OR (substance addiction[Title/Abstract]) OR (craving[Title/Abstract]) OR (alcohol dependence[Title/Abstract]) OR (alcohol abuse[Title/Abstract]) OR (alcoholism[Title/Abstract]) OR (alcoholic[Title/Abstract]) OR (alcohol[Title/Abstract]) OR (smoking[Title/Abstract]) OR (cigarette[Title/Abstract]) OR (nicotine[Title/Abstract]) OR (tobacco[Title/Abstract]) OR (psychostimulant drug[Title/Abstract]) OR (psychoactive substances[Title/Abstract]) OR (psychoactive drug[Title/Abstract]) OR (marijuana[Title/Abstract]) OR (cannabis[Title/Abstract]) OR (cannabinoids[Title/Abstract]) OR (hashish[Title/Abstract]) OR (marihuana[Title/Abstract]) OR (methamphetamine[Title/Abstract]) OR (heroin[Title/Abstract]) OR (opioid[Title/Abstract]) OR (morphine[Title/Abstract]) OR (amphetamine[Title/Abstract]) OR (methcathinone[Title/Abstract]) OR (synthetic cathinones[Title/Abstract]) OR(hallucinogens [Title/Abstract]) OR (methylenedioxymethamphetamine[Title/Abstract]) OR (MDMA[Title/Abstract]) OR (volatile Inhalants[Title/Abstract]) OR (ketamine[Title/Abstract]) OR (phencyclidine[Title/Abstract]) |
|  | #3 | #1 OR #2 |
|  | #4 | (Behaviour, Addictive[MeSH Terms]) OR (Technology Addiction[MeSH Terms]) OR (Internet Addiction Disorder[MeSH Terms]) OR (Gambling[MeSH Terms]) |
|  | #5 | (Behaviour, Addictive*[Title/Abstract]) OR (Technology Addiction*[Title/Abstract]) OR (Internet Addiction Disorder*[Title/Abstract]) OR (behavioural addiction[Title/Abstract]) OR (gaming addiction[Title/Abstract]) OR (gaming disorder[Title/Abstract]) OR (pathological video gamers[Title/Abstract]) OR (pathological gaming[Title/Abstract]) OR (Internet game disorder[Title/Abstract]) OR (online game disorder[Title/Abstract]) OR (cyberspace game disorder[Title/Abstract]) OR (computer game disorder[Title/Abstract]) OR (video game disorder[Title/Abstract]) OR (Internet game addiction[Title/Abstract]) OR (video game addiction[Title/Abstract]) OR (Internet gaming disorder[Title/Abstract]) OR (Internet addiction[Title/Abstract]) OR (pathological Internet use[Title/Abstract]) OR (Internet-use disorder[Title/Abstract]) OR (Internet gambling disorder[Title/Abstract]) OR (smartphone addiction[Title/Abstract]) OR (gambling addiction[Title/Abstract]) OR (gambling disorder[Title/Abstract]) OR (pathological gambling[Title/Abstract]) OR (pathological gaming[Title/Abstract]) OR (risk-taking behaviour[Title/Abstract]) |
|  | #6 | #4 OR #5 |
|  | #7 | #3 OR #6 |
| Cue-reactivity | #8 | (Cues [MeSH Terms]) |
|  | #9 | (“cue-reactivity*”[All Fields]) OR (conditioned withdrawal [Title/Abstract]) OR (conditioned withdrawal*[Title/Abstract]) OR (“cue reactivity”[All Fields]) OR (cue exposure[Title/Abstract]) OR (cue exposure*[Title/Abstract]) OR (Cues*[Title/Abstract]) OR (stimuli*[Title/Abstract]) OR (visual cue*[Title/Abstract]) OR (picture cue*[Title/Abstract]) OR (video cue*[Title/Abstract]) |
|  | #10 | #7 OR #8 |
| Neuroimaging | #11 | (Magnetic Resonance Imaging[MeSH Terms]) |
|  | #12 | (Magnetic Resonance imaging*[Title/Abstract]) OR (functional Magnetic Resonance Imaging[Title/Abstract]) OR (functional MRI[Title/Abstract]) OR (fMRI[Title/Abstract]) OR (MRI[Title/Abstract]) OR (neuroimaging[Title/Abstract]) OR (brain Imaging[Title/Abstract]) OR (BOLD[Title/Abstract]) OR (brain reactivity[Title/Abstract]) OR (neutral reactivity[Title/Abstract]) OR (voxel[Title/Abstract]) OR (blood-oxygen-level-dependent imaging[Title/Abstract]) |
|  | #13 | #10 OR #11 |
| Control | #14 | “control groups”[MeSH Terms] OR “controlling”[All Fields] OR “controllability”[All Fields] OR “controllable”[All Fields] OR “controllably”[All Fields] OR “controller”[All Fields] OR “controllers”[All Fields]OR “controlling”[All Fields] OR “controls”[All Fields] OR “prevention and control”[MeSH Subheading] OR (“prevention”[All Fields] AND “control”[All Fields]) OR “prevention and control”[All Fields] OR “control”[All Fields] OR (“control”[All Fields] AND “groups”[All Fields]) OR “control groups”[All Fields] OR “healthy control” [All Fields] |
| Participant | #15 | “participant*”[All Fields] OR “subject*”[All Fields] OR “individual*”[All Fields] OR “people”[All Fields] OR “case”[All Fields] |
|  | #16 | #9 AND #12 AND #13 AND #14 AND #15 |

**Table S2. Study characteristics and clinical indexes extracted from studies included**

| **Study** | **Category** | **Diagnostic** | **Cue** | **Contrast** | **Duration of substance use (d)** | **Duration of withdrawal (d)** | **Duration of treatment (d)** | **Dosage of substance use (g/d)** | **Total substance does used (g)** | **Average methadone does used (mg/d)** | **Severity of AUD** | **Obsessive-compulsive drinking degree** | **Anxiety degree** | **Depression degree** | **Duration of internet gaming (h/w)** | **Severity of IGD** |
| --- | --- | --- | --- | --- | --- | --- | --- | --- | --- | --- | --- | --- | --- | --- | --- | --- |
| Li2013 ^1^ | HUD | DSM-IV | Picture | 1 | 2679 | 17.6 | NA | 0.6 | NA | NA | NA | NA | NA | NA | NA | NA |
| Zeng2018 ^2^ | HUD | DSM-IV | Picture | 1 | NA | 42.11 | NA | 0.46 | NA | NA | NA | NA | 0.42 | NA | NA | NA |
| Huang2024 ^3^ | HUD | DSM-5 | Picture | 1 | 4007.7 | 196.78 | 184.77 | NA | NA | NA | NA | NA | NA | 1.01 | NA | NA |
| MacNiven2018 ^4^ | StiUD | DSM-5 | Picture | 1 | NA | NA | NA | NA | NA | NA | NA | NA | NA | 1.00 | NA | NA |
| Blaine2020 ^5^ | AUD | DSM-5 | Picture | 1 | NA | 5.5 | NA | NA | NA | NA | NA | NA | NA | NA | NA | NA |
| Hermann2006 ^6^ | AUD | ICD-10 | Picture | 1 | NA | 15 | NA | NA | NA | NA | 2.22 | NA | NA | 0.38 | NA | NA |
| Kim2014 ^7^ | AUD | DSM-IV | Video | 1 | NA | NA | NA | NA | NA | NA | 2.64 | 2.27 | NA | 0.87 | NA | NA |
| Hong2017 ^8^ | NUD | DSM-IV | Picture | 1 | NA | NA | NA | NA | NA | NA | NA | NA | 0.36 | 0.21 | NA | NA |
| Li2012 ^9^ | HUD | DSM-IV | Picture | 1 | 2358 | 21.7 | NA | 1 | 2171.10 | NA | NA | NA | NA | NA | NA | NA |
| Huang2018 ^10^ | MUD | DSM-IV | Picture | 1 | 1798.8 | 555 | NA | NA | NA | NA | NA | NA | NA | NA | NA | NA |
| Zhou2019 ^11^ | CanUD | DSM-IV | Picture | 1 | NA | NA | NA | NA | NA | NA | NA | NA | 0.50 | NA | NA | NA |
| Sjoerds2014 ^12^ | AUD | DSM-IV | Picture | 1 | NA | 12.2 | NA | NA | NA | NA | 2.12 | NA | 1.19 | 1.97 | NA | NA |
| Grusser2004 ^13^ | AUD | ICD-10 | Picture | 1 | NA | NA | NA | NA | NA | NA | 5.21 | NA | NA | 0.35 | NA | NA |
| Liao2018 ^14^ | KUD | DSM-IV | Video | 3 | 1251 | NA | NA | 1.4245 | NA | NA | NA | NA | NA | NA | NA | NA |
| Dakhili2022 ^15^ | MUD | DSM-IV | Picture | 1 | NA | 81.6 | NA | NA | NA | NA | NA | NA | NA | NA | NA | NA |
| Wei2021PA ^16^ | HUD | DSM-5 | Picture | 1 | 2364 | 354 | NA | 0.8 | 1880.20 | NA | NA | NA | NA | NA | NA | NA |
| Wei2021MMT ^16^ | HUD | DSM-5 | Picture | 1 | 1734 | NA | 345 | 0.4 | 510.90 | 43.60 | NA | NA | NA | NA | NA | NA |
| Fryer2013 ^17^ | AUD | DSM-IV | Picture | 1 | NA | NA | NA | NA | NA | NA | NA | NA | NA | 0.84 | NA | NA |
| Yang2009 ^18^ | HUD | DSM-IV | Video | 1 | 1679 | NA | NA | NA | NA | NA | NA | NA | NA | NA | NA | NA |
| Wrase2007 ^19^ | AUD | ICD-10 | Picture | 1 | NA | NA | NA | NA | NA | NA | NA | 3.67 | NA | 0.76 | NA | NA |
| Kong2020 ^20^ | BQD | DSM-5 | Picture | 1 | 5558.95 | NA | NA | 40.19 | NA | NA | NA | NA | NA | NA | NA | NA |
| Beck2012 ^21^ | AUD | ICD-10 | Picture | 1 | NA | 14.96 | NA | NA | NA | NA | 4.03 | NA | NA | 0.54 | NA | NA |
| Beck2018 ^22^ | AUD | ICD-10 | Picture | 1 | NA | 12.35 | NA | NA | NA | NA | NA | NA | -0.13 | 1.17 | NA | NA |
| Bach2020 ^23^ | AUD | DSM-IV | Picture | 1 | NA | NA | NA | NA | NA | NA | 2.26 | 2.84 | 1.12 | 1.34 | NA | NA |
| Bell2014 ^24^ | CoUD | DSM-IV | Picture | 1 | NA | NA | NA | NA | NA | NA | NA | NA | NA | NA | NA | NA |
| Lee2013 ^25^ | AUD | DSM-IV | Picture | 1 | NA | NA | NA | NA | NA | NA | 4.46 | 20.56 | 1.14 | 1.79 | NA | NA |
| Tapert2003 ^26^ | AUD | DSM-IV | Picture | 1 | NA | NA | NA | NA | NA | NA | NA | NA | 1.21 | 0.73 | NA | NA |
| Zijlstra2009 ^27^ | HUD | DSM-IV | Picture | 3 | 5840 | 56.7 | NA | NA | NA | NA | NA | NA | NA | NA | NA | NA |
| Lorenzetti2025 ^28^ | CanUD | DSM-5 | Picture | 1 | 2883.5 | 0.870833333 | NA | NA | NA | NA | NA | NA | 0.35 | 0.61 | NA | NA |
| Tabatabaei2014PA ^29^ | HUD | DSM-IV | Picture | 2 | 4142.75 | 276 | NA | NA | NA | NA | NA | NA | NA | NA | NA | NA |
| Tabatabaei2014MMT ^29^ | HUD | DSM-IV | Picture | 2 | 4033.25 | NA | 348.3 | NA | NA | 84.75 | NA | NA | NA | NA | NA | NA |
| Li2019 ^30^ | HUD | DSM-IV | Picture | 1 | NA | NA | 591 | 0.5 | 1284.10 | 42.70 | NA | NA | 0.80 | 0.81 | NA | NA |
| Li2015 ^31^ | HUD | DSM-IV | Picture | 1 | 2406.9 | NA | 652.2 | 0.55 | 1140.67 | 41.20 | NA | NA | 0.66 | 0.80 | NA | NA |
| Wang2014 ^32^ | HUD | DSM-IV | Picture | 1 | 1473.6 | NA | 413.1 | 0.5 | 609.95 | 42.50 | NA | NA | NA | NA | NA | NA |
| Vollstadt2011 ^33^ | NUD | DSM-IV | Picture | 1 | 5329 | NA | NA | NA | NA | NA | NA | NA | NA | NA | NA | NA |
| Kober2016CoUD ^34^ | CoUD | DSM-IV | Video | 3 | NA | NA | NA | NA | NA | NA | NA | NA | NA | NA | NA | NA |
| Sawyer2026^35^ | AUD | DSM-IV | Picture | 1 | 6460.5 | NA | NA | NA | NA | NA | NA | NA | NA | NA | NA | NA |
| Liu2017 ^36^ | IGD | CIAS | Picture | 1 | NA | NA | NA | NA | NA | NA | NA | NA | 0.41 | 0.90 | 18.92 | 3.22 |
| Wang2017 ^37^ | IGD | DSM-5 | Picture | 1 | NA | NA | NA | NA | NA | NA | NA | NA | NA | 0.21 | 18.90 | 2.18 |
| Zhang2020 ^38^ | IGD | DSM-5 | Picture | 1 | NA | NA | NA | NA | NA | NA | NA | NA | NA | NA | 18.18 | 1.46 |
| Goudriaan2010PRG ^39^ | GD | DSM-IV | Picture | 1 | NA | NA | NA | NA | NA | NA | NA | NA | NA | 0.79 | NA | NA |
| Ko2009 ^40^ | IGD | DCIA-C | Picture | 1 | NA | NA | NA | NA | NA | NA | NA | NA | NA | NA | NA | 2.62 |
| Kober2016PG ^34^ | GD | DSM-IV | Picture | 3 | NA | NA | NA | NA | NA | NA | NA | NA | NA | NA | NA | NA |
| Ko2013 ^41^ | IGD | DCIA | Picture | 1 | NA | NA | NA | NA | NA | NA | NA | NA | NA | NA | NA | 7.51 |
| Sun2012 ^42^ | IGD | CIAS | Picture | 2 | NA | NA | NA | NA | NA | NA | NA | NA | NA | NA | 51.30 | 3.47 |
| Han2010 ^43^ | IGD | DSM-IV | Video | 1 | NA | NA | NA | NA | NA | NA | NA | NA | NA | -0.43 | 45.50 | 5.28 |
| Lorenz2013 ^44^ | IGD | ICD-10 | Picture | 1 | NA | NA | NA | NA | NA | NA | NA | NA | 0.67 | 1.18 | NA | NA |
| Crockford2005 ^45^ | GD | DSM-IV | Video | 1 | NA | NA | NA | NA | NA | NA | NA | NA | NA | NA | NA | NA |
| Han2012 ^46^ | IGD | YIAS | Picture | 1 | NA | NA | NA | NA | NA | NA | NA | NA | NA | NA | 34.50 | 3.79 |
| Zhang2016 ^47^ | IGD | CIAS | Video | 1 | NA | NA | NA | NA | NA | NA | NA | NA | 0.64 | 1.25 | 27.26 | 4.36 |
| Liu2016 ^48^ | IGD | YDQ | Video | 2 | NA | NA | NA | NA | NA | NA | NA | NA | 14.24 | 6.92 | NA | NA |
| Zhou2021 ^49^ | IGD | DSM-5 | Picture | 1 | NA | NA | NA | NA | NA | NA | NA | NA | NA | NA | NA | 2.73 |
| Limbrick2017 ^50^ | GD | DSM-IV | Picture | 1 | NA | NA | NA | NA | NA | NA | NA | NA | NA | NA | NA | NA |

NA indicates that the variable was not reported or not applicable to the study design. Continuous clinical measures are presented as standardized mean values where available. Contrast: 1 means cue vs. neutral/control, 2 means only cue condition, 3 means cue vs. baseline; DSM-IV: Diagnostic and Statistical Manual of Mental Disorders, Fourth Edition; DSM-5: Diagnostic and Statistical Manual of Mental Disorders, Fifth Edition: ICD-10, International Classification of Diseases, Tenth Revision; CIAS: Chen Internet Addiction Scale; DCIA-C: Diagnostic Criteria of Internet Addiction for College Students; YIAS: Young’s Internet Addiction Scale; YDQ: Young Diagnostic Questionnaire; AUD: Alcohol Use Disorder; HUD: Heroin Use Disorder; NUD: Nicotine Use Disorder; MUD: Methamphetamine Use Disorder; CanUD: Cannabis Use Disorder; BQD: Betel-Quid Dependence; CoUD: Cocaine Use Disorder; KUD: Ketamine Use Disorder; StiUD: Stimulant Use Disorder; GD: Gambling Disorder; IGD: Internet Gaming Disorder.

**Table S3. Detailed demographic characteristics of the patient and control groups in included studies**

| **Study** | **t_thr** |  | **n_p** | **male_p** | **female_p** | **mean_p** | **sd_p** | **n_c** | **male_c** | **female_c** | **mean_c** | **sd_c** | **total age** | **total age_sd** | **gender_ratio** |
| --- | --- | --- | --- | --- | --- | --- | --- | --- | --- | --- | --- | --- | --- | --- | --- |
| Li2013 ^1^ | 3.42 |  | 14 | 14 | 0 | 35.40 | 6.40 | 15 | 15 | 0 | 31.90 | 6.80 | 33.65 | 6.72 | 1.00 |
| Zeng2018 ^2^ | 3.22 |  | 37 | 24 | 13 | 41.79 | 2.36 | 29 | 19 | 10 | 44.01 | 4.87 | 42.77 | 3.81 | 0.65 |
| Huang2024 ^3^ | 3.27 |  | 32 | 25 | 7 | 40.25 | 8.82 | 21 | 13 | 8 | 40.58 | 10.84 | 40.38 | 9.57 | 0.72 |
| MacNiven2018 ^4^ | 3.20 |  | 36 | 34 | 2 | 43.30 | 13.30 | 40 | 24 | 16 | 32.00 | 11.60 | 37.35 | 13.59 | 0.76 |
| Blaine2020 ^5^ | 3.19 |  | 44 | 28 | 16 | 33.00 | 11.00 | 43 | 23 | 20 | 32.00 | 10.00 | 32.51 | 10.47 | 0.59 |
| Hermann2006 ^6^ | 3.61 |  | 10 | 10 | 0 | 40.00 | 7.00 | 10 | 10 | 0 | 38.00 | 5.00 | 39.00 | 6.01 | 1.00 |
| Kim2014 ^7^ | 3.23 |  | 38 | 27 | 11 | 41.60 | 7.10 | 26 | 20 | 6 | 43.90 | 8.60 | 42.53 | 7.76 | 0.73 |
| Hong2017 ^8^ | 3.41 |  | 15 | 15 | 0 | 39.90 | 4.90 | 15 | 15 | 0 | 39.20 | 5.20 | 39.55 | 4.98 | 1.00 |
| Li2012 ^9^ | 3.30 |  | 24 | 24 | 0 | 32.80 | 6.60 | 20 | 20 | 0 | 35.00 | 7.00 | 33.80 | 6.80 | 1.00 |
| Huang2018 ^10^ | 3.25 |  | 28 | 28 | 0 | 31.68 | 7.06 | 27 | 27 | 0 | 33.93 | 7.21 | 32.78 | 7.16 | 1.00 |
| Zhou2019 ^11^ | 3.23 |  | 18 | 18 | 0 | 22.94 | 2.71 | 44 | 44 | 0 | 23.20 | 4.32 | 23.12 | 3.90 | 1.00 |
| Sjoerds2014 ^12^ | 2.70 |  | 30 | 16 | 14 | 46.50 | 8.50 | 15 | 11 | 4 | 46.80 | 10.00 | 46.60 | 8.91 | 0.60 |
| Grusser2004 ^13^ | 3.85 |  | 5 | NA | NA | NA | NA | 10 | 5 | 5 | 36.00 | 11.00 | NA | NA | NA |
| Liao2018 ^14^ | 3.16 |  | 40 | 32 | 8 | 26.80 | 4.93 | 89 | 71 | 18 | 27.10 | 5.72 | 27.01 | 5.47 | 0.80 |
| Dakhili2022 ^15^ | 3.21 |  | 53 | 53 | 0 | 32.12 | 5.89 | 23 | 23 | 0 | 31.27 | 5.69 | 31.86 | 5.81 | 1.00 |
| Wei2021PA ^16^ | 3.54 |  | 24 | 24 | 0 | 33.00 | 6.80 | 20 | 20 | 0 | 35.20 | 7.00 | 34.00 | 6.90 | 1.00 |
| Wei2021MMT ^16^ | 3.54 |  | 21 | 21 | 0 | 35.10 | 9.30 | 20 | 20 | 0 | 35.20 | 7.00 | 35.15 | 8.16 | 1.00 |
| Fryer2013 ^17^ | 2.44 |  | 16 | 12 | 4 | 35.50 | 10.80 | 20 | 15 | 5 | 35.90 | 10.00 | 35.72 | 10.21 | 0.75 |
| Yang2009 ^18^ | 2.02 |  | 9 | 8 | 1 | 33.60 | 4.30 | 9 | 9 | 0 | 32.20 | 5.90 | 32.90 | 5.06 | 0.94 |
| Wrase2007 ^19^ | 3.39 |  | 16 | 16 | 0 | 42.38 | 7.52 | 16 | 16 | 0 | 39.94 | 8.59 | 41.16 | 8.04 | 1.00 |
| Kong2020 ^20^ | 3.23 |  | 48 | 48 | 0 | 34.85 | 8.10 | 22 | 22 | 0 | 32.05 | 6.25 | 33.97 | 7.64 | 1.00 |
| Beck2012 ^21^ | 3.20 |  | 30 | 19 | 11 | 39.80 | 6.33 | 46 | 30 | 16 | 39.37 | 7.72 | 39.54 | 7.16 | 0.64 |
| Beck2018 ^22^ | 3.29 |  | 23 | 16 | 7 | 46.17 | 6.15 | 23 | 16 | 7 | 43.35 | 10.05 | 44.76 | 8.36 | 0.70 |
| Bach2020 ^23^ | 3.19 |  | 50 | 50 | 0 | 45.60 | 8.90 | 35 | 35 | 0 | 42.00 | 9.80 | 44.12 | 9.39 | 1.00 |
| Bell2014 ^24^ | 3.33 |  | 20 | 20 | 0 | 47.80 | 8.50 | 19 | 19 | 0 | 42.20 | 12.10 | 45.07 | 10.65 | 1.00 |
| Lee2013 ^25^ | 2.70 |  | 17 | 12 | 5 | 34.70 | 4.90 | 25 | 18 | 7 | 34.00 | 5.40 | 34.28 | 5.15 | 0.71 |
| Tapert2003 ^26^ | 2.05 |  | 15 | 9 | 6 | 16.96 | 0.78 | 15 | 9 | 6 | 16.35 | 1.02 | 16.66 | 0.94 | 0.60 |
| Zijlstra2009 ^27^ | 3.42 |  | 12 | 12 | 0 | 44.50 | 3.90 | 17 | 17 | 0 | 40.00 | 10.10 | 41.86 | 8.33 | 1.00 |
| Lorenzetti2025 ^28^ | 3.12 |  | 65 | 46 | 19 | 27.00 | 7.90 | 43 | 27 | 16 | 27.80 | 9.40 | 27.32 | 8.50 | 0.68 |
| Tabatabaei2014PA ^29^ | 2.07 |  | 20 | 20 | 0 | 32.00 | 4.40 | 20 | 20 | 0 | 30.80 | 5.00 | 31.40 | 4.69 | 1.00 |
| Tabatabaei2014MMT ^29^ | 2.07 |  | 20 | 20 | 0 | 33.60 | 4.70 | 20 | 20 | 0 | 30.80 | 5.00 | 32.20 | 5.00 | 1.00 |
| Li2019 ^30^ | 3.27 |  | 31 | 31 | 0 | 35.40 | 7.30 | 20 | 20 | 0 | 35.20 | 7.00 | 35.32 | 7.11 | 1.00 |
| Li2015 ^31^ | 3.23 |  | 44 | 44 | 0 | 35.02 | 7.05 | 20 | 20 | 0 | 35.20 | 7.00 | 35.08 | 6.98 | 1.00 |
| Wang2014 ^32^ | 3.55 |  | 30 | 30 | 0 | 33.54 | 7.75 | 17 | 17 | 0 | 34.30 | 7.30 | 33.81 | 7.52 | 1.00 |
| Vollstadt2011 ^33^ | 2.70 |  | 22 | 22 | 0 | 31.00 | 7.00 | 21 | 21 | 0 | 29.00 | 5.00 | 30.02 | 6.12 | 1.00 |
| Kober2016CoUD ^34^ | 2.64 |  | 30 | 18 | 12 | 42.93 | 6.09 | 45 | 23 | 22 | 30.40 | 10.76 | 35.41 | 11.03 | 0.55 |
| Sawyer2026^35^ | 3.19 |  | 42 | 21 | 21 | 53.9 | 11.0 | 42 | 21 | 21 | 53.9 | 12.4 | 53.9 | 11.65 | 0.50 |
| Liu2017 ^36^ | 3.23 |  | 39 | 39 | 0 | 22.64 | 2.12 | 23 | 23 | 0 | 23.09 | 2.13 | 22.81 | 2.12 | 1.00 |
| Wang2017 ^37^ | 2.65 |  | 30 | NA | NA | 21.07 | 1.34 | 40 | NA | NA | 21.45 | 1.32 | 21.29 | 1.33 | NA |
| Zhang2020 ^38^ | 2.37 |  | 49 | 27 | 22 | 22.77 | 2.22 | 49 | 27 | 22 | 23.33 | 2.04 | 23.05 | 2.14 | 0.55 |
| Goudriaan2010PRG ^39^ | 3.37 |  | 17 | 17 | 0 | 35.30 | 9.40 | 17 | 17 | 0 | 34.70 | 9.70 | 35.00 | 9.41 | 1.00 |
| Ko2009 ^40^ | 3.92 |  | 10 | 10 | 0 | 22.00 | 1.49 | 10 | 10 | 0 | 22.70 | 1.34 | 22.35 | 1.43 | 1.00 |
| Kober2016PG ^34^ | 2.65 |  | 28 | 20 | 8 | 33.39 | 10.81 | 45 | 23 | 22 | 30.40 | 10.76 | 31.55 | 10.80 | 0.59 |
| Ko2013 ^41^ | 3.41 |  | 15 | 15 | 0 | 24.67 | 3.11 | 15 | 15 | 0 | 24.47 | 2.83 | 24.57 | 2.92 | 1.00 |
| Sun2012 ^42^ | 3.69 |  | 10 | 10 | 0 | 20.40 | 1.51 | 10 | 10 | 0 | 20.30 | 0.67 | 20.35 | 1.14 | 1.00 |
| Han2010 ^43^ | 2.95 |  | 11 | 11 | 0 | 21.50 | 5.60 | 8 | 8 | 0 | 20.30 | 4.10 | 20.99 | 4.93 | 1.00 |
| Lorenz2013 ^44^ | 3.61 |  | 8 | 8 | 0 | 25.00 | 7.40 | 9 | 9 | 0 | 24.80 | 6.90 | 24.89 | 6.91 | 1.00 |
| Crockford2005 ^45^ | 2.97 |  | 10 | 10 | 0 | 39.30 | 7.60 | 10 | 10 | 0 | 39.20 | 8.30 | 39.25 | 7.75 | 1.00 |
| Han2012 ^46^ | 3.24 |  | 15 | NA | NA | 14.20 | 1.50 | 15 | NA | NA | 14.00 | 1.30 | 14.10 | 1.38 | NA |
| Zhang2016 ^47^ | 1.69 |  | 40 | 40 | 0 | 21.95 | 1.84 | 19 | 19 | 0 | 22.89 | 2.23 | 22.25 | 2.00 | 1.00 |
| Liu2016 ^48^ | 3.30 |  | 19 | 11 | 8 | 21.40 | 1.00 | 19 | 11 | 8 | 20.80 | 1.10 | 21.10 | 1.08 | 0.58 |
| Zhou2021 ^49^ | 3.27 |  | 21 | 10 | 11 | 21.29 | 1.52 | 23 | 15 | 8 | 21.61 | 1.95 | 21.46 | 1.75 | 0.57 |
| Limbrick2017 ^50^ | 2.41 |  | 19 | 19 | 0 | 31.00 | NA | 19 | 19 | 0 | 28.00 | NA | 29.50 | NA | 1.00 |

Demographic data extraction from included studies. For each study, the table provides the following information for the patient/addicted group (p) and the control group (c) separately. t_thr: t‑threshold applied in the fMRI contrast; n_pat/n_con: number of participants; male_/female_: number of male and female participants; mean_: mean age in years; sd_: standard deviation of age; total_age: the combined total mean age; total_age_sd: the standard deviation of total mean age; gender_ratio: male ratio in the combined participants. NA otherwise indicates that the specific demographic variable was not reported in the original study.

**Table S4. Weighted linear regression analyses of age and gender differences across addiction subgroups**

| Variable | Comparison | Contrasts (n_1_/n_2_) | Diff | *t* value | *p* value | 95% CIs of Diff |
| --- | --- | --- | --- | --- | --- | --- |
| Total Age *(years)* | SUDs vs BAs | 36/16 | 11.37 | 5.18 | <0.001 | [6.96, 15.78] |
|  | HUD vs AUD | 13/12 | -4.72 | -1.73 | 0.097 | [-10.36, 0.93] |
|  | HUD vs IGD | 13/12 | 14.14 | 11.08 | < 0.001 | [11.50, 16.78] |
|  | AUD vs IGD | 12/12 | 18.86 | 6.90 | < 0.001 | [13.19, 24.52] |
| Gender ratio *(% male)* | SUDs vs BAs | 36/14 | 1.90 | 0.30 | 0.769 | [-11.05, 14.85] |
|  | HUD vs AUD | 13/12 | 22.05 | 3.51 | 0.002 | [9.04, 35.06] |
|  | HUD vs IGD | 13/10 | 12.51 | 1.64 | 0.116 | [-3.36, 28.39] |
|  | AUD vs IGD | 12/10 | -9.54 | -1.11 | 0.282 | [-27.54, 8.46] |

Differences were estimated using linear regression weighted by the total sample size of each contrast. A positive difference indicates a higher value in the first-listed group. Differences in male proportion are expressed in percentage points. The 95% CIs for differences were calculated from the regression estimates and t-statistics. The number of contrasts varies because demographic data were unavailable for some contrasts. SUDs: substance use disorders; BAs: behavioural addictions; HUD: heroin use disorder; AUD: alcohol use disorder; IGD: internet gaming disorder; CIs: confidence intervals.

**Table S5. Quality assessment scores of included studies based on a 12‑item neuroimaging checklist**

| **study** | **Q1** | **Q2** | **Q3** | **Q4** | **Q5** | **Q6** | **Q7** | **Q8** | **Q9** | **Q10** | **Q11** | **Q12** | **Total** |
| --- | --- | --- | --- | --- | --- | --- | --- | --- | --- | --- | --- | --- | --- |
| Li2013 ^1^ | 1 | 1 | 1 | 1 | 1 | 0 | 1 | 1 | 1 | 1 | 1 | 1 | 11 |
| Zeng2018 ^2^ | 1 | 1 | 1 | 1 | 1 | 1 | 1 | 1 | 1 | 1 | 1 | 1 | 12 |
| Huang2024 ^3^ | 1 | 1 | 1 | 1 | 1 | 1 | 1 | 1 | 1 | 1 | 1 | 1 | 12 |
| MacNiven2018 ^4^ | 1 | 1 | 1 | 1 | 1 | 1 | 1 | 1 | 1 | 1 | 1 | 1 | 12 |
| Blaine2020 ^5^ | 1 | 1 | 1 | 1 | 1 | 1 | 1 | 1 | 1 | 1 | 1 | 1 | 12 |
| Hermann2006 ^6^ | 1 | 1 | 1 | 0 | 0.5 | 0 | 1 | 1 | 1 | 1 | 1 | 1 | 9.5 |
| Kim2014 ^7^ | 1 | 1 | 1 | 1 | 0.5 | 0 | 1 | 1 | 1 | 1 | 1 | 1 | 10.5 |
| Hong2017 ^8^ | 1 | 1 | 1 | 1 | 0.5 | 0 | 1 | 1 | 1 | 1 | 1 | 1 | 10.5 |
| Li2012 ^9^ | 1 | 1 | 1 | 1 | 1 | 0 | 1 | 1 | 1 | 1 | 1 | 1 | 11 |
| Huang2018 ^10^ | 1 | 1 | 1 | 1 | 1 | 0 | 1 | 1 | 1 | 1 | 1 | 1 | 11 |
| Zhou2019 ^11^ | 1 | 1 | 1 | 1 | 1 | 1 | 1 | 1 | 1 | 1 | 1 | 1 | 12 |
| Sjoerds2014 ^12^ | 1 | 1 | 1 | 1 | 1 | 1 | 1 | 1 | 1 | 1 | 1 | 1 | 12 |
| Grusser2004 ^13^ | 1 | 1 | 1 | 0 | 0.5 | 0 | 1 | 1 | 1 | 1 | 1 | 1 | 9.5 |
| Liao2018 ^14^ | 1 | 1 | 1 | 1 | 1 | 1 | 1 | 1 | 1 | 1 | 1 | 1 | 12 |
| Dakhili2022 ^15^ | 1 | 1 | 1 | 1 | 1 | 1 | 1 | 1 | 1 | 1 | 1 | 1 | 12 |
| Wei2021PA ^16^ | 1 | 1 | 1 | 1 | 1 | 0 | 1 | 1 | 1 | 1 | 1 | 1 | 11 |
| Wei2021MMT ^16^ | 1 | 1 | 1 | 1 | 1 | 0 | 1 | 1 | 1 | 1 | 1 | 1 | 11 |
| Fryer2013 ^17^ | 1 | 1 | 1 | 1 | 1 | 0 | 1 | 1 | 1 | 1 | 1 | 1 | 11 |
| Yang2009 ^18^ | 1 | 1 | 1 | 0 | 0.5 | 0 | 1 | 1 | 1 | 1 | 1 | 1 | 9.5 |
| Wrase2007 ^19^ | 1 | 1 | 1 | 1 | 0.5 | 0 | 1 | 1 | 1 | 1 | 1 | 1 | 10.5 |
| Kong2020 ^20^ | 1 | 1 | 1 | 1 | 1 | 1 | 1 | 1 | 1 | 1 | 1 | 1 | 12 |
| Beck2012 ^21^ | 1 | 1 | 1 | 1 | 0.5 | 0 | 1 | 1 | 1 | 1 | 1 | 1 | 10.5 |
| Beck2018 ^22^ | 1 | 1 | 1 | 1 | 1 | 1 | 1 | 1 | 1 | 1 | 1 | 1 | 12 |
| Bach2020 ^23^ | 1 | 1 | 1 | 1 | 1 | 1 | 1 | 1 | 1 | 1 | 1 | 1 | 12 |
| Bell2014 ^24^ | 1 | 1 | 1 | 1 | 1 | 1 | 1 | 1 | 1 | 1 | 1 | 1 | 12 |
| Lee2013 ^25^ | 1 | 1 | 1 | 1 | 0.5 | 0 | 1 | 1 | 1 | 1 | 1 | 1 | 10.5 |
| Tapert2003 ^26^ | 1 | 1 | 1 | 1 | 0 | 0 | 1 | 1 | 1 | 1 | 1 | 1 | 10 |
| Zijlstra2009 ^27^ | 1 | 1 | 1 | 1 | 1 | 1 | 1 | 1 | 1 | 1 | 1 | 1 | 12 |
| Lorenzetti2025 ^28^ | 1 | 1 | 1 | 1 | 1 | 1 | 1 | 1 | 1 | 1 | 1 | 1 | 12 |
| Tabatabaei2014PA ^29^ | 1 | 1 | 1 | 1 | 0.5 | 1 | 1 | 1 | 1 | 1 | 1 | 1 | 11.5 |
| Tabatabaei2014MMT ^29^ | 1 | 1 | 1 | 1 | 0.5 | 1 | 1 | 1 | 1 | 1 | 1 | 1 | 11.5 |
| Li2019 ^30^ | 1 | 1 | 1 | 1 | 1 | 0 | 1 | 1 | 1 | 1 | 1 | 1 | 11 |
| Li2015 ^31^ | 1 | 1 | 1 | 1 | 1 | 0 | 1 | 1 | 1 | 1 | 1 | 1 | 11 |
| Wang2014 ^32^ | 1 | 1 | 1 | 1 | 1 | 0 | 1 | 1 | 1 | 1 | 1 | 1 | 11 |
| Vollstadt2011 ^33^ | 1 | 1 | 1 | 1 | 0.5 | 0 | 1 | 1 | 1 | 1 | 1 | 1 | 10.5 |
| Kober2016CoUD ^34^ | 1 | 1 | 1 | 1 | 1 | 0 | 1 | 1 | 1 | 1 | 1 | 1 | 11 |
| Sawyer2026^35^ | 1 | 1 | 1 | 1 | 1 | 0 | 1 | 1 | 1 | 1 | 1 | 1 | 11 |
| Liu2017 ^36^ | 1 | 1 | 1 | 1 | 1 | 1 | 1 | 1 | 1 | 1 | 1 | 1 | 12 |
| Wang2017 ^37^ | 1 | 1 | 1 | 1 | 1 | 1 | 1 | 1 | 1 | 1 | 1 | 1 | 12 |
| Zhang2020 ^38^ | 1 | 1 | 1 | 1 | 1 | 1 | 1 | 1 | 1 | 1 | 1 | 1 | 12 |
| Goudriaan2010PRG ^39^ | 1 | 1 | 1 | 1 | 1 | 1 | 1 | 1 | 1 | 1 | 1 | 1 | 12 |
| Ko2009 ^40^ | 1 | 1 | 1 | 0 | 1 | 1 | 1 | 1 | 1 | 1 | 1 | 1 | 11 |
| Kober2016PG ^34^ | 1 | 1 | 1 | 1 | 1 | 0 | 1 | 1 | 1 | 1 | 1 | 1 | 11 |
| Ko2013 ^41^ | 1 | 1 | 1 | 1 | 1 | 1 | 1 | 1 | 1 | 1 | 1 | 1 | 12 |
| Sun2012 ^42^ | 1 | 1 | 1 | 0 | 1 | 0 | 1 | 1 | 1 | 1 | 1 | 1 | 10 |
| Han2010 ^43^ | 1 | 1 | 1 | 0.5 | 0.5 | 0 | 1 | 1 | 1 | 1 | 1 | 1 | 10 |
| Lorenz2013 ^44^ | 1 | 1 | 1 | 0 | 1 | 0 | 1 | 1 | 1 | 1 | 1 | 1 | 10 |
| Crockford2005 ^45^ | 1 | 1 | 1 | 0 | 1 | 0 | 1 | 1 | 1 | 1 | 1 | 1 | 10 |
| Han2012 ^46^ | 1 | 1 | 1 | 1 | 1 | 1 | 0 | 1 | 1 | 1 | 1 | 1 | 11 |
| Zhang2016 ^47^ | 1 | 1 | 1 | 1 | 1 | 1 | 1 | 1 | 1 | 1 | 1 | 1 | 12 |
| Liu2016 ^48^ | 1 | 1 | 1 | 1 | 1 | 0 | 1 | 1 | 1 | 1 | 1 | 1 | 11 |
| Zhou2021 ^49^ | 1 | 1 | 1 | 1 | 1 | 1 | 1 | 1 | 1 | 1 | 1 | 1 | 12 |
| Limbrick2017 ^50^ | 1 | 1 | 1 | 1 | 1 | 1 | 1 | 1 | 1 | 1 | 1 | 1 | 12 |

Each study was evaluated using a checklist encompassing three domains: (1) Subject selection and characteristics (Q1: Patients were evaluated prospectively, specific diagnostic criteria were applied, and demographic data were reported.; Q2: Healthy comparison subjects were evaluated prospectively, psychiatric and medical illnesses were excluded; Q3: Important variables (e.g., age, gender, medication status, comorbidity, severity of illness) were checked, either by stratification or statistically; and Q4: Sample size per group > 10); (2) Methods for image acquisition and analysis (Q5: Magnet strength at least 1.5 T, scored as 0.5 for 1.5 T and 1 for 3 T; Q6: MRI slice-thickness ≤ 3 mm; Q7: Whole brain analysis was automated with no a priori regional selection; Q8: Coordinates are reported in a standard space; Q9: The imaging technique used was clearly described so that it could be reproduced; and Q10: Measurements were clearly described so that they could be reproduced); and (3) Results and conclusions (Q11: Statistical parameters for significant, and important non-significant, differences were provided; and Q12: Conclusions were consistent with the results obtained, and the limitations were discussed). Each item was scored as 0 (not met), 0.5 (partially met), or 1 (fully met), with the Total column representing the sum across all 12 items (maximum possible score = 12).

**Table S6. Meta-analytic results of cue-reactivity related neural activation for addiction categories**

| **Macroanatomical label** | | **MNI (x,y,z)** | | **SDM-Z** | | **Voxels** | | ***I2 (%)*** | | **Egger’s bias** | | **Egger’s p** | **Function decoding** |
| --- | --- | --- | --- | --- | --- | --- | --- | --- | --- | --- | --- | --- | --- |
| (a) SUDs ∪ BAs (n=53) | | | | | | | | | | | | | |
| Cingulate_Mid_R ^a^ | 4,-18,30 | | 5.63 | 9085 | 30.87 | | 0.88 | | 0.29 | | stop signal; response inhibition; stop; inhibitory; inhibition | | |
| Midbrain (PAG vicinity) ^a^ | 0,-22,-6 | | 4.46 | 235 | 3.60 | | 0.58 | | 0.46 | | reward; physiological; fear; conditioning; behaviors | | |
| Precentral_L ^a^ | -46,8,32 | | 5.18 | 46 | 7.72 | | 0.82 | | 0.30 | | phonological; reading; word; language; demands | | |
| ACC_pre_L | -8,40,-4 | | 5.43 | 3991 | 39.50 | | 0.37 | | 0.70 | | reward; money; gambling; valence; neurobiology | | |
| Frontal_Inf_Oper_R | 48,6,22 | | 4.41 | 970 | 42.72 | | 0.43 | | 0.65 | | reaction time; difficulty; visual; subtraction; recruited | | |
| Parietal_Sup_L | -24,-66,46 | | 4.86 | 357 | 22.51 | | 0.97 | | 0.29 | | working memory; numbers; processes; memory; color | | |
| Temporal_Inf_L | -52,-54,-16 | | 3.38 | 23 | 1.77 | | 0.65 | | 0.38 | | words; orthographic; visual word; semantic; word form | | |
| (b) SUDs (n=37) | | | | | | | | | | | | | |
| Cingulate_Mid_L ^a^ | -2,-12,32 | | 8.20 | 10317 | 1.54 | | 0.10 | | 0.92 | | personality traits; facilitation; traits; personality; adulthood | | |
| Frontal_Inf_Orb_2_R | 46,34,-10 | | 5.38 | 886 | 1.45 | | 0.59 | | 0.56 | | reappraisal; autobiographical; emotional; remembering; social | | |
| Angular_L | -34,-68,44 | | 5.35 | 620 | 30.75 | | 1.10 | | 0.33 | | retrieval; semantic; memory; memory retrieval; term memory | | |
| Frontal_Inf_Oper_L | -50,10,24 | | 3.88 | 132 | 5.46 | | 0.75 | | 0.47 | | phonological; language; semantic; words; orthographic | | |
| Temporal_Mid_L | -52,-64,-4 | | 3.63 | 62 | 2.66 | | 0.46 | | 0.62 | | visual; objects; sighted; visual motion; tools | | |
| Temporal_Mid_R | 58,-48,-6 | | 3.04 | 56 | 17.62 | | 0.31 | | 0.77 | | distress; numbers; simply; reveals; modulatory | | |
| Frontal_Inf_Oper_R | 48,6,22 | | 3.14 | 27 | 47.85 | | 0.37 | | 0.76 | | reaction time; difficulty; visual; subtraction; recruited | | |
| Temporal_Mid_R | 54,-52,16 | | 3.28 | 26 | 0.18 | | 0.24 | | 0.81 | | theory mind; social cognitive; mind; mental states; social interaction | | |
| Temporal_Mid_L | -66,-32,2 | | 3.00 | 27 | 16.52 | | -0.11 | | 0.92 | | speech; auditory; words; spoken; voice | | |
| Parietal_Inf_R | 44,-58,50 | | 3.14 | 26 | 18.12 | | 0.53 | | 0.60 | | choices; recognition memory; relational; partner; number | | |
| (c) BAs (n=16) | | | | | | | | | | | | | |
| Frontal_Inf_Tri_R | 52,18,18 | | 3.74 | 319 | 4.92 | | 0.98 | | 0.42 | | accurately; lexical; language; difficulty; everyday life | | |
| Frontal_Mid_2_L | -46,10,36 | | 3.76 | 257 | 9.31 | | 0.86 | | 0.53 | | verbal; stroop; chinese; processes; memory | | |
| Cerebellum_9_L | -6,-46,-54 | | 3.38 | 124 | 1.67 | | 0.31 | | 0.81 | | autonomic; nociceptive; imagine; noxious; pathophysiological | | |
| Calcarine_L | -12,-54,4 | | 3.19 | 46 | 8.96 | | 1.52 | | 0.21 | | navigation; constructed; autobiographical; divergent; scene | | |
| Cerebellum_8_L | -16,-58,-50 | | 2.79 | 25 | 12.68 | | 0.44 | | 0.75 | | coordination; profile; trajectories; handedness; motor performance | | |
| (d) SUDs > BAs | | | | | | | | | | | | | |
| Hippocampus_L ^a^ | -20,-10,-22 | | 2.09 | 563 | 30.65 | | 0.35 | | 0.61 | | emotional; emotion; faces; neutral; facial | | |
| ACC_pre_L ^a^ | -8,42,2 | | 2.16 | 563 | 43.18 | | 0.37 | | 0.64 | | money; amnestic; autobiographical; regulation; regulatory | | |
| Precuneus_L ^a^ | -4,-56,22 | | 2.26 | 88 | 9.43 | | 0.33 | | 0.60 | | autobiographical; default; default mode; autobiographical memory; dmn | | |
| (e) SUDs ∩ BAs | | | | | | | | | | | | | |
| Frontal_Inf_Oper_L | -50,8,28 | | 3.15 | 51 | NA | | NA | | NA | | phonological; language; reading; word; letter | | |
| Frontal_Inf_Oper_R | 48,4,22 | | 2.93 | 26 | NA | | NA | | NA | | blocked; discrimination; recruited; characters; hemisphere | | |

Brain regions showing significant hyperactivation during cue-reactivity across different addiction cohorts and subgroup comparisons. The table is organized as follows: (a) pooled sample of all SUDs and BAs (SUDs ∪ BAs); (b) SUDs subgroup only; (c) BAs subgroup only; (d) regions where SUDs exhibited greater activation than BAs (SUDs > BAs); and (e) shared regions between SUDs and BAs (SUDs ∩ BAs). Macroanatomical label: brain region name based on AAL atlas; MNI: Montreal Neurological Institute coordinates of the peak voxel (x,y,z); SDM-Z: peak z-value reflecting effect size; Voxels: cluster size; *I²* (%): heterogeneity statistic across studies; Egger’s bias and Egger’s *p*: publication bias estimates (not applicable for conjunction analyses); Function decoding: function decoding of the peak coordinates in Neurosynth. Results were significant at an uncorrected *p* < 0.005.

^a^ Survived to the FWE correction threshold of *p* < 0.05.

**Table S7. Subgroup‑specific and comparative neural activation patterns for addiction categories**

| **Macroanatomical label** | | **MNI (x,y,z)** | | | **SDM-Z** | | **Voxels** | | | ***I2 (%)*** | | **Egger’s bias** | **Egger’s p** | **Function decoding** |
| --- | --- | --- | --- | --- | --- | --- | --- | --- | --- | --- | --- | --- | --- | --- |
| (a) HUD (n=13) | | | | | | | | | | | | | | |
| Thal_MDl_R ^a^ | 6,-8,4 | | 4.79 | 5043 | | 14.60 | | 1.31 | 0.61 | | pain; reward; basal ganglia; basal; ganglia | | | |
| ACC_pre_L ^a^ | 0,44,10 | | 5.11 | 4576 | | 2.02 | | 1.29 | 0.56 | | default; independent component; mode; default mode; affective | | | |
| Frontal_Inf_Orb_2_R ^a^ | 44,34,-8 | | 5.12 | 1176 | | 4.45 | | 0.21 | 0.93 | | emotional; reappraisal; emotions; showing; emotional information | | | |
| Cerebellum_Crus1_L ^a^ | -16,-76,-24 | | 3.82 | 364 | | 12.29 | | 0.82 | 0.77 | | crus; lobules; facilitation; block; images acquired | | | |
| Parietal_Sup_L ^a^ | -22,-66,50 | | 4.24 | 104 | | 5.50 | | -0.29 | 0.91 | | visual; spatial; working memory; attention network; rotation | | | |
| Parietal_Inf_L ^a^ | -46,-50,48 | | 4.08 | 94 | | 5.11 | | -0.28 | 0.92 | | network; memory; networks involved; working memory; domain general | | | |
| Amygdala_R | 22,-4,-18 | | 4.46 | 979 | | 11.80 | | 2.38 | 0.25 | | emotional; faces; neutral; emotion; facial | | | |
| Temporal_Inf_R | 62,-58,-12 | | 4.21 | 321 | | 3.38 | | 0.37 | 0.89 | | requirements; lesser; correlating; emergence; underlying mechanisms | | | |
| Precuneus_L | -6,-52,18 | | 3.88 | 181 | | 3.52 | | 0.00 | 1.00 | | autobiographical; default; autobiographical memory; default mode; mode | | | |
| Caudate_R | 12,16,6 | | 3.96 | 156 | | 6.28 | | 0.97 | 0.70 | | reward; monetary; incentive; rewards; basal ganglia | | | |
| Precuneus_R | 2,-54,38 | | 3.04 | 58 | | 8.26 | | 1.29 | 0.59 | | mind tom; tom; theory mind; default; mode | | | |
| Temporal_Mid_L | -52,-64,-4 | | 3.54 | 45 | | 1.52 | | 0.73 | 0.76 | | visual; objects; sighted; visual motion; tools | | | |
| Fusiform_R | 26,-80,-14 | | 3.17 | 47 | | 13.86 | | 2.48 | 0.24 | | segregated; face ffa; faces; response time; visual | | | |
| Frontal_Inf_Oper_L | -50,10,24 | | 3.23 | 45 | | 9.46 | | -0.09 | 0.97 | | phonological; language; semantic; words; orthographic | | | |
| Lingual_L | -22,-86,-14 | | 3.02 | 33 | | 6.15 | | 2.34 | 0.26 | | face; navigation; visual; vision; faces | | | |
| Frontal_Inf_Oper_R | 50,6,20 | | 3.19 | 31 | | 26.49 | | -1.21 | 0.70 | | produced; working memory; spatial information; recruited; difficulty | | | |
| (b) AUD (n=13) | | | | | | | | | | |  | | | |
| Cingulate_Mid_R | 2,-30,42 | | 3.73 | 450 | | 6.44 | | 1.03 | 0.48 | | functional networks; default mode; network dmn; mode; default | | | |
| Temporal_Mid_L | -64,-38,6 | | 4.36 | 279 | | 6.25 | | -0.45 | 0.78 | | auditory; speech; voice; listening; words | | | |
| Rectus_L | -20,12,-14 | | 3.26 | 126 | | 13.24 | | 1.07 | 0.50 | | reward; reward anticipation; anticipation; monetary reward; ventral tegmental | | | |
| Frontal_Sup_Medial_L | -4,46,38 | | 3.37 | 101 | | 6.47 | | 1.26 | 0.37 | | negative positive; inferences; self referential; mentalizing; referential | | | |
| Angular_R | 38,-56,48 | | 3.27 | 82 | | 11.90 | | 1.03 | 0.52 | | working memory; memory; irrelevant; symbolic; numbers | | | |
| Frontal_Med_Orb_L | 0,52,-4 | | 3.10 | 48 | | 10.45 | | 0.30 | 0.85 | | default; default mode; mode; mode network; autobiographical | | | |
| Precuneus_R | 6,-62,36 | | 2.89 | 28 | | 4.49 | | 0.65 | 0.69 | | default; default mode; mode; mentalizing; person | | | |
| ParaHippocampal_L | -26,-40,-6 | | 2.99 | 26 | | 13.46 | | 1.38 | 0.34 | | navigation; encoding; scene; memory; scenes | | | |
| (c) IGD (n=12) | | | | | | | | | | |  | | | |
| Frontal_Inf_Tri_R | 52,20,22 | | 3.74 | 193 | | 3.67 | | 0.84 | 0.53 | | sts; sentence; expressions; word; decision task | | | |
| Frontal_Mid_2_L | -46,10,36 | | 3.20 | 96 | | 21.62 | | 0.86 | 0.60 | | verbal; stroop; chinese; processes; memory | | | |
| (d) HUD > AUD | | | | | | | | | | |  | | | |
| Frontal_Inf_Orb_2_R ^a^ | 48,36,-8 | | 3.04 | 973 | | 6.59 | | 0.51 | 0.61 | | reappraisal; emotional information; conveyed; emotional; knowledge | | | |
| Pallidum_R ^a^ | 22,4,2 | | 3.09 | 434 | | 11.41 | | 0.51 | 0.62 | | basal ganglia; ganglia; basal; motor; supplementary | | | |
| N_Acc_R ^a^ | 8,16,-6 | | 2.49 | 216 | | 46.35 | | 0.51 | 0.68 | | reward; monetary; anticipation; incentive; incentive delay | | | |
| Thal_MDm_R ^a^ | 4,-12,4 | | 2.73 | 110 | | 60.14 | | 0.61 | 0.65 | | reward; mesolimbic; pain; sexual; anticipation | | | |
| Temporal_Inf_R ^a^ | 62,-58,-10 | | 3.19 | 82 | | 21.52 | | 0.56 | 0.60 | | response times; compensation; vary; requirements; mild cognitive | | | |
| Caudate_L ^a^ | -10,8,10 | | 2.30 | 52 | | 31.57 | | 0.44 | 0.69 | | ganglia; basal ganglia; basal; reward; globus | | | |
| (e) HUD > IGD | | | | | | | | | | | | | | |
| Hippocampus_L ^a^ | -18,-8,-20 | | 3.30 | 989 | | 5.17 | | 0.51 | 0.58 | | emotional; emotion; faces; neutral; fearful | | | |
| ACC_pre_L ^a^ | -6,44,8 | | 2.78 | 782 | | 2.31 | | 0.48 | 0.61 | | mode network; mode; default mode; appraisal; default | | | |
| Insula_R ^a^ | 36,-16,14 | | 2.70 | 306 | | 41.76 | | 0.58 | 0.64 | | pain; painful; nociceptive; noxious; somatosensory | | | |
| Frontal_Inf_Orb_2_R ^a^ | 44,34,-8 | | 2.92 | 247 | | 3.53 | | 0.50 | 0.61 | | emotional; reappraisal; emotions; showing; emotional information | | | |
| Amygdala_R ^a^ | 20,-4,-16 | | 2.64 | 227 | | 1.70 | | 0.43 | 0.65 | | emotional; faces; emotion; neutral; facial | | | |
| Thal_MDm_R ^a^ | 6,-8,2 | | 2.60 | 140 | | 20.65 | | 0.50 | 0.65 | | reward; mesolimbic; pain; basal; responses | | | |
| Frontal_Sup_2_L ^a^ | -18,24,54 | | 2.23 | 102 | | 70.71 | | 0.66 | 0.66 | | shifts; sparse; memory performance; resting; know | | | |
| Caudate_L ^a^ | -8,4,8 | | 1.91 | 22 | | 2.14 | | 0.34 | 0.73 | | anticipation; incentive delay; reward; monetary; reward anticipation | | | |
| (f) IGD > AUD | | | | | | | | | | | | | | |
| Frontal_Inf_Tri_R ^a^ | 50,20,20 | | 2.58 | 67 | | 1.18 | | 0.40 | 0.60 | | inferior middle; spectrum disorder; higher levels; emotional; socially | | | |
| (g) HUD ∩ AUD | | | | | | | | | | | | | | |
| Cingulate_Mid_R | 4,-20,32 | | 3.14 | 30 | | NA | | NA | NA | | stop signal; response inhibition; stop; inhibit; inhibition | | | |
| Precuneus_L | -8,-52,24 | | 3.14 | 29 | | NA | | NA | NA | | default; autobiographical; default mode; mode network; mode | | | |

Meta‑analytic results for cue‑related hyperactivation within and between the three largest diagnostic subgroups: AUD, HUD and IGD. (a) AUD‑specific activation; (b) HUD‑specific activation; (c) IGD‑specific activation; (d) regions with greater activation in HUD than in AUD (HUD > AUD); (e) regions with greater activation in HUD than in IGD (HUD > IGD); (f) regions with greater activation in IGD than in AUD (IGD > AUD); (g) shared activation between HUD and AUD (HUD ∩ AUD). Macroanatomical label: brain region name based on AAL atlas; MNI: Montreal Neurological Institute coordinates of the peak voxel (x,y,z); SDM-*Z*: peak z-value reflecting effect size; Voxels: cluster size; *I²* (%): heterogeneity statistic across studies; Egger’s bias and Egger’s *p*: publication bias estimates (not applicable for conjunction analyses); Function decoding: function decoding of the peak coordinates in Neurosynth. Results were significant at an uncorrected *p* < 0.005.

^a^ Survived to the FWE correction threshold of *p* < 0.05.

**Table S8. Brain–behavior correlations between clinical measures and cue‑reactivity activation**

| **Macroanatomical label** | **MNI (x,y,z)** | **SDM-*Z*** | **Voxels** | ***I^2^* (%)** | **Egger’s bias** | **Egger’s *p*** | **Function decoding** |
| --- | --- | --- | --- | --- | --- | --- | --- |
| (a) Correlation between HUD and drug use duration (n=11) | | | | | | | |
| Hippocampus_L ^a^ | -18,-10,-22 | -3.60 | 1066 | 0.50 | 0.00 | 1.00 | emotional; faces; emotion; emotions; emotionally |
| Hippocampus_R ^a^ | 22,-6,-20 | -3.25 | 938 | 0.71 | 0.00 | 1.00 | faces; emotional; emotion; neutral; fear |
| Parietal_Inf_L ^a^ | -44,-50,48 | -2.57 | 1027 | 13.04 | 0.00 | 1.00 | working memory; memory; network; networks involved; question |
| Frontal_Inf_Tri_R ^a^ | 42,42,-2 | -3.41 | 450 | 8.77 | 0.00 | 1.00 | sensations; frontopolar; actively; matrix; force |
| Frontal_Sup_2_L ^a^ | -18,26,52 | -3.67 | 375 | 3.58 | 0.00 | 1.00 | control network; default; resting; default network; mode network |
| Temporal_Mid_L ^a^ | -48,-72,10 | -3.20 | 223 | 8.82 | 0.00 | 1.00 | motion; videos; mt; visual; video |
| Cingulate_Mid_L ^a^ | 0,-8,34 | -2.55 | 69 | 4.71 | 0.00 | 1.00 | pain; impulsivity; reward; impulsive; somatosensory cortices |
| Frontal_Mid_2_R | 48,46,14 | -3.36 | 97 | 9.68 | 0.00 | 1.00 | taken; choices; noxious; differentiate; ability |
| (b) Correlation between HUD and withdrawal duration (n=7) | | | | | | | |
| Frontal_Sup_Medial_L ^a^ | -2,44,24 | -3.46 | 385 | 9.14 | 0.00 | 1.00 | covariance; ica; networks involved; independent component; mentalizing |
| Mibrain | 2,-14,-2 | -3.01 | 37 | 2.85 | 0.00 | 1.00 | reward; unpleasant; mesolimbic; sexual; pleasant |
| (c) Correlation between HUD and total drug consumption (n=6) | | | | | | | |
| Frontal_Inf_Tri_R | 42,42,0 | -3.08 | 42 | 18.38 | 0.00 | 1.00 | matrix; pain; sensations; circuits; actively |
| (d) Correlation between HUD and treatment duration (n=6) | | | | | | | |
| Frontal_Inf_Oper_R | 46,8,22 | 3.76 | 52 | 1.05 | 0.00 | 1.00 | visual; difficulty; recruited; reaction time; task difficulty |
| (e) Correlation between AUD and depression (n=11) | | | | | | | |
| Frontal_Mid_2_L ^a^ | -26,18,42 | -4.01 | 63 | 4.03 | -0.96 | 0.46 | autobiographical memory; recall; default network; intact; judgments |
| (f) Correlation between AUD and withdrawal duration (n=7) | | | | | | | |
| Cingulate_Mid_L ^a^ | -4,-46,34 | 4.73 | 1234 | 2.67 | 0.07 | 0.97 | default; default mode; mode; mode network; dmn |
| Angular_R ^a^ | 50,-62,38 | 3.39 | 712 | 62.34 | 0.10 | 0.98 | default; mode network; default mode; networks; mode |
| Frontal_Sup_Medial_L ^a^ | -6,54,30 | 4.19 | 449 | 1.63 | 0.06 | 0.98 | mental states; tom; theory mind; mind tom; mind |
| Frontal_Sup_2_R | 26,20,38 | 3.38 | 61 | 60.75 | 0.10 | 0.98 | functional network; 70; addition; strategic; contributed |
| Temporal_Pole_Sup_R | 56,16,-6 | 3.22 | 57 | 62.65 | 0.09 | 0.98 | aversive; cognitive neuroscience; foot; code; involved |
| Temporal_Sup_R | 70,-12,0 | 3.39 | 29 | 65.17 | 0.10 | 0.98 | sounds; pitch; auditory; spoken; voice |
| (g) Correlation between AUD and symptom severity (n=7) | | | | | | | |
| ParaHippocampal_L ^a^ | -24,-42,-6 | -2.37 | 44 | 11.38 | -0.41 | 0.78 | scene; scenes; autobiographical; navigation; remembering |
| (h) Correlation between AUD and anxiety (n=5) | | | | | | | |
| Heschl_R | 46,-16,6 | 2.96 | 114 | 2.90 | 0.89 | 0.76 | auditory; listening; primary auditory; heschl; planum |
| (i) Correlation between IGD and weekly gaming hours (n=7) | | | | | | | |
| Frontal_Inf_Tri_L ^a^ | -44,36,16 | 3.09 | 35 | 14.00 | 0.00 | 1.00 | phonological; retrieval; word; demands; semantic |
| (j) Correlation between IGD and symptom severity (n=10) | | | | | | | |
| Cingulate_Mid_R | 2,-6,30 | 3.39 | 81 | 3.44 | 0.20 | 0.90 | involving; developmental; shift; structures involved; reward |
| Frontal_Inf_Oper_L | -48,10,28 | 3.04 | 62 | 0.22 | 0.23 | 0.88 | phonological; language; reading; word; semantic |

The table lists exploratory study-level meta-regressions by diagnostic subgroup and clinical measure, with direction, n, coordinates, heterogeneity, and functional decoding shown for each analysis as labeled. FWE-corrected peaks are marked with superscript a; other displayed peaks meet only the exploratory uncorrected threshold. Macroanatomical label: brain region name based on AAL atlas; MNI: Montreal Neurological Institute coordinates of the peak voxel (x,y,z); SDM-*Z*: peak z-value reflecting effect size; Voxels: cluster size; *I²* (%): heterogeneity statistic across studies; Egger’s bias and Egger’s *p*: publication bias estimates (not applicable for conjunction analyses); Function decoding: function decoding of the peak coordinates in Neurosynth; n: the number of contrasts involved in the meta-regression. Results were significant at an uncorrected *p* < 0.005.

^a^ Survived to the FWE correction threshold of *p* < 0.05.

**Table S9. Demographic sensitivity analyses for subgroup comparisons**

| **Macroanatomical label** | | **MNI (x,y,z)** | | | **SDM-Z** | | **Voxels** | | **I2 (%)** | | **Egger’s bias** | | **Egger’s p** | **Function decoding** |
| --- | --- | --- | --- | --- | --- | --- | --- | --- | --- | --- | --- | --- | --- | --- |
| **(A) Control age** | | | | | | | | | | | | | | |
| (a) SUDs (n=36) > BAs (n=16) (control age) | | | | | | | | | | | | | | |
| ACC_sub_L ^a^ | -6,36,-4 | | 3.56 | 272 | | 13.80 | | 0.56 | | 0.42 | | money; rewards; value; characterizing; states | | |
| Hippocampus_L ^a^ | -24,-8,-22 | | 2.53 | 286 | | 46.27 | | 0.55 | | 0.48 | | emotional; neutral; fearful; faces; emotion | | |
| Precuneus_L ^a^ | -2,-56,22 | | 2.76 | 239 | | 15.51 | | 0.48 | | 0.48 | | autobiographical; default; default mode; mode; ica | | |
| Frontal_Sup_2_L ^a^ | -22,28,46 | | 2.61 | 135 | | 50.81 | | 0.59 | | 0.47 | | autobiographical; undergoing; default; default network; functional networks | | |
| (b) HUD (n=13) > AUD (n=12) (control age) | | | | | | | | | | | | | | |
| Frontal_Inf_Orb_2_R ^a^ | 50,36,-8 | | 2.92 | 1235 | | 5.49 | | 0.49 | | 0.69 | | knowledge; preferentially; personality traits; involved cognitive; conveyed | | |
| Pallidum_R ^a^ | 22,4,2 | | 2.93 | 248 | | 12.09 | | 0.51 | | 0.68 | | basal ganglia; ganglia; basal; motor; supplementary | | |
| N_Acc_R ^a^ | 8,16,-6 | | 2.38 | 177 | | 45.65 | | 0.51 | | 0.74 | | reward; monetary; anticipation; incentive; incentive delay | | |
| Temporal_Inf_R ^a^ | 62,-58,-10 | | 3.04 | 65 | | 23.66 | | 0.56 | | 0.68 | | response times; compensation; vary; requirements; mild cognitive | | |
| (c) IGD (n=12) > AUD (n=12) (control age) | | | | | | | | | | | | | | |
| Frontal_Mid_2_R ^a^ | 42,42,16 | | 2.93 | 149 | | 73.69 | | 1.39 | | 0.32 | | nociceptive; painful; response inhibition; administered; executive | | |
| Temporal_Pole_Sup_L ^a^ | -30,8,-20 | | -3.11 | 1540 | | 74.79 | | -1.28 | | 0.33 | | emotional; emotion; neutral; coupling; neutral pictures | | |
| undefined ^a^ | 18,-8,-32 | | -2.90 | 857 | | 74.04 | | -1.11 | | 0.39 | | reward anticipation; treatments; emotionally; conditioning; threat | | |
| Cingulate_Post_L ^a^ | -2,-42,24 | | -3.64 | 655 | | 79.08 | | -1.48 | | 0.27 | | recollection; retrieval; memory; spin labeling; remembered | | |
| Supp_Motor_Area_L ^a^ | -4,-8,76 | | -2.92 | 702 | | 72.87 | | -1.10 | | 0.39 | | supplementary; compatible; motor; bold responses; essential | | |
| undefined ^a^ | -2,52,-28 | | -2.92 | 649 | | 73.59 | | -1.09 | | 0.39 | | alcohol; years old; elucidated; negativity; mapped | | |
| undefined ^a^ | -2,-8,-20 | | -2.82 | 514 | | 72.52 | | -1.09 | | 0.40 | | runs; hypoactivation; cross modal; validity; treatment | | |
| Precuneus_R ^a^ | 12,-76,50 | | -2.89 | 394 | | 73.71 | | -1.05 | | 0.41 | | construct; number; numbers; consistently; overlapping | | |
| Temporal_Sup_L ^a^ | -64,-12,2 | | -2.87 | 150 | | 73.24 | | -1.14 | | 0.39 | | voice; speech; auditory; sounds; speaker | | |
| Frontal_Mid_2_L ^a^ | -24,26,50 | | -2.92 | 56 | | 72.23 | | -1.16 | | 0.38 | | default; default network; autobiographical; networks; deactivation | | |
| Angular_R ^a^ | 38,-70,40 | | -2.68 | 57 | | 65.80 | | -1.00 | | 0.43 | | retrieval; wm task; task difficulty; spatial; abstract | | |
| **(B) Male-only sensitivity analysis** | | | | | | | | | | | | | | |
| (a) SUDs (n=20) > BAs (n=10) (male-only) | | | | | | | | | | | | | | |
| Insula_R ^a^ | 34,-16,14 | | 3.54 | 549 | | 30.74 | | 0.73 | | 0.56 | | pain; noxious; painful; nociceptive; chronic pain | | |
| Precentral_L ^a^ | -48,8,36 | | -2.81 | 32 | | 0.41 | | -0.50 | | 0.63 | | verbal; reading; phonological; demands; language | | |
| (b) HUD (n=10) > IGD (n=7) (male-only) | | | | | | | | | | | | | | |
| Insula_R ^a^ | 38,-18,14 | | 5.35 | 847 | | 0.27 | | 1.16 | | 0.42 | | pain; painful; nociceptive; noxious; somatosensory | | |
| Precentral_L ^a^ | -50,8,38 | | -3.09 | 106 | | 1.26 | | -0.73 | | 0.62 | | reading; demands; processes; phonological; language | | |
| Occipital_Mid_R ^a^ | 34,-82,32 | | -2.99 | 183 | | 15.29 | | -0.62 | | 0.66 | | locations; visuospatial; v1; rotation; lateralization | | |

Age-adjusted diagnostic comparisons (A) and comparisons restricted to all-male patient and control samples (B) are shown. The male-only analyses are not male-proportion covariate models. Macroanatomical label: brain region name based on AAL atlas; MNI: Montreal Neurological Institute coordinates of the peak voxel (x,y,z); SDM-*Z*: peak z-value reflecting effect size; Voxels: cluster size; *I²* (%): heterogeneity statistic across studies; Egger’s bias and Egger’s *p*: publication bias estimates (not applicable for conjunction analyses); Function decoding: function decoding of the peak coordinates in Neurosynth. Results were significant at an uncorrected *p* < 0.005.

^a^ Survived to the FWE correction threshold of *p* < 0.05.

**Table S10. Sensitivity analyses in the contrast “addiction cues vs. control cues”**

| Macroanatomical label | | MNI (x,y,z) | | SDM-Z | | Voxels | | I2 (%) | | Egger’s bias | | Egger’s p | | Function decoding |
| --- | --- | --- | --- | --- | --- | --- | --- | --- | --- | --- | --- | --- | --- | --- |
| (a) SUDs ∪ BAs (n=45) | | | | | | | | | | | | | | |
| Olfactory_L ^a^ | -10,12,-14 | | 4.55 | | 3002 | | 4.54 | | 0.86 | | 0.29 | | reward; incentive; monetary; motivation; anticipation | |
| Cingulate_Mid_R ^a^ | 4,-18,30 | | 4.64 | | 2357 | | 41.74 | | 1.15 | | 0.24 | | stop signal; response inhibition; stop; inhibitory; inhibition | |
| Midbrain (PAG vicinity) ^a^ | 0,-22,-6 | | 4.73 | | 401 | | 5.03 | | 0.50 | | 0.55 | | reward; physiological; fear; conditioning; behaviors | |
| Frontal_Inf_Oper_L ^a^ | -48,10,26 | | 5.33 | | 144 | | 31.87 | | 1.40 | | 0.13 | | phonological; semantic; language; word; reading | |
| Frontal_Inf_Oper_R | 48,6,22 | | 4.36 | | 379 | | 45.61 | | 0.29 | | 0.79 | | reaction time; difficulty; visual; subtraction; recruited | |
| Frontal_Inf_Orb_2_R | 46,28,-8 | | 3.86 | | 284 | | 0.21 | | 0.22 | | 0.80 | | emotional; affective; social; negative affect; interpersonal | |
| Parietal_Sup_L | -24,-66,46 | | 4.68 | | 149 | | 21.08 | | 0.51 | | 0.60 | | working memory; numbers; processes; memory; color | |
| (b) SUDs (n=32) | | | | | | | | | | | | | | |
| ACC_sup_L ^a^ | 0,36,14 | | 5.64 | | 5964 | | 2.88 | | 0.72 | | 0.54 | | reward; options; money; joint; reward anticipation | |
| Cingulate_Mid_L ^a^ | -2,-14,34 | | 7.09 | | 3616 | | 0.89 | | 0.34 | | 0.76 | | pain; personality traits; believed; neurofunctional; stimulus response | |
| Parietal_Sup_L | -24,-66,46 | | 5.63 | | 612 | | 10.37 | | 0.87 | | 0.44 | | working memory; numbers; processes; memory; color | |
| Frontal_Inf_Orb_2_R | 42,42,-12 | | 4.98 | | 546 | | 10.16 | | 0.82 | | 0.51 | | behavioral evidence; graph; neurological; potentially; emotional faces | |
| Frontal_Inf_Oper_L | -50,10,24 | | 4.29 | | 175 | | 4.77 | | 0.78 | | 0.48 | | phonological; language; semantic; words; orthographic | |
| Temporal_Mid_L | -52,-64,-4 | | 4.17 | | 116 | | 2.27 | | 0.53 | | 0.63 | | visual; objects; sighted; visual motion; tools | |
| Temporal_Mid_L | -66,-40,0 | | 3.29 | | 77 | | 12.21 | | -0.09 | | 0.94 | | listening; language; meaning; verbs; english | |
| Temporal_Mid_R | 58,-48,-6 | | 3.30 | | 76 | | 16.49 | | 0.30 | | 0.80 | | distress; numbers; simply; reveals; modulatory | |
| Parietal_Inf_R | 44,-58,50 | | 3.65 | | 58 | | 15.56 | | 0.56 | | 0.62 | | choices; recognition memory; relational; partner; number | |
| Frontal_Inf_Oper_R | 48,6,22 | | 3.44 | | 40 | | 47.81 | | 0.35 | | 0.81 | | reaction time; difficulty; visual; subtraction; recruited | |
| Temporal_Inf_R | 50,-56,-20 | | 3.32 | | 28 | | 4.89 | | 1.28 | | 0.22 | | faces; face; ffa; spatial information; actively | |
| Caudate_R | 14,14,8 | | 2.95 | | 23 | | 37.69 | | 0.77 | | 0.58 | | basal ganglia; ganglia; reward; basal; seeking | |
| (c) BAs (n=13) | | | | | | | | | | | | | | |
| Precentral_L | -48,8,34 | | 3.88 | | 252 | | 32.54 | | 1.30 | | 0.41 | | reading; phonological; demands; language; required | |
| Frontal_Inf_Oper_R | 52,16,18 | | 4.03 | | 215 | | 15.42 | | 1.05 | | 0.47 | | recruited; sequences; implementation; subtraction; broca | |
| Cerebellum_8_L | -16,-58,-52 | | 2.92 | | 96 | | 10.02 | | 0.38 | | 0.81 | | trajectories; crus; profile; coordination; noxious | |
| Calcarine_L | -12,-52,4 | | 3.24 | | 62 | | 12.65 | | 1.84 | | 0.19 | | navigation; scene; mental; autobiographical; constructed | |
| (d) SUDs > BAs | | | | | | | | | | | | | | |
| Hippocampus_L ^a^ | -22,-10,-22 | | 2.27 | | 466 | | 34.29 | | 0.40 | | 0.59 | | emotional; neutral; faces; fearful; threat | |
| (e) SUDs ∩ BAs | | | | | | | | | | | | | | |
| Precentral_L | -50,8,30 | | 3.26 | | 49 | | NA | | NA | | NA | | phonological; reading; language; word; calculation | |
| (f) HUD (n=10) | | | | | | | | | | | | | | |
| ParaHippocampal_R ^a^ | 18,0,-20 | | 5.4 | | 6051 | | 5.58 | | 3.16 | | 0.11 | | emotional; neutral; faces; fear; emotion | |
| ACC_sup_L a | -2,34,24 | | 4.983 | | 3047 | | 4.50 | | 2.02 | | 0.39 | | monetary; impulsivity; rewards; abuse; losses | |
| Frontal_Inf_Orb_2_R ^a^ | 40,38,-12 | | 5.441 | | 1276 | | 7.99 | | 0.08 | | 0.98 | | retrieved; autobiographical; emotions; period; relies | |
| Parietal_Sup_L ^a^ | -24,-66,50 | | 5.047 | | 1216 | | 8.30 | | -0.02 | | 1.00 | | attention network; english; spatial; visual; working memory | |
| Cingulate_Mid_R ^a^ | 2,-12,32 | | 5.321 | | 531 | | 2.88 | | -0.25 | | 0.93 | | connectivity anterior; pain; fluctuations; personality traits; attenuation | |
| Precuneus_L ^a^ | -6,-52,16 | | 4.317 | | 507 | | 3.13 | | -0.32 | | 0.91 | | autobiographical; episodic; episodic memory; memory retrieval; autobiographical memory | |
| Caudate_R ^a^ | 14,14,8 | | 4.634 | | 128 | | 5.02 | | 0.56 | | 0.83 | | basal ganglia; ganglia; reward; basal; seeking | |
| Temporal_Inf_R ^a^ | 60,-60,-12 | | 5.087 | | 38 | | 10.30 | | 0.97 | | 0.71 | | confirmed; segregated; extra; streams; connectivity analyses | |
| Temporal_Mid_L | -52,-64,-4 | | 3.837 | | 169 | | 0.92 | | 1.10 | | 0.64 | | visual; objects; sighted; visual motion; tools | |
| Occipital_Inf_R | 30,-82,-12 | | 3.63 | | 126 | | 13.15 | | 2.71 | | 0.21 | | faces; face; visual; perception; face ffa | |
| Occipital_Inf_L | -22,-90,-10 | | 3.782 | | 118 | | 10.11 | | 3.03 | | 0.16 | | naming; reading; visual; color; response time | |
| Frontal_Inf_Oper_L | -48,10,28 | | 3.655 | | 114 | | 2.89 | | 0.37 | | 0.90 | | phonological; language; reading; word; semantic | |
| Frontal_Inf_Oper_R | 50,6,20 | | 3.495 | | 82 | | 18.86 | | -0.96 | | 0.76 | | produced; working memory; spatial information; recruited; difficulty | |
| Frontal_Sup_Medial_R | 6,38,50 | | 3.102 | | 26 | | 9.90 | | 2.96 | | 0.19 | | cognitive; voxelwise; key role; mind; reappraisal | |
| (g) AUD (n=13) | | | | | | | | | | | | | | |
| Cingulate_Mid_R | 2,-30,42 | | 3.725 | | 450 | | 6.44 | | 1.03 | | 0.48 | | functional networks; default mode; network dmn; mode; default | |
| Temporal_Mid_L | -64,-38,6 | | 4.359 | | 279 | | 6.25 | | -0.45 | | 0.78 | | auditory; speech; voice; listening; words | |
| Rectus_L | -20,12,-14 | | 3.256 | | 126 | | 13.24 | | 1.07 | | 0.50 | | reward; reward anticipation; anticipation; monetary reward; ventral tegmental | |
| Frontal_Sup_Medial_L | -4,46,38 | | 3.371 | | 101 | | 6.47 | | 1.26 | | 0.37 | | negative positive; inferences; self referential; mentalizing; referential | |
| Angular_R | 38,-56,48 | | 3.274 | | 82 | | 11.90 | | 1.03 | | 0.52 | | working memory; memory; irrelevant; symbolic; numbers | |
| Frontal_Med_Orb_L | 0,52,-4 | | 3.103 | | 48 | | 10.45 | | 0.30 | | 0.85 | | default; default mode; mode; mode network; autobiographical | |
| Precuneus_R | 6,-62,36 | | 2.893 | | 28 | | 4.49 | | 0.65 | | 0.69 | | default; default mode; mode; mentalizing; person | |
| ParaHippocampal_L | -26,-40,-6 | | 2.994 | | 26 | | 13.46 | | 1.38 | | 0.34 | | navigation; encoding; scene; memory; scenes | |
| (h) IGD (n=10) | | | | | | | | | | | | | | |
| Frontal_Inf_Tri_R | 52,20,22 | | 4.061 | | 182 | | 6.57 | | 1.18 | | 0.42 | | sts; sentence; expressions; word; decision task | |
| Precentral_L | -52,2,32 | | 3.365 | | 126 | | 4.35 | | 0.43 | | 0.78 | | motor; execution; movements; motor imagery; action | |
| (i) HUD > AUD | | | | | | | | | | | | | | |
| N_Acc_R ^a^ | 6,14,-6 | | 3.082 | | 642 | | 3.69 | | 0.55 | | 0.58 | | reward; monetary; anticipation; incentive delay; mesolimbic | |
| Pallidum_R ^a^ | 22,4,2 | | 3.725 | | 591 | | 6.15 | | 0.61 | | 0.54 | | basal ganglia; ganglia; basal; motor; supplementary | |
| Frontal_Inf_Orb_2_R ^a^ | 48,40,-8 | | 3.475 | | 535 | | 10.80 | | 0.59 | | 0.57 | | focusing; middle inferior; recruit; accurate | |
| Thal_MDm_R ^a^ | 4,-12,4 | | 3.334 | | 266 | | 57.38 | | 0.75 | | 0.57 | | reward; mesolimbic; pain; sexual; anticipation | |
| Temporal_Inf_R ^a^ | 62,-58,-10 | | 3.806 | | 116 | | 8.45 | | 0.67 | | 0.52 | | response times; compensation; vary; requirements; mild cognitive | |
| Parietal_Inf_L ^a^ | -44,-52,50 | | 3.401 | | 52 | | 25.74 | | 0.64 | | 0.57 | | retention; memory; fronto; network; retrieval | |
| Caudate_L | -10,8,10 | | 3.024 | | 20 | | 18.22 | | 0.56 | | 0.60 | | ganglia; basal ganglia; basal; reward; globus | |
| (j) HUD > IGD | | | | | | | | | | | | | | |
| Hippocampus_L ^a^ | -18,-6,-22 | | 3.689 | | 1767 | | 14.24 | | 0.73 | | 0.48 | | emotional; neutral; fearful; fear; emotion | |
| ParaHippocampal_R ^a^ | 18,-2,-20 | | 3.178 | | 1066 | | 5.47 | | 0.58 | | 0.57 | | emotional; faces; neutral; fear; emotion | |
| ACC_sup_L ^a^ | -2,36,20 | | 2.6 | | 281 | | 4.89 | | 0.45 | | 0.66 | | cognitive emotional; nociceptive; inhibition; discriminative; connectivity networks | |
| Frontal_Inf_Orb_2_R ^a^ | 44,36,-12 | | 3.287 | | 207 | | 1.61 | | 0.60 | | 0.56 | | autobiographical; disruption; valence; relies; adulthood | |
| Frontal_Sup_2_L ^a^ | -18,22,56 | | 2.45 | | 197 | | 51.74 | | 0.64 | | 0.64 | | correlation analyses; comprehensive; memory performance; appear; demand | |
| Parietal_Inf_L ^a^ | -44,-52,50 | | 2.759 | | 48 | | 29.73 | | 0.63 | | 0.61 | | retention; memory; fronto; network; retrieval | |
| Thal_MDm_L | -4,-6,4 | | 2.935 | | 26 | | 7.44 | | 0.57 | | 0.59 | | sexual; autobiographical; replicate; reactions; structures | |
| (k) HUD ∩ AUD | | | | | | | | | | | | | | |
| Precuneus_L | -6,-56,24 | | 3.162 | | 67 | | NA | | NA | | NA | | autobiographical; autobiographical memory; remembering; default; default mode | |
| Cingulate_Mid_R | 2,-20,32 | | 3.264 | | 34 | | NA | | NA | | NA | | stop signal; response inhibition; sentence comprehension; inhibit; producing | |
| (l) HUD ∩ IGD | | | | | | | | | | | | | | |
| Precentral_L | -50,8,30 | | 2.939 | | 22 | | NA | | NA | | NA | | phonological; reading; language; word; calculation | |

Sensitivity analyses restricted to contrast (addiction cues vs. control cues) are organized as labeled, including the pooled sample, diagnostic categories and subtypes, direct comparisons, and conjunctions. Macroanatomical label: brain region name based on AAL atlas; MNI: Montreal Neurological Institute coordinates of the peak voxel (x,y,z); SDM-Z: peak z-value reflecting effect size; Voxels: cluster size; *I²* (%): heterogeneity statistic across studies; Egger’s bias and Egger’s *p*: publication bias estimates (not applicable for conjunction analyses); Function decoding: function decoding of the peak coordinates in Neurosynth. Results were significant at an uncorrected *p* < 0.005.

^a^ Survived to the FWE correction threshold of *p* < 0.05.

**Table S11. Sensitivity analyses in different cue types**

| **Macroanatomical label** | **MNI (x,y,z)** | **SDM-*Z*** | **Voxels** | ***I^2^* (%)** | **Egger’s bias** | **Egger’s *p*** | **Function decoding** |
| --- | --- | --- | --- | --- | --- | --- | --- |
| (a) SUDs ∪ BAs (video) (n=9) | | | | | | | |
| Frontal_Med_Orb_R ^a^ | 2,50,-4 | 3.78 | 277 | 1.62 | -0.17 | 0.91 | default; default mode; mode; mode network; networks |
| Frontal_Sup_Medial_R ^a^ | 2,52,22 | 3.46 | 30 | 1.75 | 0.21 | 0.89 | beliefs; mentalizing; default; theory mind; mind |
| Precuneus_R | 10,-66,54 | 3.79 | 339 | 13.09 | 1.74 | 0.22 | spatial; working memory; shared; navigation; virtual |
| Cerebellum_Crus2_L | -4,-84,-24 | 3.07 | 53 | 11.08 | 1.68 | 0.25 | crus; lobules; mr; handedness; loop |
| Precuneus_L | 0,-60,32 | 3.01 | 27 | 7.41 | -0.63 | 0.68 | theory mind; independent component; mentalizing; social; default |
| (b) SUDs ∪ BAs (pic) (n=44) | | | | | | | |
| Cingulate_Mid_R ^a^ | 4,-18,30 | 4.62 | 2426 | 39.77 | 1.32 | 0.19 | stop signal; response inhibition; stop; inhibitory; inhibition |
| ACC_sup_L ^a^ | -2,36,12 | 4.66 | 1949 | 2.28 | 0.53 | 0.59 | value; reward; positive negative; money; valence |
| Midbrain ^a^ | 4,-20,-4 | 4.21 | 190 | 28.65 | 1.00 | 0.31 | reward; ventral tegmental; tegmental; flexibly; mesolimbic |
| Frontal_Inf_Orb_2_R | 46,32,-10 | 4.58 | 496 | 3.82 | 0.25 | 0.79 | reappraisal; emotional; remembering; autobiographical; social |
| Parietal_Sup_L | -24,-66,46 | 5.35 | 316 | 27.37 | 1.21 | 0.24 | working memory; numbers; processes; memory; color |
| Frontal_Inf_Oper_L | -50,10,24 | 3.98 | 197 | 15.50 | 0.85 | 0.38 | phonological; language; semantic; words; orthographic |
| Frontal_Inf_Oper_R | 46,8,24 | 3.61 | 113 | 60.90 | 1.15 | 0.35 | visual; characters; demands; interference; reaction |
| Supp_Motor_Area_L | -8,24,54 | 3.66 | 101 | 12.37 | 0.59 | 0.55 | semantic; chinese; names; judgments; remembered |
| Frontal_Inf_Tri_R | 48,22,16 | 3.61 | 94 | 14.94 | 1.08 | 0.25 | sentences; socially; inferior middle; native; basic |
| Temporal_Inf_R | 64,-56,-8 | 3.20 | 78 | 36.99 | 0.64 | 0.57 | taken suggest; healthy young; reasoning; vary; concentration |
| (c) SUDs (pic) (n=33) | | | | | | | |
| Cingulate_Mid_R ^a^ | 2,-14,34 | 7.28 | 8131 | 1.23 | 0.27 | 0.81 | retrieved; believed; control processes; comprised; personality traits |
| Frontal_Inf_Orb_2_R ^a^ | 46,32,-10 | 5.25 | 47 | 1.09 | 0.60 | 0.62 | reappraisal; emotional; remembering; autobiographical; social |
| ACC_pre_L | 0,38,14 | 5.77 | 4198 | 4.10 | 1.04 | 0.36 | mesolimbic; value; reward; money; joint |
| Parietal_Sup_L | -24,-66,46 | 5.59 | 506 | 11.59 | 0.85 | 0.51 | working memory; numbers; processes; memory; color |
| Temporal_Inf_R | 64,-56,-6 | 3.49 | 163 | 31.11 | 0.08 | 0.95 | current functional; systematic; 37; reasoning; differed |
| Frontal_Inf_Oper_L | -50,10,24 | 4.01 | 144 | 4.47 | 0.76 | 0.54 | phonological; language; semantic; words; orthographic |
| Temporal_Inf_L | -48,-60,-10 | 3.62 | 62 | 0.36 | 0.48 | 0.66 | words; word; visual word; reading; word form |
| Temporal_Mid_L | -66,-32,2 | 3.12 | 45 | 16.47 | -0.12 | 0.93 | speech; auditory; words; spoken; voice |
| Frontal_Inf_Oper_R | 48,6,22 | 3.17 | 33 | 47.60 | 0.32 | 0.83 | reaction time; difficulty; visual; subtraction; recruited |
| Parietal_Inf_R | 44,-58,50 | 3.19 | 29 | 18.03 | 0.59 | 0.64 | choices; recognition memory; relational; partner; number |
| (d) BAs (pic) (n=11) | | | | | | | |
| Frontal_Inf_Tri_R | 52,20,20 | 3.33 | 61 | 7.72 | 1.33 | 0.34 | hyperactivation; language; higher levels; greater; difficulty |
| Cerebellum_8_L | -18,-58,-48 | 2.94 | 42 | 6.55 | 0.42 | 0.79 | crus; motor performance; materials; coordination; trajectories |
| (e) SUDs > BAs (pic) | | | | | | | |
| Precuneus_L ^a^ | 2,-60,32 | 1.89 | 190 | 5.64 | 0.29 | 0.68 | default; independent component; theory mind; social; default mode |
| (f) HUD (pic) (n=12) | | | | | | | |
| ACC_pre_L ^a^ | -6,44,8 | 5.11 | 3852 | 5.12 | 2.90 | 0.31 | mode network; mode; default mode; appraisal; default |
| Thal_MDl_R ^a^ | 6,-8,4 | 4.82 | 3808 | 14.32 | 3.34 | 0.31 | pain; reward; basal ganglia; basal; ganglia |
| Frontal_Inf_Orb_2_R ^a^ | 44,34,-8 | 5.15 | 1005 | 5.20 | 1.30 | 0.70 | emotional; reappraisal; emotions; showing; emotional information |
| Cerebellum_Crus1_L ^a^ | -16,-76,-24 | 3.97 | 429 | 12.97 | 1.86 | 0.64 | crus; lobules; facilitation; block; images acquired |
| Parietal_Sup_L ^a^ | -24,-66,48 | 4.32 | 141 | 12.91 | 0.66 | 0.86 | working memory; attention network; visual; interference; rotation |
| Parietal_Inf_L ^a^ | -46,-48,48 | 4.07 | 134 | 11.99 | 0.55 | 0.89 | working memory; memory; networks involved; network; processes |
| Precuneus_L ^a^ | -6,-52,18 | 3.86 | 50 | 5.29 | 0.94 | 0.78 | autobiographical; default; autobiographical memory; default mode; mode |
| Putamen_L ^a^ | -20,6,-2 | 3.45 | 26 | 8.04 | 0.79 | 0.85 | basal ganglia; ganglia; basal; reward; monetary |
| Pallidum_R | 22,6,2 | 4.28 | 411 | 7.97 | 2.60 | 0.40 | basal ganglia; ganglia; basal; motor; movement |
| Caudate_R | 14,16,8 | 3.98 | 166 | 29.42 | 2.91 | 0.39 | basal ganglia; ganglia; basal; reward; seeking |
| Hippocampus_L | -28,-4,-26 | 3.55 | 153 | 10.72 | 0.44 | 0.91 | neutral; fearful; faces; neutral faces; emotional |
| Temporal_Inf_R | 62,-58,-12 | 3.82 | 127 | 9.25 | 0.19 | 0.96 | requirements; lesser; correlating; emergence; underlying mechanisms |
| Precentral_L | -50,8,30 | 3.28 | 40 | 7.46 | 0.47 | 0.90 | phonological; reading; language; word; calculation |
| Precuneus_L | -2,-56,40 | 2.91 | 40 | 12.43 | 2.88 | 0.40 | theory mind; mind tom; tom; mind; mental states |
| Frontal_Inf_Oper_R | 50,6,20 | 3.11 | 33 | 26.18 | -1.20 | 0.80 | produced; working memory; spatial information; recruited; difficulty |
| (g) AUD (pic) (n=12) | | | | | | | |
| Cingulate_Mid_R | 4,-28,38 | 3.59 | 375 | 13.77 | 1.07 | 0.48 | retrieval; default mode; mode; mode network; default |
| Temporal_Mid_L | -64,-40,4 | 4.40 | 233 | 7.76 | -0.41 | 0.80 | auditory; speech; language; sentences; listening |
| OFCmed_L | -18,24,-18 | 3.14 | 98 | 4.30 | 0.66 | 0.65 | olfactory; widespread; conditioning; valence; skin conductance |
| Frontal_Sup_Medial_L | -4,46,38 | 3.26 | 85 | 8.42 | 0.91 | 0.52 | negative positive; inferences; self referential; mentalizing; referential |
| Occipital_Mid_R | 44,-74,24 | 3.17 | 68 | 3.74 | 0.98 | 0.53 | correspond; episodic memory; retrieval; actively; episodic |
| ParaHippocampal_L | -26,-40,-6 | 3.06 | 22 | 10.03 | 1.23 | 0.40 | navigation; encoding; scene; memory; scenes |
| (h) IGD (pic) (n=9) | | | | | | | |
| Frontal_Inf_Tri_R | 50,22,20 | 3.19 | 60 | 10.17 | 1.63 | 0.26 | inferior middle; word; sts; sentence; facial |
| (i) HUD > AUD (pic) | | | | | | | |
| Frontal_Inf_Orb_2_R ^a^ | 48,40,-8 | 3.13 | 456 | 13.08 | 0.52 | 0.64 | 47; focusing; middle inferior; recruit; accurate |
| Pallidum_R ^a^ | 22,4,2 | 3.12 | 180 | 12.52 | 0.54 | 0.62 | basal ganglia; ganglia; basal; motor; supplementary |
| Thal_MDm_R ^a^ | 4,-8,6 | 2.73 | 67 | 30.74 | 0.50 | 0.67 | reward; sexual; aversive; monetary reward; subjective |
| (j) HUD > IGD (pic) | | | | | | | |
| ACC_sup_L ^a^ | -2,36,26 | 2.58 | 488 | 1.87 | 0.43 | 0.69 | preferences; losses; networks; pain; rewards |
| Frontal_Inf_Orb_2_R ^a^ | 44,36,-12 | 2.71 | 137 | 4.10 | 0.52 | 0.65 | autobiographical; disruption; valence; relies; adulthood |
| Hippocampus_L ^a^ | -18,-6,-22 | 2.32 | 139 | 26.26 | 0.49 | 0.69 | emotional; neutral; fearful; fear; emotion |
| Thal_MDm_R ^a^ | 6,-8,2 | 2.39 | 129 | 23.78 | 0.52 | 0.68 | reward; mesolimbic; pain; basal; responses |
| (k) HUD ∩ AUD (pic) | | | | | | | |
| Precuneus_L | -6,-56,24 | 2.94 | 29 | NA | NA | NA | autobiographical; autobiographical memory; remembering; default; default mode |
| Cingulate_Mid_L | -2,-20,32 | 3.08 | 28 | NA | NA | NA | noxious; stop signal; incorrect; pain; sensation |

Picture- and video-cue analyses are reported separately. The video subset was analyzed only for the pooled addiction group because of its small size. Macroanatomical label: brain region name based on AAL atlas; MNI: Montreal Neurological Institute coordinates of the peak voxel (x,y,z); SDM-Z: peak z-value reflecting effect size; Voxels: cluster size; *I²* (%): heterogeneity statistic across studies; Egger’s bias and Egger’s *p*: publication bias estimates (not applicable for conjunction analyses); Function decoding: function decoding of the peak coordinates in Neurosynth. Results were significant at an uncorrected *p* < 0.005.

^a^ Survived to the FWE correction threshold of *p* < 0.05.

**Table S12. Sensitivity analyses for methadone maintenance treatment in HUD**

| **Macroanatomical label** | **MNI (x,y,z)** | **SDM-*Z*** | **Voxels** | ***I^2^* (%)** | **Egger’s bias** | **Egger’s *p*** | **Function decoding** |
| --- | --- | --- | --- | --- | --- | --- | --- |
| (a) HUD without methadone therapy (n=7) | | | | | | | |
| ACC_pre_L ^a^ | -2,44,10 | 4.60 | 1501 | 4.57 | 1.15 | 0.66 | default mode; default; arousal; mode; independent component |
| Cingulate_Mid_L ^a^ | 0,8,38 | 3.46 | 53 | 6.60 | 0.33 | 0.91 | pain; painful; somatosensory; primary secondary; autonomic |
| Amygdala_R | 32,0,-22 | 3.02 | 95 | 8.66 | 1.95 | 0.46 | emotional; neutral; fear; fearful; disgust |
| OFCpost_R | 50,18,-12 | 3.61 | 85 | 3.77 | 2.09 | 0.36 | fear; comprised; neutral; notably; paralimbic |
| Thal_MDm_R | 4,-14,-2 | 3.51 | 82 | 8.23 | 1.73 | 0.51 | reward; mesolimbic; monetary; dopamine; tegmental |
| Putamen_R | 24,4,2 | 3.18 | 54 | 27.88 | 1.72 | 0.59 | basal ganglia; ganglia; basal; motor; primary motor |
| Amygdala_L | -22,-2,-26 | 2.95 | 54 | 10.04 | 2.40 | 0.31 | neutral; emotional; faces; fear; fearful |
| (b) HUD with methadone therapy (n=6) | | | | | | | |
| Cerebellum_Crus1_L ^a^ | -16,-76,-26 | 4.35 | 757 | 11.33 | 8.19 | 0.21 | crus; inference; online; block; personality traits |
| Frontal_Inf_Orb_2_R ^a^ | 46,32,-8 | 4.05 | 22 | 3.92 | 2.41 | 0.78 | reappraisal; emotional; social; mind; affective |
| Hypothalamus | -6,2,-12 | 3.81 | 319 | 10.87 | -3.43 | 0.77 | reward; losses; sexual; punishment; arousal |
| Cingulate_Mid_R | 2,-14,34 | 3.76 | 125 | 7.30 | 1.19 | 0.89 | retrieved; believed; control processes; comprised; personality traits |
| Parietal_Sup_L | -22,-66,50 | 3.85 | 117 | 4.65 | -0.85 | 0.93 | visual; spatial; working memory; attention network; rotation |
| Temporal_Inf_R | 62,-60,-12 | 4.19 | 114 | 4.37 | -2.99 | 0.76 | emergence; correlating; choose; predictions; extra |
| Frontal_Sup_2_L | -14,38,44 | 3.24 | 88 | 6.61 | 1.88 | 0.84 | attempt; remembering; semantic; socially; pictures |
| Thalamus | -2,-6,6 | 2.98 | 34 | 12.64 | 2.60 | 0.80 | correspond; engages; seeking; autobiographical; retrieval |
| Precuneus_L | -8,-54,16 | 3.19 | 31 | 17.64 | -8.47 | 0.53 | autobiographical; episodic; autobiographical memory; memory retrieval; episodic memory |
| Parietal_Inf_L | -46,-50,48 | 3.25 | 24 | 7.90 | 1.06 | 0.91 | network; memory; networks involved; working memory; domain general |
| Frontal_Inf_Oper_R | 50,8,16 | 3.21 | 21 | 11.66 | -5.02 | 0.68 | imitation; paced; basal ganglia; ganglia; motor task |
| (c) HUD without methadone therapy > HUD with methadone therapy | | | | | | | |
| Frontal_Sup_Medial_R ^a^ | 6,52,26 | 2.86 | 846 | 8.15 | 0.59 | 0.77 | mentalizing; theory mind; mental states; mind; mind tom |
| Cerebellum_Crus1_L ^a^ | -16,-72,-28 | -2.93 | 1262 | 2.07 | -0.69 | 0.73 | force; crus; personality traits; increasing; rehearsal |

HUD contrasts are stratified by reported methadone maintenance treatment (MMT) status; the direct comparison is observational and exploratory. Macroanatomical label: brain region name based on AAL atlas; MNI: Montreal Neurological Institute coordinates of the peak voxel (x,y,z); SDM-Z: peak z-value reflecting effect size; Voxels: cluster size; *I²* (%): heterogeneity statistic across studies; Egger’s bias and Egger’s *p*: publication bias estimates (not applicable for conjunction analyses); Function decoding: function decoding of the peak coordinates in Neurosynth. Results were significant at an uncorrected *p* < 0.005.

^a^ Survived to the FWE correction threshold of *p* < 0.05.

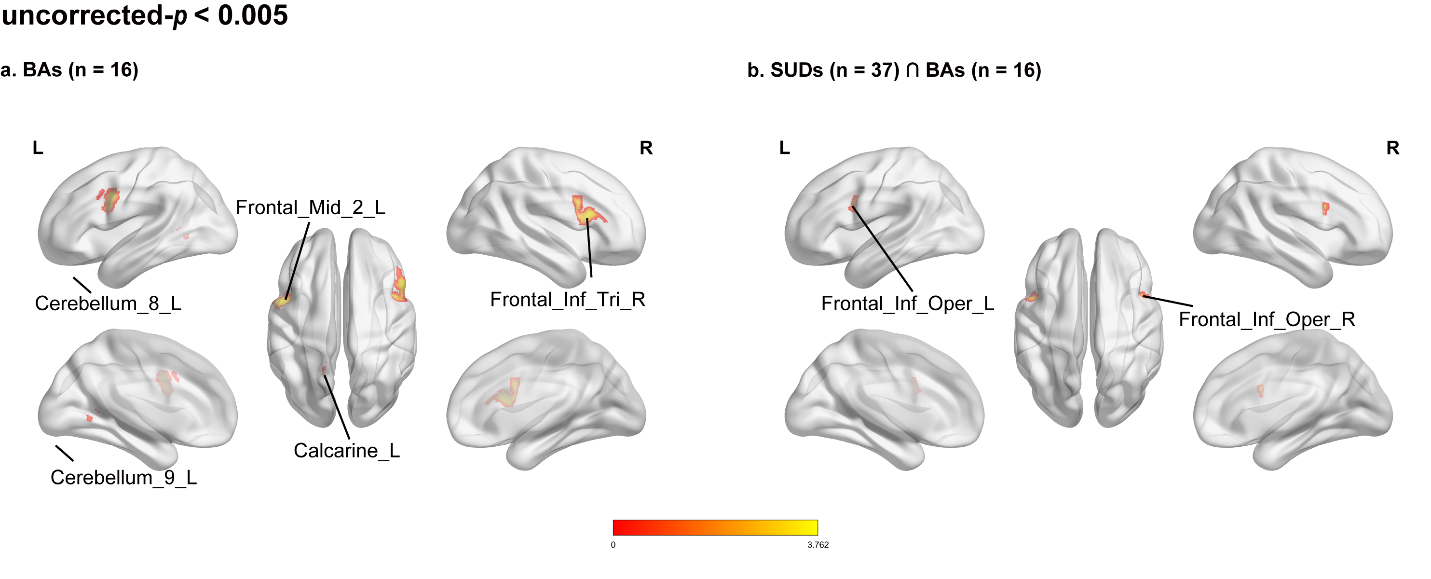

**Figure S1. Activation patterns in BAs and shared neural core across addictions**

Maps are thresholded at uncorrected p < 0.005. (a) In BAs (n = 16), annotated clusters include the left middle frontal gyrus (Frontal_Mid_2_L), left cerebellar lobules VIII and IX (Cerebellum_8_L and Cerebellum_9_L), left calcarine cortex (Calcarine_L), and right triangular inferior frontal gyrus (Frontal_Inf_Tri_R). (b) The SUDs ∩ BAs conjunction (SUDs n = 37; BAs n = 16) shows annotated clusters in the left and right opercular inferior frontal gyri (Frontal_Inf_Oper_L and Frontal_Inf_Oper_R). Maps are displayed on a standard MNI template in neurological orientation. The warm color bar indicates positive SDM-Z values; the symbol ∩ denotes conjunction.

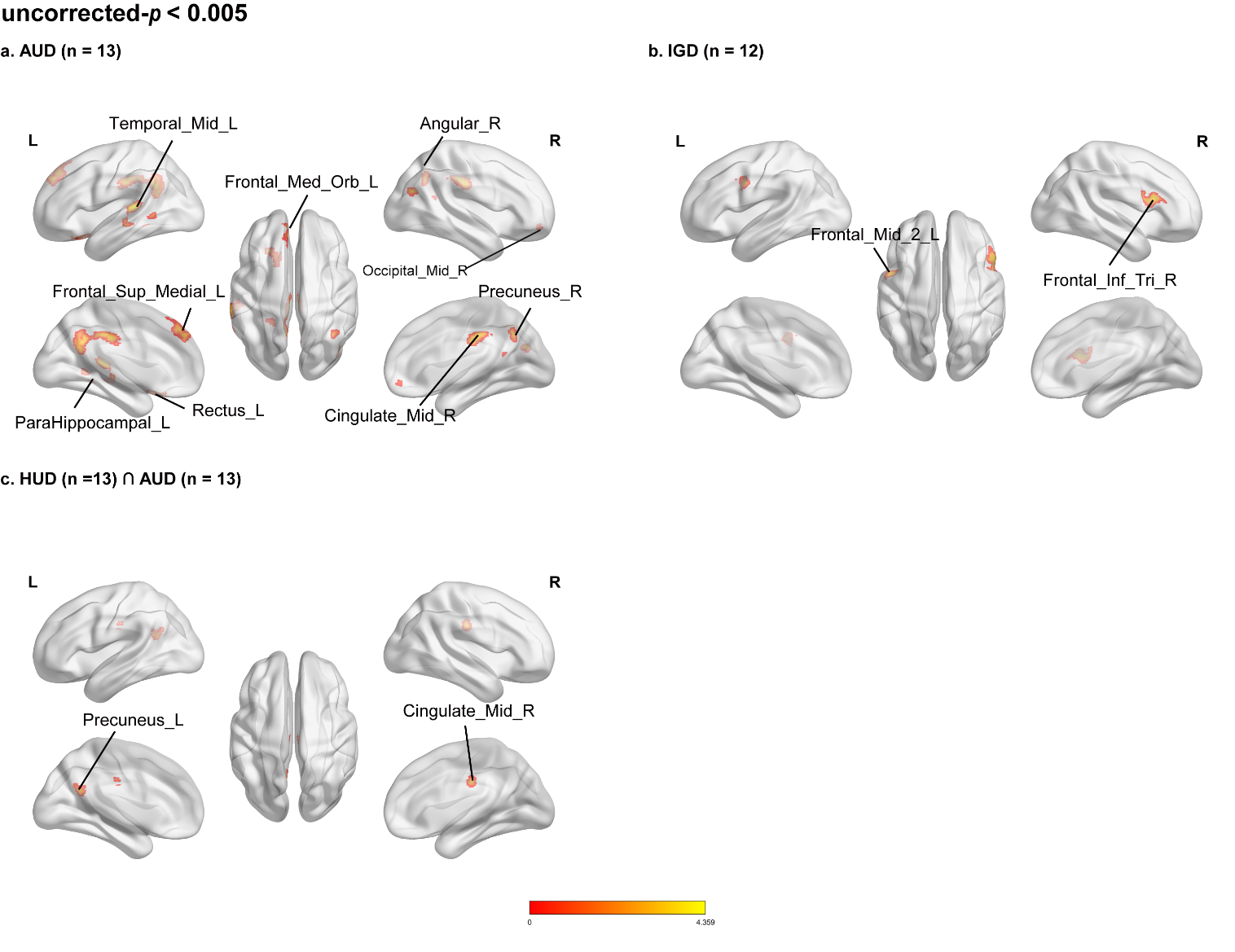

**Figure S2. Subtype-specific and shared neural activation patterns across specific addiction subtypes**

Maps are thresholded at uncorrected p < 0.005. (a) In AUD (n = 13), annotated clusters include the left middle temporal gyrus (Temporal_Mid_L), medial superior frontal gyrus (Frontal_Sup_Medial_L), parahippocampal gyrus (ParaHippocampal_L), gyrus rectus (Rectus_L), and medial orbitofrontal cortex (Frontal_Med_Orb_L), as well as the right angular gyrus (Angular_R), middle occipital gyrus (Occipital_Mid_R), precuneus (Precuneus_R), and middle cingulate gyrus (Cingulate_Mid_R). (b) In IGD (n = 12), annotated clusters include the left middle frontal gyrus (Frontal_Mid_2_L) and right triangular inferior frontal gyrus (Frontal_Inf_Tri_R). (c) The HUD ∩ AUD conjunction (n = 13 in each subgroup) shows annotated clusters in the left precuneus (Precuneus_L) and right middle cingulate gyrus (Cingulate_Mid_R). Maps are displayed on a standard MNI template in neurological orientation. The warm color bar indicates positive SDM-Z values; ∩ denotes conjunction.

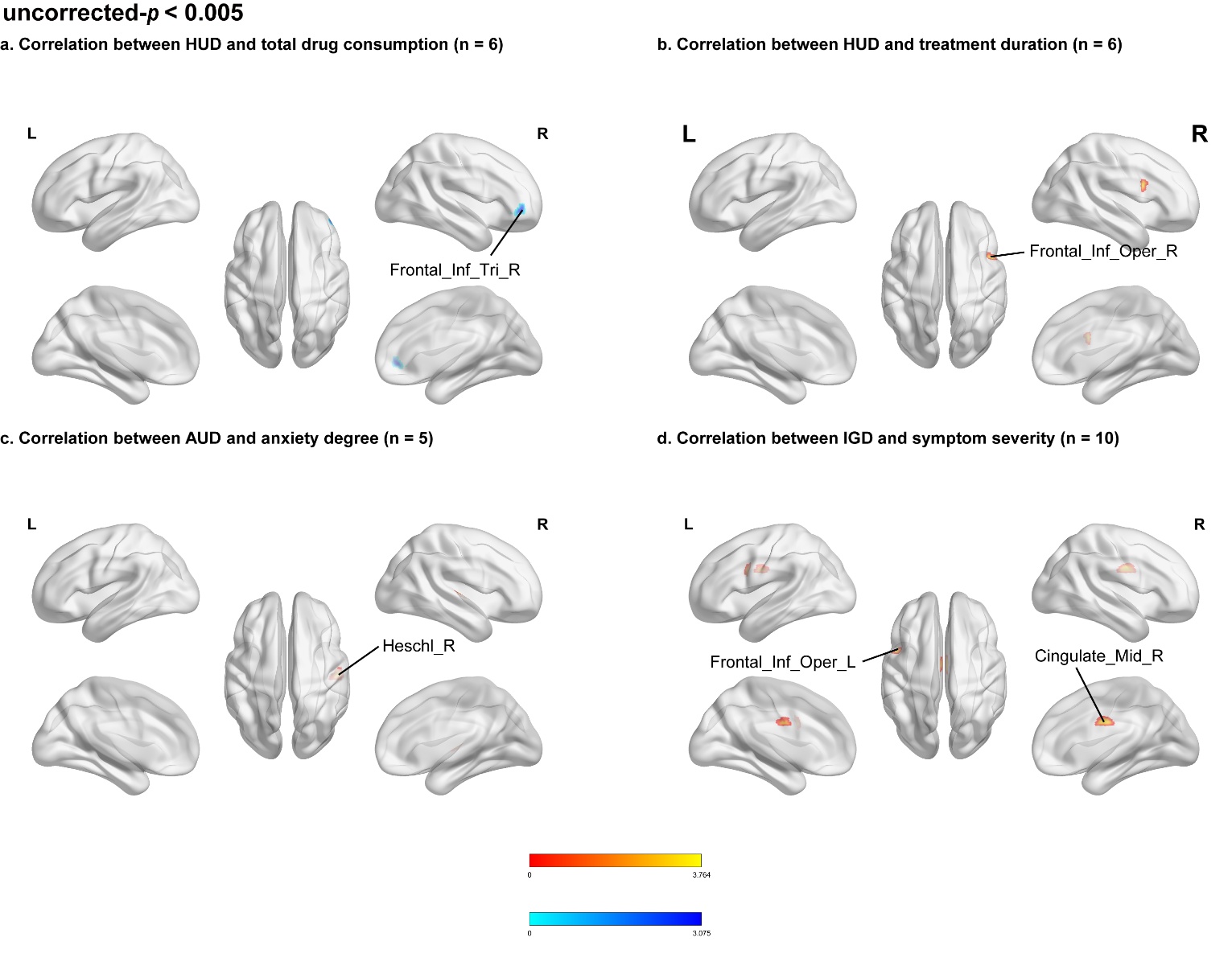

**Figure S3. Exploratory correlation between brain activation and clinical measures**

Maps are thresholded at uncorrected p < 0.005. (a) In HUD, total drug consumption (n = 6) was negatively associated with effects in the right triangular inferior frontal gyrus (Frontal_Inf_Tri_R). (b) In HUD, treatment duration (n = 6) was positively associated with effects in the right opercular inferior frontal gyrus (Frontal_Inf_Oper_R). (c) In AUD, anxiety (n = 5) was positively associated with effects in the right Heschl’s gyrus (Heschl_R). (d) In IGD, symptom severity (n = 10) was positively associated with effects in the left opercular inferior frontal gyrus (Frontal_Inf_Oper_L) and right middle cingulate gyrus (Cingulate_Mid_R). Maps are displayed on a standard MNI template in neurological orientation. Warm and cool colors indicate positive and negative associations, respectively.

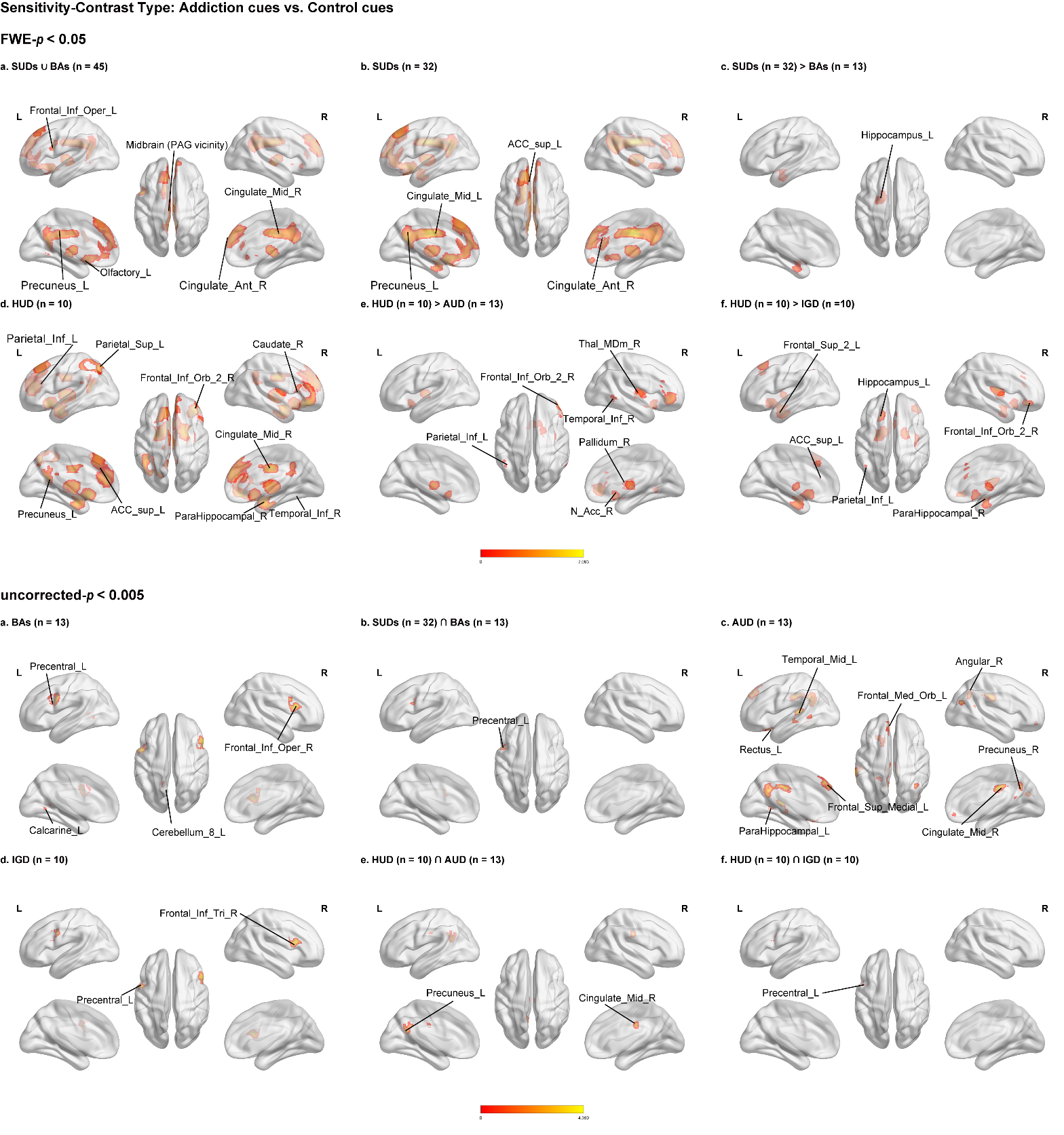

**Figure S4. Sensitivity analysis restricted to “addiction cue vs. control cue” contrasts**

The upper block shows FWE-corrected results (p < 0.05). (a) The pooled SUDs ∪ BAs analysis (n = 45) shows annotated clusters in the left opercular inferior frontal gyrus (Frontal_Inf_Oper_L), left precuneus (Precuneus_L), left olfactory cortex (Olfactory_L), right middle and anterior cingulate gyri (Cingulate_Mid_R and Cingulate_Ant_R), and a midbrain cluster (PAG vicinity). (b) In SUDs (n = 32), annotated clusters include the left supracallosal anterior cingulate cortex (ACC_sup_L), left precuneus (Precuneus_L), left middle cingulate gyrus (Cingulate_Mid_L), and right anterior cingulate gyrus (Cingulate_Ant_R). (c) SUDs (n = 32) > BAs (n = 13) shows a cluster in the left hippocampus (Hippocampus_L). (d) In HUD (n = 10), annotated clusters include the left inferior and superior parietal lobules (Parietal_Inf_L and Parietal_Sup_L), left precuneus (Precuneus_L), left supracallosal anterior cingulate cortex (ACC_sup_L), right caudate (Caudate_R), right orbital inferior frontal gyrus (Frontal_Inf_Orb_2_R), right parahippocampal gyrus (ParaHippocampal_R), right middle cingulate gyrus (Cingulate_Mid_R), and right inferior temporal gyrus (Temporal_Inf_R). (e) HUD (n = 10) > AUD (n = 13) shows annotated clusters in the left inferior parietal lobule (Parietal_Inf_L), right orbital inferior frontal gyrus (Frontal_Inf_Orb_2_R), right mediodorsal thalamus (Thal_MDm_R), right pallidum (Pallidum_R), right nucleus accumbens (N_Acc_R), and right inferior temporal gyrus (Temporal_Inf_R). (f) HUD (n = 10) > IGD (n = 10) shows annotated clusters in the left superior frontal gyrus (Frontal_Sup_2_L), left supracallosal anterior cingulate cortex (ACC_sup_L), left inferior parietal lobule (Parietal_Inf_L), left hippocampus (Hippocampus_L), right orbital inferior frontal gyrus (Frontal_Inf_Orb_2_R), and right parahippocampal gyrus (ParaHippocampal_R). The lower block shows exploratory results at uncorrected p < 0.005: (a) BAs (n = 13), (b) SUDs ∩ BAs (SUDs n = 32; BAs n = 13), (c) AUD (n = 13), (d) IGD (n = 10), (e) HUD ∩ AUD (n = 10 and n = 13, respectively), and (f) HUD ∩ IGD (n = 10 in each subgroup). Annotated regions in these panels include the precentral gyrus, calcarine cortex, cerebellar lobule VIII, triangular inferior frontal gyrus, precuneus, middle cingulate gyrus, and the AUD regions listed in panel (a). Maps are displayed on a standard MNI template in neurological orientation. Warm colors indicate positive SDM-Z values; ∪ and ∩ denote pooling and conjunction, respectively.

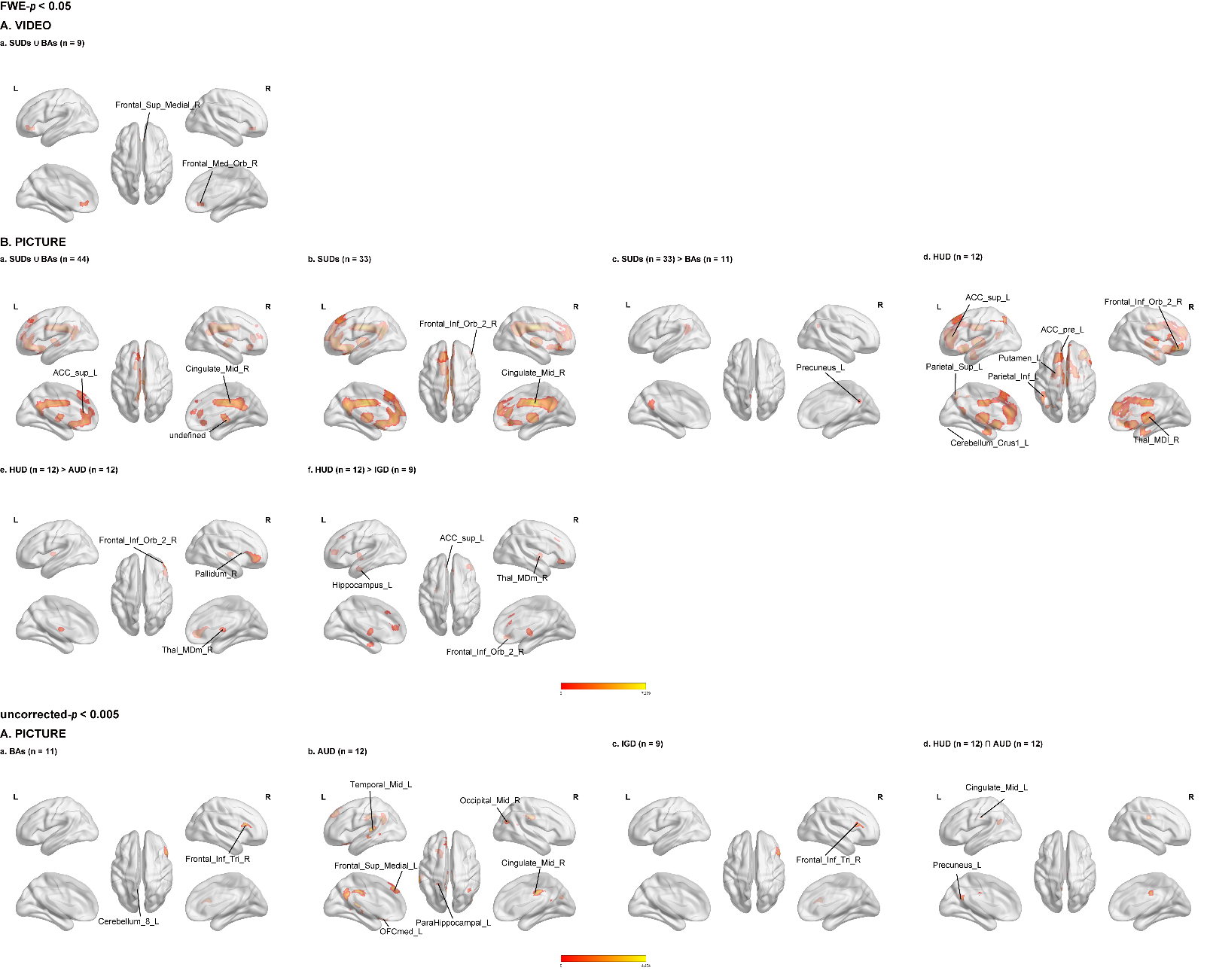

**Figure S5. Sensitivity analyses stratified by visual cue type**

The upper block shows FWE-corrected results (p < 0.05). (A) For video cues, the pooled SUDs ∪ BAs analysis (n = 9) shows annotated clusters in the right medial superior frontal gyrus (Frontal_Sup_Medial_R) and right medial orbitofrontal cortex (Frontal_Med_Orb_R). (B) For picture cues: (a) the pooled SUDs ∪ BAs analysis (n = 44) shows annotated clusters in the left supracallosal anterior cingulate cortex (ACC_sup_L), left precuneus (Precuneus_L), left olfactory cortex (Olfactory_L), right middle and anterior cingulate gyri (Cingulate_Mid_R and Cingulate_Ant_R), and an atlas-unlabeled midline cluster; (b) SUDs (n = 33) shows annotated clusters in the right orbital inferior frontal gyrus (Frontal_Inf_Orb_2_R), right middle cingulate gyrus (Cingulate_Mid_R), left precuneus (Precuneus_L), and left supracallosal anterior cingulate cortex (ACC_sup_L); (c) SUDs (n = 33) > BAs (n = 11) shows a cluster in the left precuneus (Precuneus_L); (d) HUD (n = 12) shows annotated clusters in the left supracallosal and pregenual anterior cingulate cortices (ACC_sup_L and ACC_pre_L), left inferior and superior parietal lobules (Parietal_Inf_L and Parietal_Sup_L), left putamen (Putamen_L), left cerebellar Crus I (Cerebellum_Crus1_L), right orbital inferior frontal gyrus (Frontal_Inf_Orb_2_R), and right mediodorsal thalamus (Thal_MDl_R); (e) HUD (n = 12) > AUD (n = 12) shows annotated clusters in the right orbital inferior frontal gyrus (Frontal_Inf_Orb_2_R), right pallidum (Pallidum_R), and right mediodorsal thalamus (Thal_MDm_R); and (f) HUD (n = 12) > IGD (n = 9) shows annotated clusters in the left supracallosal anterior cingulate cortex (ACC_sup_L), left hippocampus (Hippocampus_L), right orbital inferior frontal gyrus (Frontal_Inf_Orb_2_R), right mediodorsal thalamus (Thal_MDm_R), and right parahippocampal gyrus (ParaHippocampal_R). The lower block shows exploratory picture-cue results at uncorrected p < 0.005: (a) BAs (n = 11), (b) AUD (n = 12), (c) IGD (n = 9), and (d) HUD ∩ AUD (n = 12 in each subgroup). Annotated regions include the right triangular inferior frontal gyrus and left cerebellar lobule VIII in BAs; the left middle temporal gyrus, medial superior frontal gyrus, parahippocampal gyrus, medial orbitofrontal cortex, right middle occipital gyrus, and right middle cingulate gyrus in AUD; the right triangular inferior frontal gyrus in IGD; and the left middle cingulate gyrus and left precuneus in the HUD ∩ AUD conjunction. Maps are displayed on a standard MNI template in neurological orientation. Warm colors indicate positive SDM-Z values; ∪ and ∩ denote pooling and conjunction.

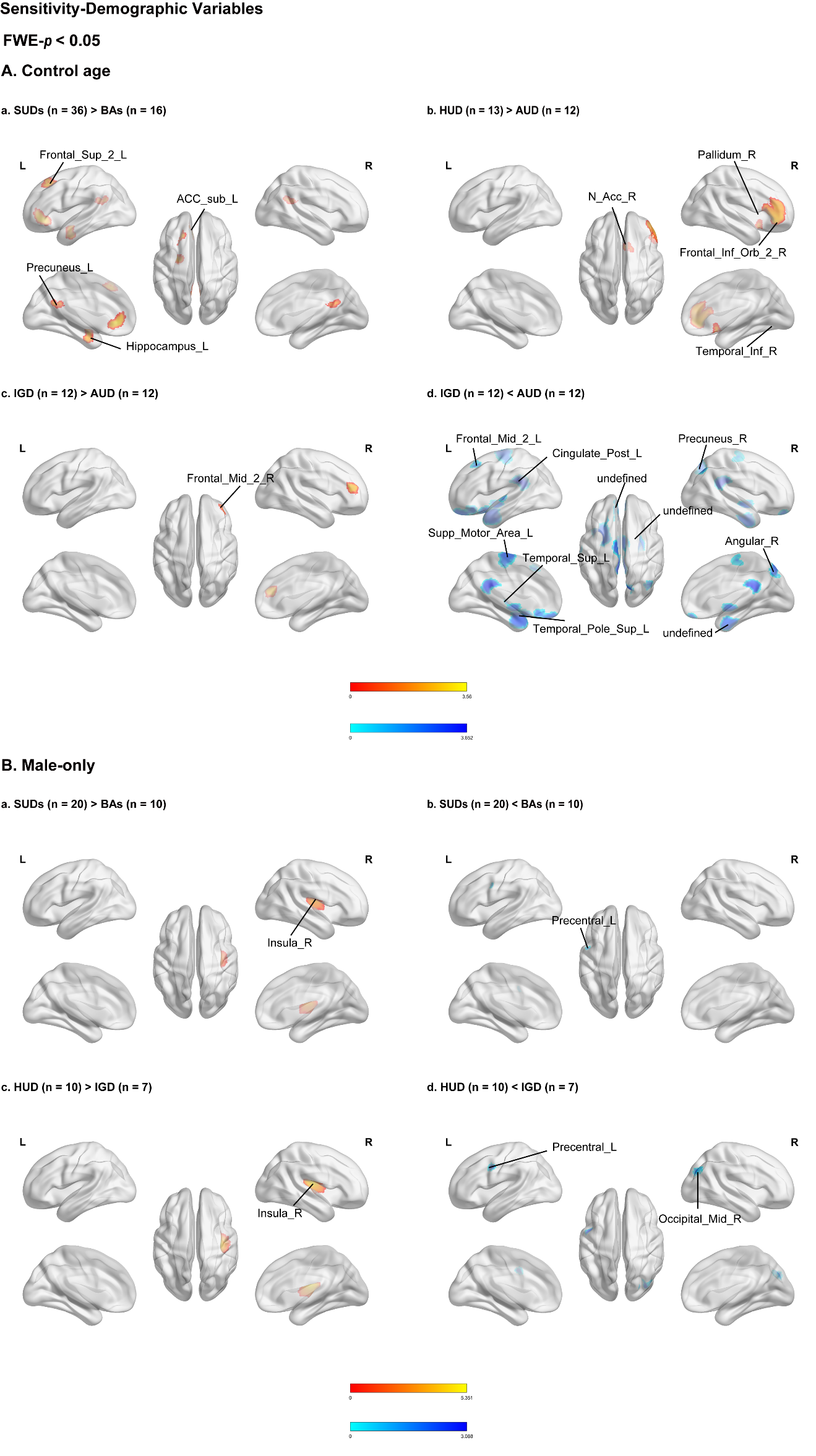

**Figure S6. Age-adjusted diagnostic comparisons and male-only sensitivity analyses**
Maps show FWE-corrected results (p < 0.05). (A) After age adjustment: (a) SUDs (n = 36) > BAs (n = 16) shows annotated clusters in the left superior frontal gyrus (Frontal_Sup_2_L), subgenual anterior cingulate cortex (ACC_sub_L), precuneus (Precuneus_L), and hippocampus (Hippocampus_L); (b) HUD (n = 13) > AUD (n = 12) shows clusters in the right nucleus accumbens (N_Acc_R), pallidum (Pallidum_R), orbital inferior frontal gyrus (Frontal_Inf_Orb_2_R), and inferior temporal gyrus (Temporal_Inf_R); (c) IGD (n = 12) > AUD (n = 12) shows a cluster in the right middle frontal gyrus (Frontal_Mid_2_R); and (d) IGD (n = 12) < AUD (n = 12) shows annotated clusters in the left middle frontal gyrus (Frontal_Mid_2_L), posterior cingulate gyrus (Cingulate_Post_L), supplementary motor area (Supp_Motor_Area_L), superior temporal gyrus (Temporal_Sup_L), superior temporal pole (Temporal_Pole_Sup_L), right precuneus (Precuneus_R), right angular gyrus (Angular_R), and atlas-unlabeled regions. (B) In male-only analyses: (a) SUDs (n = 20) > BAs (n = 10) and (c) HUD (n = 10) > IGD (n = 7) show clusters in the right insula (Insula_R); (b) SUDs (n = 20) < BAs (n = 10) shows a cluster in the left precentral gyrus (Precentral_L); and (d) HUD (n = 10) < IGD (n = 7) shows clusters in the left precentral gyrus (Precentral_L) and right middle occipital gyrus (Occipital_Mid_R). Maps are displayed on a standard MNI template in neurological orientation. Warm and cool colors indicate positive and negative differences, respectively.

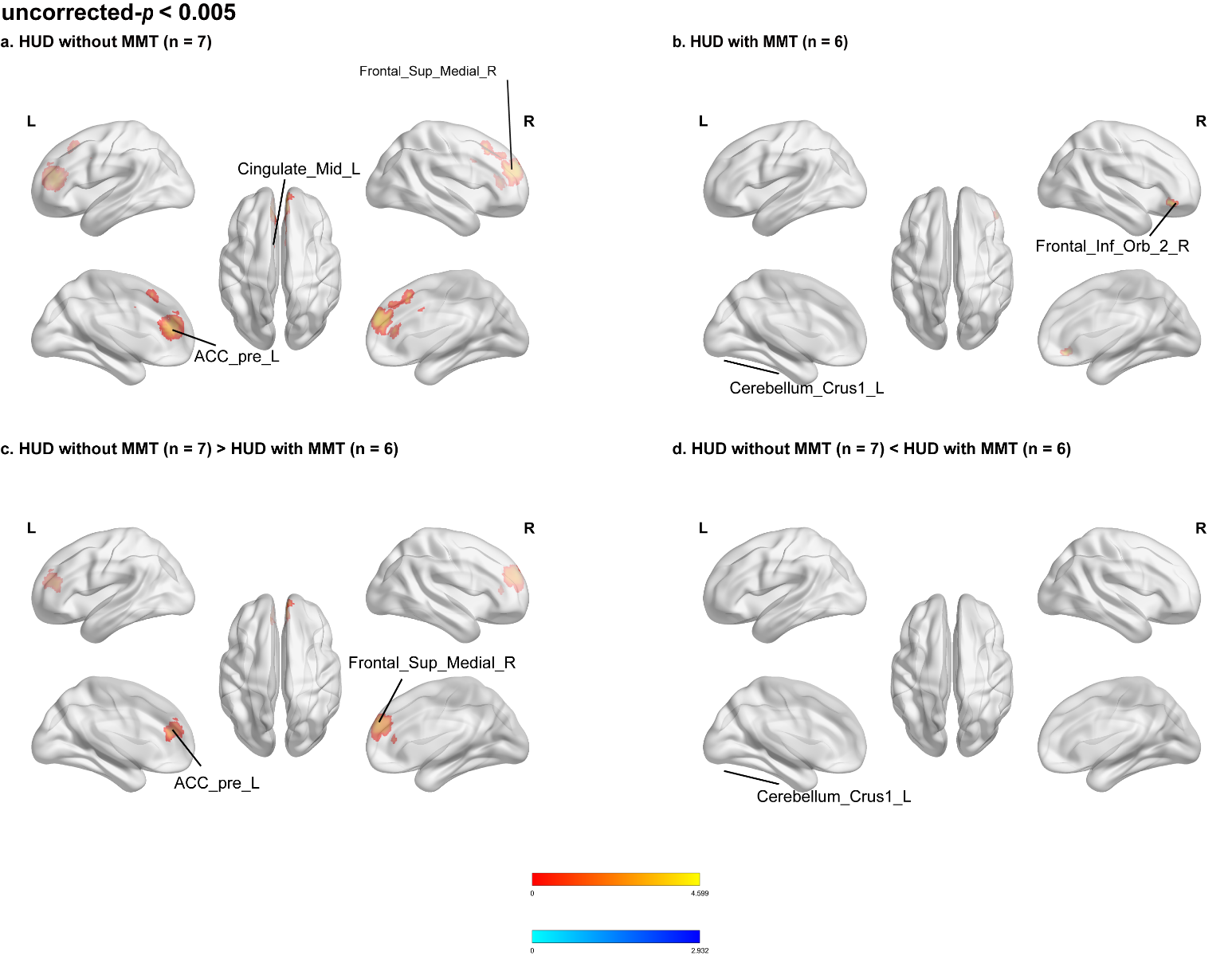

**Figure S7. Exploratory HUD analyses stratified by methadone maintenance treatment**

Maps are thresholded at uncorrected p < 0.005. (a) In HUD studies without methadone maintenance treatment (MMT; n = 7), annotated clusters include the left pregenual anterior cingulate cortex (ACC_pre_L), left middle cingulate gyrus (Cingulate_Mid_L), and right medial superior frontal gyrus (Frontal_Sup_Medial_R). (b) In HUD studies with MMT (n = 6), annotated clusters include the right orbital inferior frontal gyrus (Frontal_Inf_Orb_2_R) and left cerebellar Crus I (Cerebellum_Crus1_L). (c) The comparison of HUD without MMT (n = 7) > HUD with MMT (n = 6) shows annotated clusters in the left pregenual anterior cingulate cortex (ACC_pre_L) and right medial superior frontal gyrus (Frontal_Sup_Medial_R). (d) The reverse comparison shows a cluster in the left cerebellar Crus I (Cerebellum_Crus1_L). Maps are displayed on a standard MNI template in neurological orientation. Warm and cool colors indicate positive and negative differences, respectively.

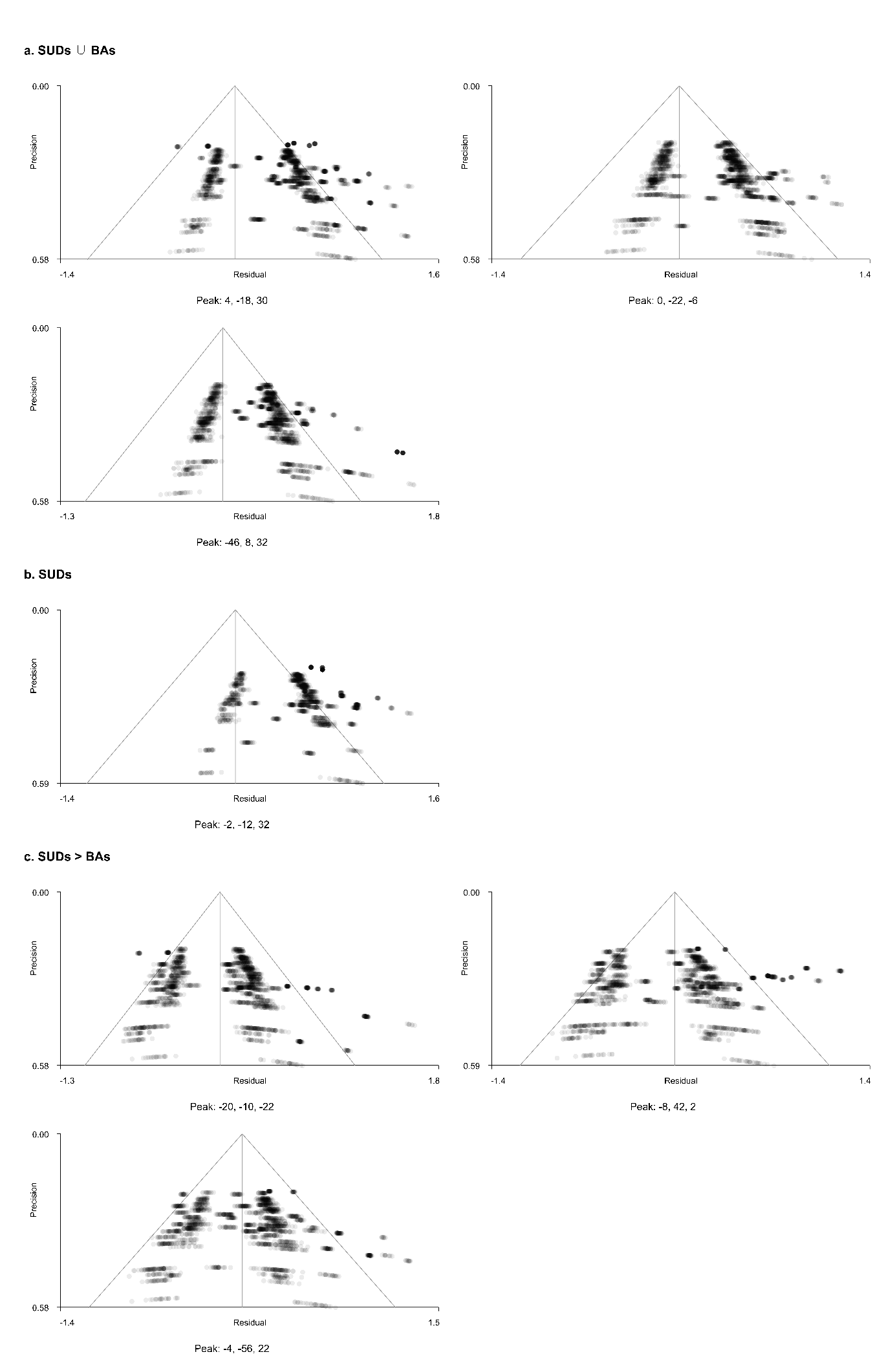

**Figure S8. Funnel plots for FWE-corrected peaks in Table S6**

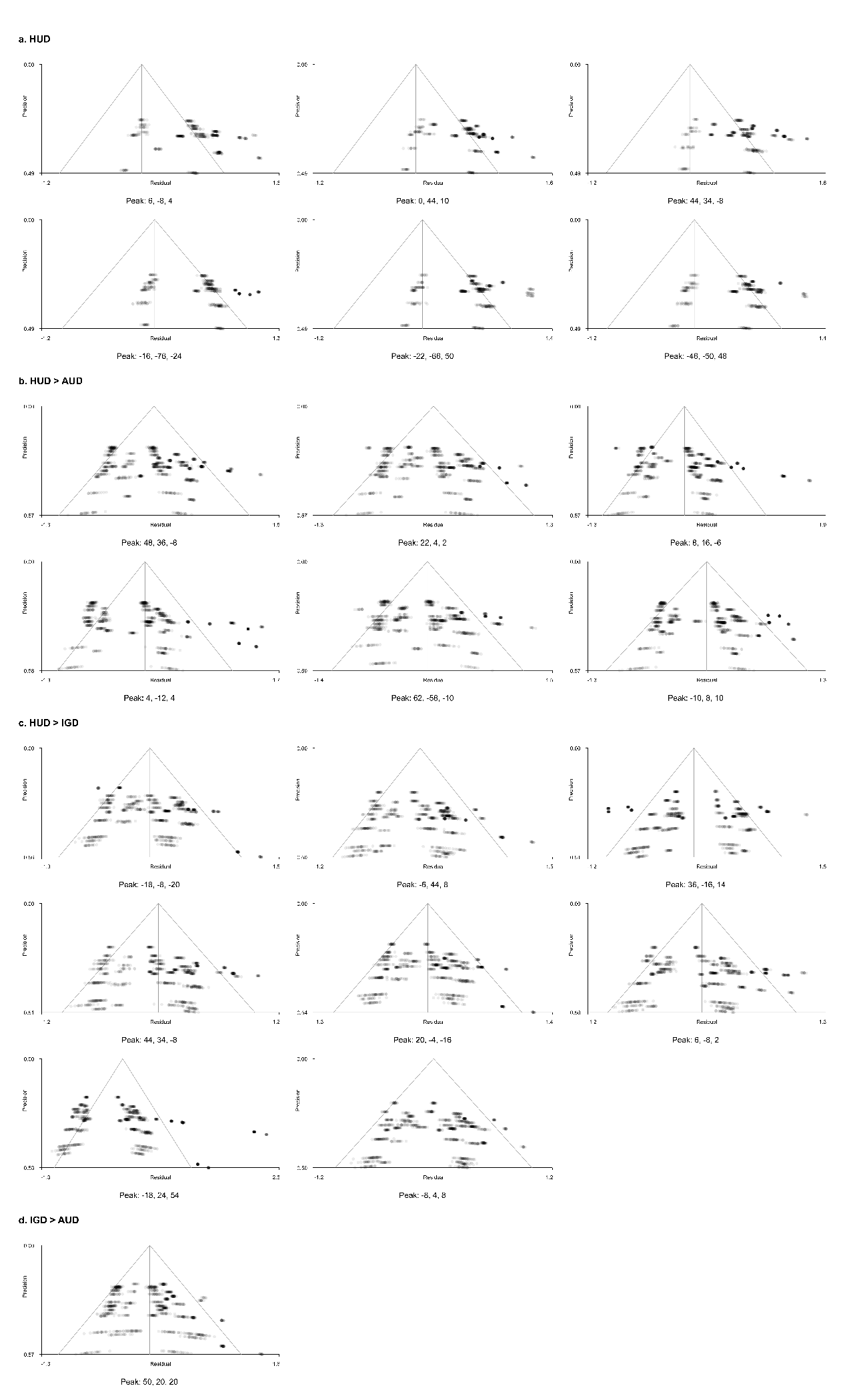

**Figure S9. Funnel plots for FWE-corrected peaks in Table S7**

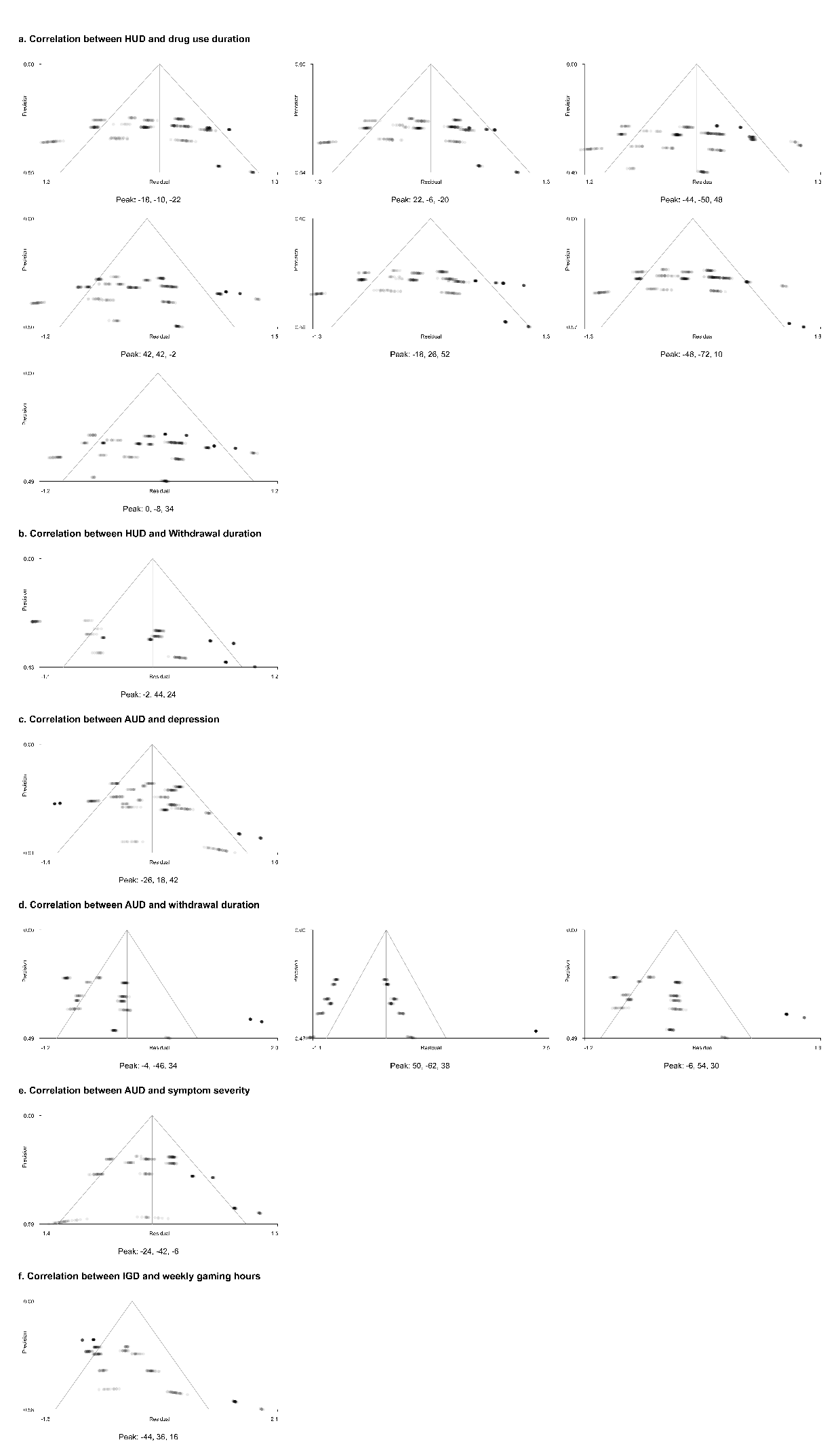

**Figure S10. Funnel plots for FWE-corrected peaks in Table S8**
